# Beyond gene-disease validity: capturing structured data on inheritance, allelic-requirement, disease-relevant variant classes, and disease mechanism for inherited cardiac conditions

**DOI:** 10.1101/2023.04.03.23287612

**Authors:** Katherine S Josephs, Angharad M Roberts, Pantazis Theotokis, Roddy Walsh, Philip J Ostrowski, Matthew Edwards, Andrew Fleming, Courtney Thaxton, Jason D Roberts, Melanie Care, Wojciech Zareba, Arnon Adler, Amy C Sturm, Rafik Tadros, Valeria Novelli, Emma Owens, Lucas Bronicki, Olga Jarinova, Bert Callewaert, Stacey Peters, Tom Lumbers, Elizabeth Jordan, Babken Asatryan, Neesha Krishnan, Ray E Hershberger, C. Anwar A. Chahal, Andrew P. Landstrom, Cynthia James, Elizabeth M McNally, Daniel P Judge, Peter van Tintelen, Arthur Wilde, Michael Gollob, Jodie Ingles, James S Ware

**Affiliations:** National Heart and Lung Institute, Imperial College London, London, UK; Royal Brompton and Harefield Hospitals, Guy’s and St Thomas’ NHS Foundation Trust, London UK; Great Ormond Street Hospital NHS Foundation Trust, London, UK; Amsterdam University Medical Centre, University of Amsterdam, Heart Center, Department of Experimental Cardiology, Amsterdam Cardiovascular Sciences, Amsterdam, The Netherlands; Clinical Genetics & Genomics Lab, Royal Brompton and Harefield Hospitals, Guy’s and St Thomas’ NHS Foundation Trust, London UK; Department of Genetics, University of North Carolina at Chapel Hill, Chapel Hill, NC, USA; Population Health Research Institute, McMaster University, and Hamilton Health Sciences, Hamilton, Ontario, Canada; Department of Molecular Genetics, University of Toronto, Toronto, Canada; Division of Cardiology, Toronto General Hospital, Toronto, Canada; Clinical Cardiovascular Research Center, University of Rochester, Rochester, New York, USA; Division of Cardiology, Peter Munk Cardiac Centre, University Health Network and Department of Medicine, University of Toronto, Toronto, ON, Canada; 23andMe, Sunnyvale, California, Genomic Health; Cardiovascular Genetics Center, Montreal Heart Institute, and Faculty of Medicine, Université de Montréal; Unit of Immunology and Functional Genomics, Centro Cardiologico Monzino IRCCS, Milano, Italy; CHEO Research Institute, University of Ottawa, Ontario, Canada; Department of Genetics, CHEO, Ontario, Canada; Center for Medical Genetics, Ghent University Hospital; Department of Biomolecular Medicine, Ghent University; Department of Cardiology and Genomic Medicine, Royal Melbourne Hospital, Melbourne, Australia; University of Melbourne, Melbourne, Australia; Barts Health & University College London Hospitals NHS Trusts, London, UK; Institute of Health Informatics, University College London, London, UK; Division of Human Genetics, The Ohio State University, Columbus, Ohio USA; Department of Cardiology, Inselspital, Bern University Hospital, University of Bern, Bern, Switzerland; Division of Cardiology, Department of Medicine, Johns Hopkins University School of Medicine, Baltimore, MD, USA; Centre for Population Genomics, Garvan Institute of Medical Research, and UNSW Sydney, Sydney, Australia; Center for Inherited Cardiovascular Diseases, WellSpan Health, Lancaster, PA USA; Cardiac Electrophysiology and Inherited Cardiovascular Diseases, Cardiovascular Division, Hospital of the University of Pennsylvania, Philadelphia, PA USA; Department of Cardiovascular Medicine, Mayo Clinic, Rochester, MN USA; Barts Heart Centre, St Bartholomew’s Hospital, Barts Health NHS Trust, London, UK; Department of Pediatrics and Cell Biology, Duke University School of Medicine, Durham, North Carolina, US; Johns Hopkins Center for Inherited Heart Diseases, Department of Medicine, Johns Hopkins University, Baltimore, Maryland, US; Center for Genetic Medicine, Dept of Medicine (Cardiology), Northwestern University Feinberg School of Medicine, Chicago, IL US; Medical University of South Carolina, Charleston, SC USA; Department of Genetics, University Medical Center Utrecht, Utrecht, the Netherlands; Amsterdam UMC location University of Amsterdam, Department of Cardiology, Meibergdreef 9, Amsterdam, the Netherlands; Amsterdam Cardiovascular Sciences, Heart Failure and arrhythmias, Amsterdam, the Netherlands; Inherited Arrhythmia and Cardiomyopathy Program, Division of Cardiology, University of Toronto, Toronto ON Canada; MRC London Institute of Medical Sciences, Imperial College London, London, UK

**Keywords:** Inherited cardiac conditions, inheritance, allelic requirement, disease mechanism, gene curation, genomic variant filtering, variant interpretation, variant classification

## Abstract

**Background:** As availability of genomic testing grows, variant interpretation will increasingly be performed by genomic generalists, rather than domain-specific experts. Demand is rising for laboratories to accurately classify variants in inherited cardiac condition (ICC) genes, including as secondary findings.

**Methods:** We analyse evidence for inheritance patterns, allelic requirement, disease mechanism and disease-relevant variant classes for 65 ClinGen-curated ICC gene-disease pairs. We present this information for the first time in a structured dataset, CardiacG2P, and assess application in genomic variant filtering.

**Results:** For 36/65 gene-disease pairs, loss-of-function is not an established disease mechanism, and protein truncating variants are not known to be pathogenic. Using CardiacG2P as an initial variant filter allows for efficient variant prioritisation whilst maintaining a high sensitivity for retaining pathogenic variants compared with two other variant filtering approaches.

**Conclusions:** Access to evidence-based structured data representing disease mechanism and allelic requirement aids variant filtering and analysis and is pre-requisite for scalable genomic testing.

## Background

Inherited cardiac conditions (ICCs) are a group of disorders that share the potential for devastating outcomes, including heart failure and sudden cardiac death at a young age.

Early diagnosis is vital and allows prompt treatment, risk stratification, and primary prevention for sudden cardiac arrest in high-risk individuals. Genetic testing is a routine part of evaluation and can aid diagnosis and alter clinical management(1–3).

The scope of genetic testing for ICC-associated genes is growing. In addition to patients undergoing evaluation for confirmed or suspected disease, opportunistic screening for secondary findings is increasing as more patients undergo exome (ES) or genome sequencing (GS) in diverse clinical settings or via consumer-initiated testing. There are 47 of 90 medically actionable gene-disease pairs on the American College of Medical Genetics and Genomics Secondary Findings list (ACMG SF V3.1)(4)related to cardiovascular (CV) disease. The ACMG recommends that these genes are analysed whenever clinical ES or GS is performed, and that pathogenic or likely pathogenic (P/LP) variants are reported back to patients. Therefore, many laboratories, regardless of their expertise, will soon need the capability to rapidly interpret variants in CV genes. This creates the potential for variant misclassification and/or poor communication of the interpretation of secondary findings to clinicians which could have significant downstream effects on patients and their families(5).

As access to sequencing and sharing of genomic data has improved, the number of genes and variants reported to be associated with any given disease has grown. Bioinformatic filtering pipelines often prioritise protein truncating variants that are indeed enriched for disease-causing variants in aggregate, but may not be pathogenic if loss-of-function (LoF) is not a mechanism for the relevant disease. At best, this results in time consuming false positives and, at worst, can lead to misinterpretation of genomic test results. For ICCs, incomplete penetrance, genetic heterogeneity, oligogenic and modifying variants, overlapping phenotypes, and different disease mechanisms make variant interpretation particularly challenging.

There are international efforts underway to re-evaluate the validity of previously published gene-disease relationships. The Gene Curation Coalition (GenCC)(6) is a consortium of parties engaged in gene curation, and theGenCC.org (https://thegencc.org/) is a harmonised repository of curated gene-disease relationships from many groups. Having established a robust gene-disease relationship, clinical interpretation of variation within a disease gene is critically dependent on an understanding of the allelic requirement for the disease, and of the mechanism of pathogenicity and disease-relevant variant classes. This data has not previously been consistently available in a structured format for variant prioritisation.

Here, we have analysed the inheritance, allelic requirement, disease mechanism, and disease-relevant variant classes for robust ICC-associated gene-disease pairs using a standardised terminology recently developed by the GenCC(7). The results of this analysis have been approved by international multidisciplinary expert review panels comprised of scientists and clinicians with expertise in ICCs. Structured data sets with this type of information do not exist currently and are shared here and as a publicly available resource, CardiacG2P, to aid filtering and analysis of ICC genetic variants.

CardiacG2P is an evidence-based dataset hosted on G2P (https://www.ebi.ac.uk/gene2phenotype/downloads), an online system set up to establish, curate and distribute datasets for diagnostic variant filtering(8). Each dataset entry annotates a disease with an allelic requirement, information pertaining to disease mechanism (represented as a disease-associated variant consequence), and known disease-relevant variant classes at a defined locus. This dataset is compatible with the existing G2P Ensembl Variant Effect Predictor (VEP)(9) plugin to support automated filtering of genomic variants accounting for inheritance pattern and mutational consequence. Other G2P datasets for developmental disorders and ophthalmic conditions have shown this approach can help to discriminate between variants, improving the precision of diagnostic variant filtering(8,10). G2P data are also available through the GenCC hub [theGenCC.org]. Here we assess CardiacG2P and show its impact on the efficiency of variant prioritisation.

## Methods

### Analysis of inheritance and disease-associated variant consequences in genes implicated in inherited cardiac conditions

We analysed evidence to determine the inheritance pattern, allelic requirement, disease mechanism, and disease-relevant variant classes for 65 gene-disease pairs for major ICCs (Figure 1). We analysed genes classified with “Definitive” or “Strong” evidence by The Clinical Genome Resource(11) (ClinGen) Gene Curation Expert Panels (GCEPs) for seven CV diseases under a Mendelian (monogenic) model (accessed November 2020): Hypertrophic cardiomyopathy (HCM), Dilated cardiomyopathy (DCM), Arrhythmogenic right ventricular cardiomyopathy (ARVC), Long QT Syndrome (LQTS), Brugada syndrome (BrS), Catecholaminergic polymorphic ventricular tachycardia (CPVT), and Short QT syndrome (SQTS)(12–17). Information on these ClinGen expert panels, membership, and curation activity can be found at www.clinicalgenome.org. For HCM, we included both genes causing typical HCM, and also genes associated with syndromic disorders where apparently isolated left ventricular hypertrophy (LVH) may be the presenting feature (genocopies)(16) Seven channelopathy gene-disease pairs classified by ClinGen as having “Moderate” strength of evidence for monogenic disease are included (*CALM1-*CPVT*, CALM2-*CPVT*, CALM3-*CPVT*, CASQ2-*CPVT*, KCNE1-*JLN*, SLC4A3*-SQTS, *KCNJ2*-SQTS), following discussion with the channelopathy expert review panel for this project, and where there was sufficient data to adjudicate the required fields. *SLC22A5* was also evaluated as a phenotypic mimic of SQTS: although it is classified as “Disputed” by ClinGen Short QT GCEP in relation to true SQTS, it is definitively associated with systemic primary carnitine deficiency disease, which can present similarly to SQTS and might reasonably be included in gene panels for diagnostic assessment of patients presenting with this phenotype. See Table 1, Table 2 and Additional File 1 for a complete list of the gene-disease pairs evaluated.

**Figure 1:**
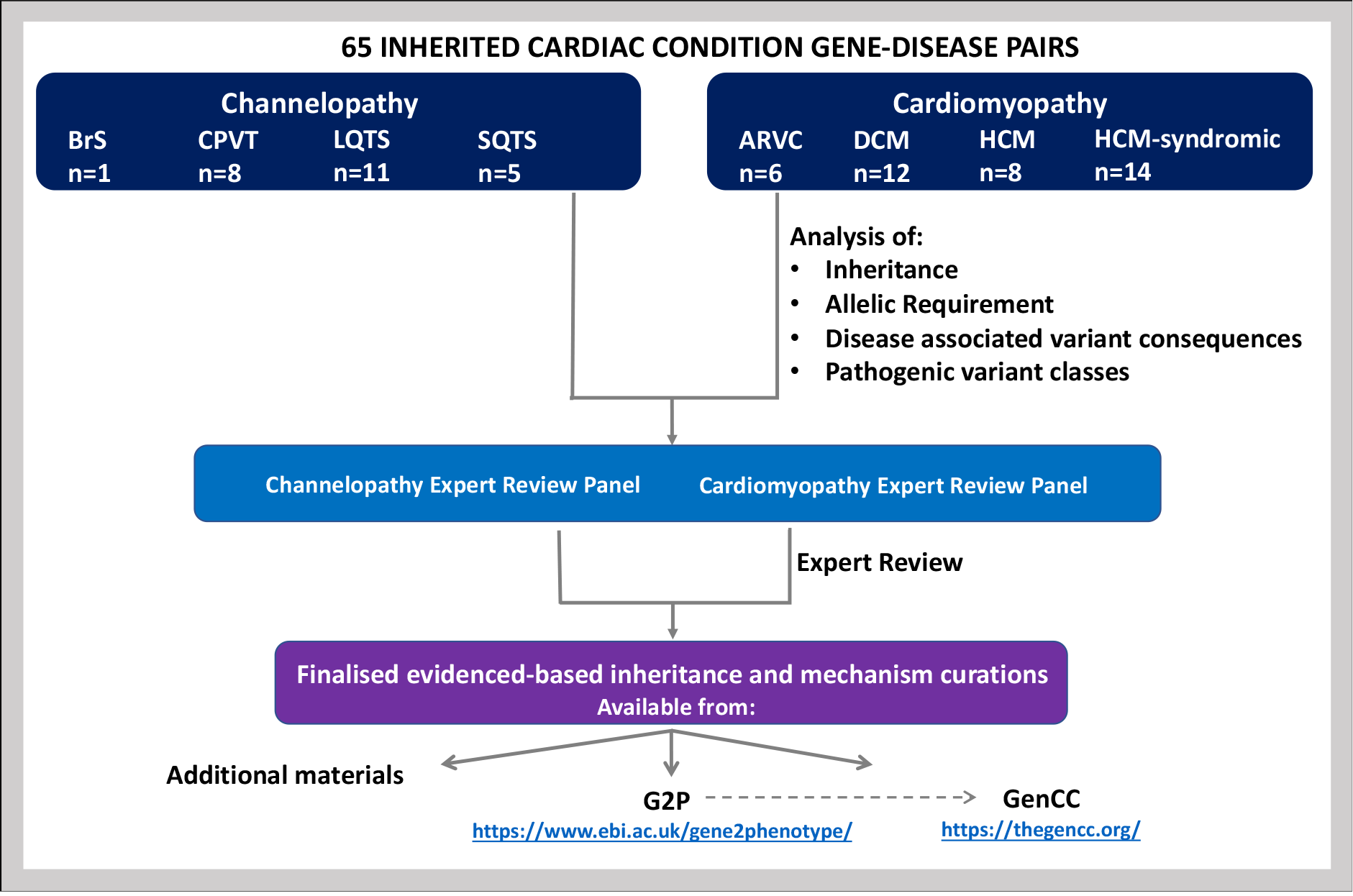
Flow chart depicting the analysis of inheritance and disease mechanism in established inherited cardiac genes. A structured representation of the resulting data is available in the Additional Materials, and also through G2P (https://www.ebi.ac.uk/gene2phenotype/downloads), which is also searchable through the GenCC portal (https://thegencc.org/). *ARVC – arrhythmogenic right ventricular cardiomyopathy, BrS – Brugada syndrome, CPVT – catecholaminergic polymorphic ventricular tachycardia, DCM – dilated cardiomyopathy, G2P – gene2phenotype, GenCC – Gene Curation Coalition, HCM – hypertrophic cardiomyopathy, LQTS – long QT syndrome, SQTS – short QT syndrome*.

**Table 1:**
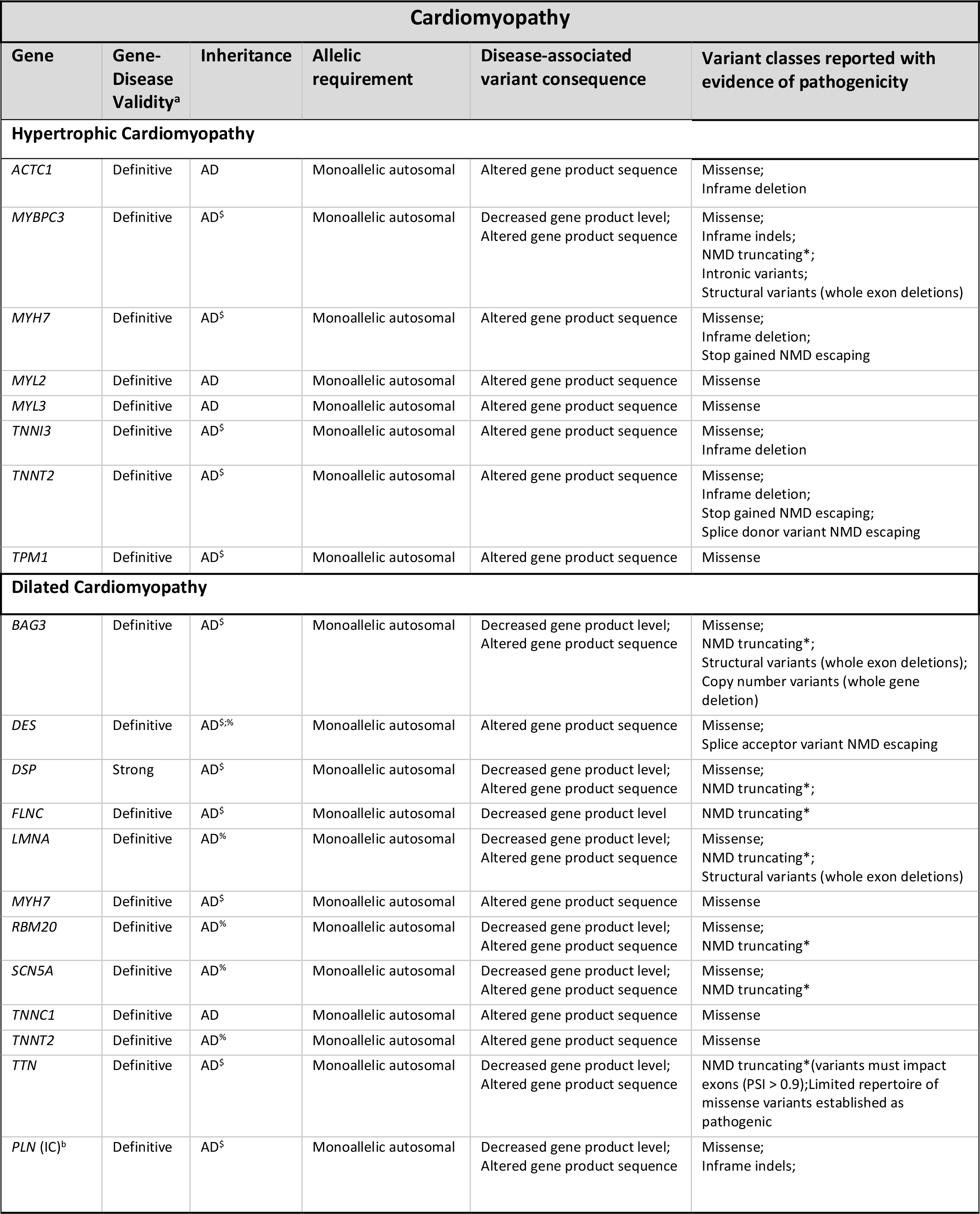
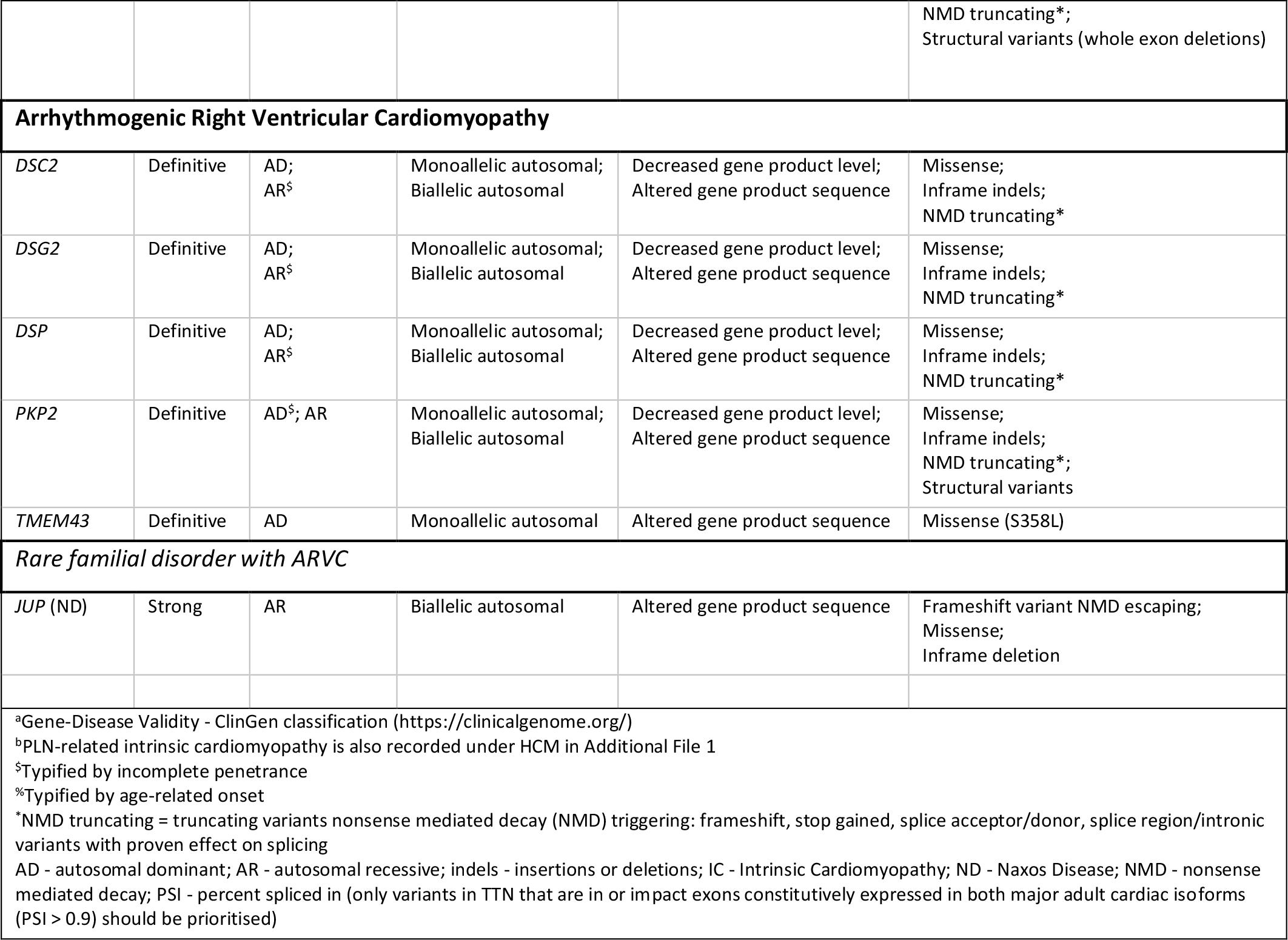
Structured representation of data from curation of core cardiomyopathy gene-disease pairs (HCM, DCM, ARVC)

**Table 2:** Structured representation of data from curation of channelopathy gene-disease pairs (LQTS, SQTS, CPVT, BrS)

| Channelopathy |  |  |  |  |  |
| --- | --- | --- | --- | --- | --- |
| Gene | Gene-Disease Validity<sup>a</sup> | Inheritance | Allelic requirement | Disease-associated variant consequence | Variant classes reported with evidence of pathogenicity |
| **Long QT Syndrome (LQTS)** |  |  |  |  |  |
| *Familial Long QT Syndrome* |  |  |  |  |  |
| KCNQ1 | Definitive | AD;<br>AR<sup>§</sup> | Monoallelic autosomal;<br>Biallelic autosomal | Decreased gene product level;<br>Altered gene product sequence | Missense;<br>Inframe indels;<br>NMD truncating*;<br>Structural variants (multi exon deletions and a duplication) |
| KCNH2 | Definitive | AD<sup>§</sup> | Monoallelic autosomal | Decreased gene product level;<br>Altered gene product sequence | Missense;<br>Inframe indels;<br>NMD truncating*;<br>Structural variants (whole exon deletions and duplications) |
| SCN5A | Definitive | AD<sup>§</sup> | Monoallelic autosomal | Altered gene product sequence | Missense;<br>Inframe indels |
| *Long QT Syndrome with atypical features* |  |  |  |  |  |
| CALM1 | Definitive | AD<sup>&</sup> | Monoallelic autosomal | Altered gene product sequence | Missense |
| CALM2 | Definitive | AD<sup>&</sup> | Monoallelic autosomal | Altered gene product sequence | Missense |
| CALM3 | Definitive | AD<sup>&</sup> | Monoallelic autosomal | Altered gene product sequence | Missense |
| TRDN | Strong | AR<sup>&</sup> | Biallelic autosomal | Absent gene product level;<br>Altered gene product sequence | NMD truncating*;<br>Missense |
| *Syndrome with QT prolongation and cardiac arrhythmias* |  |  |  |  |  |
| KCNQ1 (JLNS) | Definitive | AR | Biallelic autosomal | Absent gene product level;<br>Altered gene product sequence | Missense;<br>Inframe indels;<br>NMD truncating*;<br>Structural variants (whole exon deletions)<br>Complex rearrangements |
| KCNE1 (JLNS) | Moderate | AR | Biallelic autosomal | Altered gene product sequence | Missense;<br>Inframe indels;<br>Stop gained NMD escaping |
| KCNJ2 (ATS) | Definitive | AD | Monoallelic autosomal | Altered gene product sequence | Missense;<br>Inframe indels;<br>Stop gained NMD escaping |
| CACNA1C (TS) | Definitive | AD<sup>&</sup> | Monoallelic autosomal | Altered gene product sequence | Missense |
| **Brugada Syndrome (BrS)** |  |  |  |  |  |
| SCN5A | Definitive | AD<sup>§</sup> | Monoallelic autosomal | Decreased gene product level;<br>Altered gene product sequence | Missense;<br>Inframe indels;<br>NMD truncating* |
| **Catecholaminergic Polymorphic Ventricular Tachycardia (CPVT)** |  |  |  |  |  |
| *Classic CPVT Phenotype* |  |  |  |  |  |
| RYR2 | Definitive | AD<sup>§</sup> | Monoallelic autosomal | Altered gene product sequence | Missense;<br>Structural variants (exon 3 deletion) |
| CASQ2 | Definitive | AR | Biallelic autosomal | Absent gene product level;<br>Altered gene product<br>sequence | Missense;<br>NMD truncating* |
| CASQ2 | Moderate | AD<sup>§</sup> | Monoallelic autosomal | Decreased gene product level;<br>Altered gene product<br>sequence | Missense;<br>NMD truncating* |
| **Atypical CPVT Phenotype** |  |  |  |  |  |
| CALM1 | Moderate | AD<sup>&</sup> | Monoallelic autosomal | Altered gene product<br>sequence | Missense |
| CALM2 | Moderate | AD<sup>&</sup> | Monoallelic autosomal | Altered gene product<br>sequence | Missense |
| CALM3 | Moderate | AD<sup>&</sup> | Monoallelic autosomal | Altered gene product<br>sequence | Missense |
| TRDN | Definitive | AR | Biallelic autosomal | Absent gene product level;<br>Altered gene product<br>sequence | Missense;<br>NMD truncating*;<br>Structural variants (exon 2 deletion) |
| TECRL | Definitive | AR | Biallelic autosomal | Absent gene product level;<br>Altered gene product<br>sequence | Missense;<br>NMD truncating*;<br>Structural variants (exon 2 deletion) |
| **Short QT Syndrome (SQTS)** |  |  |  |  |  |
| *Classic SQTS* |  |  |  |  |  |
| KCNH2 | Definitive | AD | Monoallelic autosomal | Altered gene product<br>sequence | Missense |
| KCNQ1 | Strong | AD<sup>&</sup> | Monoallelic autosomal | Altered gene product<br>sequence | Missense |
| SLC4A3 | Moderate | AD | Monoallelic autosomal | Altered gene product<br>sequence | Missense |
| KCNJ2 | Moderate | AD | Monoallelic autosomal | Altered gene product<br>sequence | Missense |
| **Syndrome including shortened QT and cardiac arrhythmias** |  |  |  |  |  |
| SLC22A5<br>(PSCD) | Definitive | AR | Biallelic autosomal | Altered gene product<br>sequence | Missense |
| <sup>a</sup>Gene-Disease Validity - ClinGen classification (https://clinicalgenome.org/)<br><sup>§</sup>Typified by incomplete penetrance<br><sup>&</sup>Typically de novo<br>AD - autosomal dominant; AR - autosomal recessive; ATS - Andersen-Tawil Syndrome; indels – insertions or deletions; JLNS – Jervell and Lange-Nielsen Syndrome; NMD – nonsense mediated decay; PSCD – primary systemic carnitine deficiency; TS – Timothy Syndrome<br>*NMD truncating = truncating variants nonsense mediated decay (NMD) triggering: frameshift, stop gained, splice acceptor/donor, splice region/intronic variants with proven effect on splicing |  |  |  |  |  |

Inheritance, allelic requirement, and disease-associated variant consequences (as a proxy for disease mechanism), are described using previously agreed standardised terms developed by the GenCC (https://thegencc.org/)(7). These terms are formalised in the sequence ontology (SO) (http://www.sequenceontology.org) and human phenotype ontology (HPO) (https://hpo.jax.org/app/). Briefly, since the precise disease mechanism is not always known, six high-level variant-consequence terms are used to describe disease-associated variant consequences. These are assigned depending on which variant classes are associated with disease (see Tables 2 and 3 in Roberts AM et al(7)). As examples, “decreased gene product level” [SO:0002316] is used when disease is caused by variants that decrease the level or amount of gene product produced (e.g. variants leading to premature termination codons (PTCs) that trigger nonsense mediated decay (NMD), and gene deletions) and “altered gene product sequence” [SO:0002318] is used for non-truncating variants that instead alter the sequence of the gene product such as the amino acid sequence of a protein (e.g. missense variants, inframe insertions or deletions (indels), PTCs predicted to escape NMD, and stop loss). Variants producing PTCs are often referred to as “loss of function (LoF)” variants, but a PTC could lead to LoF, gain of function (GoF) through loss of a terminal regulatory region, or dominant negative effect. Similarly missense variants can cause GoF, LoF, or dominant negative effects. Using known pathogenic variant classes to describe which consequences, at a sequence level, have been associated with disease allows prediction of which other variant classes may be pathogenic whilst recognising that the downstream mechanisms following a particular sequence consequence can be diverse(7). More than one disease-associated variant consequence term can be used for each gene-disease pair.

Evidence was collected primarily from published, peer-reviewed literature, but also publicly accessible resources such as ClinGen and variant databases (e.g., ClinVar). Building on the previous work by ClinGen GCEPs to determine gene-disease validity, each gene-disease pair was analysed by an individual curator following a standard operating procedure for determining inheritance and disease-associated variant consequences (see Additional File 2). Curation results were then reviewed by panels of international experts (clinicians and scientists) drawn from the relevant disease area.

### Development of CardiacG2P

A structured representation of the resulting data is available in the Additional Materials, and also through G2P (https://www.ebi.ac.uk/gene2phenotype/downloads), which is also searchable through the GenCC portal (https://thegencc.org/).

For each curation entry, a gene or locus is linked to a disease via a disease associated variant consequence (as a proxy for disease mechanism) and allelic requirement. Additional information including a confidence category of gene-disease validity (as previously assigned by ClinGen), a narrative summary describing key messages from the expert review, and relevant publication identifiers is also stored.

Unless specifically mentioned, genes previously curated for validity by ClinGen, but not classified as “Definitive” or “Strong” for cardiac disease are included on the panel for completeness. The panel reports the gene-disease validity classification (e.g., “Limited” evidence), but does not speculate on inheritance and mechanism terms where the gene- disease relationship is not established (for information on the current version of the ClinGen gene-disease validity SOP see: https://clinicalgenome.org/curation-activities/gene-disease-validity/training-materials/).

### Validating CardiacG2P

We evaluated the utility of CardiacG2P by comparing a variant prioritisation pipeline incorporating data from this structured resource against two alternative generic approaches available to an analyst without disease-specific expertise (see Figure 2). All three pipelines interrogate the same gene list which includes the 21 HCM and 12 DCM genes evaluated here.

● **Pipeline 1:** Generic bioinformatics analysis pipeline with 3-step filtering approach: filtering on gene symbol (for 33 gene-disease relationships classified by ClinGen as “Strong” or “Definitive” for HCM and/or DCM), retaining only rare variants (gnomAD(18) global allele frequency <0.0001), retaining only protein altering variants (PAVs).
● **Pipeline 2:** Generic bioinformatics analysis pipeline with 4-step filtering approach: on gene symbol, retaining only rare variants (gnomAD global allele frequency <0.0001), retaining variants that are either high impact (i.e. protein truncating variants (e.g. stop gained, frameshift) AND predicted to result in loss of function with high confidence by LOFTEE(18), a VEP plugin), OR that are previously classified in ClinVar as P/LP (as annotated by VEP).
● **Pipeline 3 (Cardiac G2P):** Using CardiacG2P dataset, variants were filtered: on gene symbol, retaining only rare variants (gnomAD global allele frequency <0.0001), and with allelic requirement, variant consequence, and gene specific annotations of a restricted repertoire of pathogenic alleles all appropriate for the disease under interrogation – e.g. restricted variant classes, specific variants, or restricted regions of the protein.

**Figure 2:**
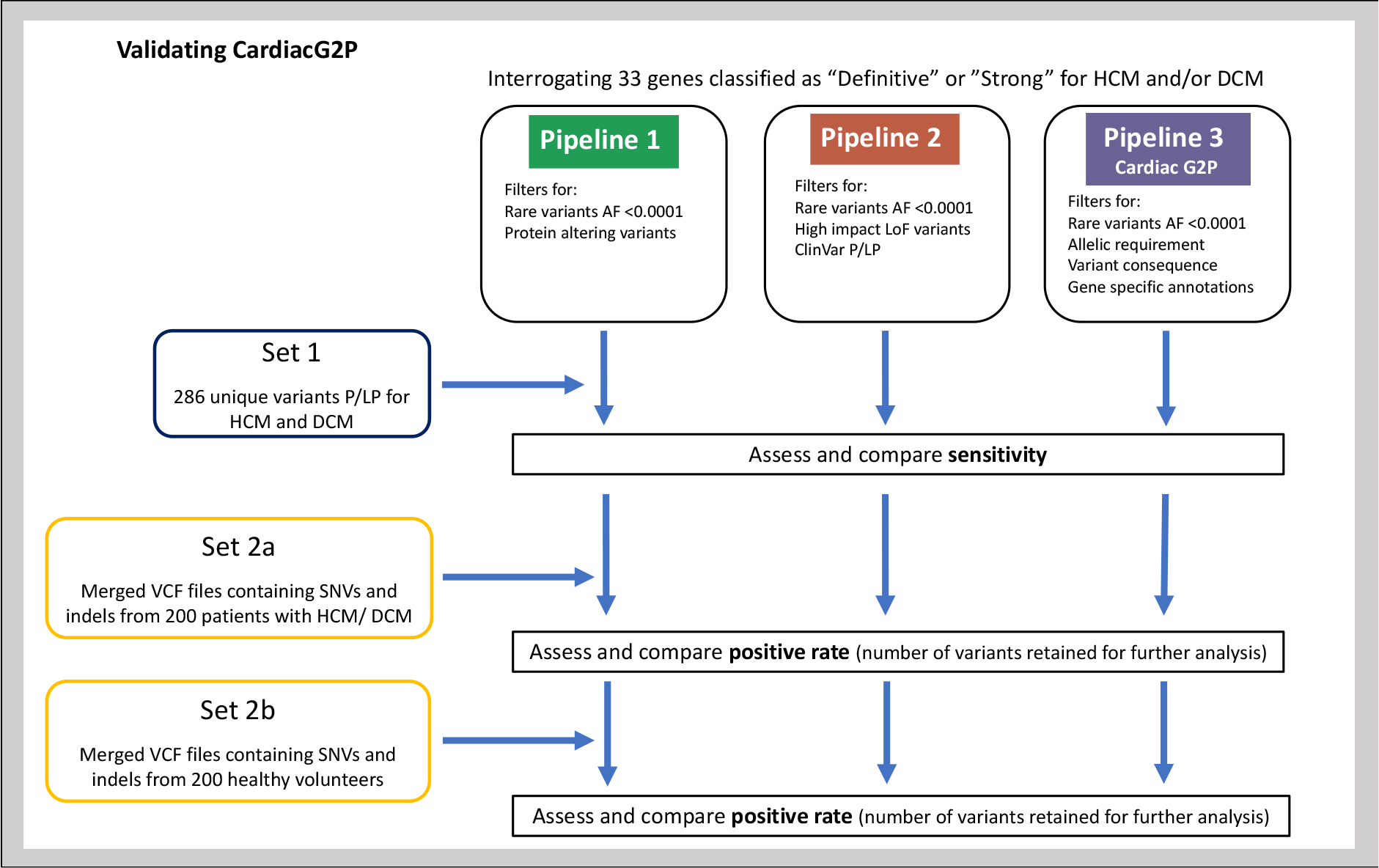
Validating CardiacG2P. Two generic variant prioritisation pipelines (pipeline 1 &2) were compared to CardiacG2P (pipeline 3). All 3 pipelines interrogate the same gene list which includes 21 HCM and 12 DCM genes. ***Pipeline 1****: filtered rare (gnomAD global allele frequency (AF) <0.0001) AND protein altering variants*. ***Pipeline 2****: filtered rare (AF <0.0001) AND ((high impact variants (e.g. stop gained, frameshift) AND high confidence by LOFTEE (VEP plugin) LoF variants) OR ClinVar P/LP variants)*. ***CardiacG2P (pipeline 3****): filtered rare variants (AF <0.0001) and incorporates allelic requirement, variant consequence and gene specific annotations of a restricted repertoire of pathogenic alleles appropriate for the disease under interrogation – e.g. restricted variant classes, specific variants, or restricted regions of the protein*. ***Set 1****: contains 286 unique variants identified and classified as P/LP for HCM or DCM by a specialist NHS cardiovascular genetics lab. A VCF file with these variants was created, annotated by VEP and filtered according to the 3 pipelines. Sensitivity (number of P/LP variants retained) was assessed*. ***Set 2a****: is a merged VCF file with SNVs and indels from 200 patients with HCM or DCM*. ***Set2b****: is a merged VCF file with SNVs and indels from 200 healthy volunteers*. *Set2a and 2b were separately annotated by VEP and filtered according to the 3 pipelines. Positive rate (the number of variants retained for further analysis) was assessed*. *AF – allele frequency, DCM – dilated cardiomyopathy, HCM – hypertrophic cardiomyopathy, indels – insertion or deletion variants, LoF – loss of function, P/LP – pathogenic/likely pathogenic, SNVs – single nucleotide variants, VCF – variant call format, VEP – variant effect predictor*

To compare these different approaches, two test sets of data were generated (see Figure 2).

### Set 1: to assess sensitivity

Set 1 contains 286 unique gold-standard true positive variants classified as P/LP for HCM and DCM in the last 3 years by the Clinical Genetics & Genomics Laboratory of the NHS Genomic Medicine Service South East Genomics Laboratory Hub at the Royal Brompton Hospital, London, which is one of 4 NHS England specialist cardiovascular genetics labs. These variants were identified using a custom gene panel using Agilent SureSelect QXT library preparation sequenced on Illumina MiSeq or NextSeq platforms. All variants were evaluated following guidelines produced by the ACMG/AMP(19) and the Association for Clinical Genomic Science (ACGS)(20)using an in-house validated pipeline.

For this study, a variant call format (VCF) file was created using these variants, and then annotated using VEP(9) version 104 and filtered according to the 3 pipelines. We compared the number of P/LP variants retained by each of the 3 methods.

### Set 2: to assess the positive rate - the number of variants retained for further analysis

Set 2a contains data from 200 patients with cardiomyopathy (either HCM or DCM) from the Royal Brompton & Harefield Hospitals Cardiovascular Research Biobank. Set 2b contains data from 200 healthy volunteers recruited for the digital heart project(21). Participants provided written informed consent, and research had ethics committee approval. No individual patient data is reported. The GRCh37 reference genome assembly was used for sequencing and analysis. Details of the sequencing panels and platforms and the bioinformatics pipelines used for variant calling are previously reported(22). Briefly, samples were sequenced using the Illumina TruSight Cardio Sequencing Kit, which includes 174 genes reported as associated with ICCs, on the Illumina MiSeq and NextSeq platforms. Targeted DNA libraries were prepared according to manufacturers’ protocols before performing paired-end sequencing. For this study, merged VCF files containing single nucleotide variants (SNVs), and insertion or deletion variants were annotated using VEP version 104 and filtered according to the 3 pipelines described above.

Since it is not possible to define a gold-standard classification for these variants that does not incorporate the same expert knowledge captured in CardiacG2P (except potentially for a very small number of variants with orthogonal segregation data), we report the total number of variants retained by each of the three methods (the positive rate), rather than positive predictive value. This is indicative of the analytical burden for a diagnostic laboratory manually interpreting variants of interest retained by a filtering pipeline. We have included a healthy cohort to represent the potential analytical burden of secondary findings.

## Results

### Inheritance and disease-associated variant consequences in established ICC genes

Forty cardiomyopathy gene-disease pairs (22 for HCM, 12 for DCM and 6 for ARVC; overall 33 unique genes) were analysed for inheritance pattern, allelic requirement, disease-associated variant consequences, and variant classes reported with evidence of pathogenicity. These are presented in Table 1 (typical HCM, DCM and ARVC) and Additional File 1 (syndromic disorders that include HCM where LVH may be a presenting feature). Twenty-five channelopathy gene-disease pairs (11 for LQTS, 1 for BrS, 8 for CPVT and 5 for SQTS; overall 15 unique genes) are presented in Table 2. Narrative summaries accompany each gene-disease pair, with content including relevant transcripts, specific pathogenic variants, mutational hotspots, phenotype notes and other important information raised during the expert panel reviews and discussion (see Additional File 3 or Additional File 4).

### Cardiomyopathy

Cardiomyopathy genes are predominately characterised by autosomal dominant inheritance with incomplete penetrance. However, 3/6 ARVC genes demonstrate both autosomal dominant and recessive inheritance; *JUP*-related Naxos disease (a syndrome characterised by ARVC, woolly hair and palmoplantar keratoderma) is exclusively inherited in an autosomal recessive manner, and 3/14 syndromic HCM genes (*FHL1, GLA* and *LAMP2*) are X-linked.

Importantly, only one of the eight core sarcomere-encoding HCM-associated genes (*MYBPC3*) causes disease through haploinsufficiency. LoF is not an established mechanism for the other 7 core HCM genes (as listed in Table 1) and NMD-competent PTCs are not known to be pathogenic. Instead, missense variants and variants predicted to escape NMD leading to an altered gene product sequence rather than decreased gene product level should be prioritised. This is also the case for 8/14 syndromic HCM gene-disease pairs (*CACNA1C, FLNC, PRKAG2, PTPN11* (Noonan), *PTPN11* (Noonan syndrome with multiple lentigines), *RAF1, RIT1, TTR*), 3/12 DCM (*DES, TNNC1* and *TNNT2*) and 2/6 ARVC (*JUP, TMEM43*).

Additional useful information for variant filtering is captured in individual narrative summaries. For example, for *TTN*-related DCM, only PTCs that are in exons constitutively expressed in both major adult cardiac isoforms (PSI > 0.9) should be prioritised (21,23,24).

Very few pathogenic missense variants in *TTN-*related DCM have been identified: to our knowledge there are only three reported with segregation evidence(25–27). Individually rare missense variants in *TTN* are collectively extremely common in the population (>50%, depending on allele frequency cut-off), and there are seldom established approaches to prioritise these in the absence of an informative pedigree. There are instances where evidence for disease comes primarily from one variant class such as missense variants only in *MYL2, MYL3* and *TPM1*-related HCM, or from a single well-characterised variant, such as *TMEM43*-related ARVC and the founder missense variant NM_024334.3(TMEM43) c.1073C>T (p.S358L)(28). Pathogenicity of other variant classes, or indeed other missense variants, for *TMEM43* is not established and this should guide interpretation of variants in these gene-disease relationships.

For some gene-disease relationships, there are gene regions where there is a high confidence for pathogenicity, for example exon 9 in *RBM20* related DCM (RS motif, amino acids 634-638). Other examples of mutational hotspots are referenced in individual curations.

### Channelopathy

The channelopathy genes are predominately characterised by autosomal dominant inheritance, though 7/25 gene-disease pairs demonstrate autosomal recessive inheritance. For 7/11 LQTS, 4/7 CPVT and 5/5 SQTS, disease is due to altered gene product sequence and not a decrease in gene product level. For these gene-disease relationships, it is missense variants and other non-truncating variants that should be prioritised and assessed for pathogenicity.

Many of the channelopathy genes are implicated in more than one phenotype, or overlapping phenotypes; 25 gene-disease relationships are evaluated here but only 15 unique genes. Importantly, for several genes distinct variant classes drive different phenotypes through distinct mechanisms. As an example, both PTCs and missense variants leading to LoF of *KCNQ1* are associated with LQTS and Jervell Lange-Nielsen syndrome. In contrast, almost all evidence for *KCNQ1* as a cause of SQTS is derived from a single missense variant (p.V141M) and functional studies in cell models have confirmed GoF as the mechanism (29,30). Similarly, both PTCs and non-truncating variants leading to LoF of *SCN5A* are associated with BrS, whereas *SCN5A*-related LQTS is caused by pathogenic missense variants and inframe indels leading to GoF.

For certain gene-disease pairs, there are gene regions where there is a higher confidence for pathogenicity such as, for non-truncating variants, the transmembrane regions and C-terminus domains for *KCNQ1*-related LQTS (31,32), and the ion channel transmembrane regions and specific N-terminus and C-terminus domains for *KCNH2*-related LQTS(32). There are other examples of mutational hotspots referenced in individual curations (see Additional File 3 or Additional File 4) .

### CardiacG2P reduces the number of variants prioritised, without compromising sensitivity to detect true positives

#### 1. Assessing sensitivity

We assessed variant filtering using the CardiacG2P dataset for the identification of known P/LP variants previously classified by the cardiovascular laboratory of the NHS Genomic Medicine Service South East Genomics Laboratory Hub at the Royal Brompton Hospital, London. A total of 286 P/LP variants in 16 HCM/DCM genes were used to assess the performance of CardiacG2P compared to two other generic pipelines (see Figure 3A). CardiacG2P correctly identified 282/286 variants, a sensitivity of 98.6%. This was superior to both alternative approaches (pipeline 1 272/286, sensitivity 95.1%, P_fisher_=0.028; pipeline 2 199/286, 69.6%, P_fisher_=1.8x10^-24^). Four variants were not retained by CardiacG2P. These comprised 1 *TTN* missense variant and 2 intronic and 1 synonymous variant in *LMNA*. All four of these variants were classified as P/LP by the clinical laboratory due to impacts on splicing, so the limited sensitivity is due to an incomplete upstream annotation of the variant consequence, rather than an “error” in downstream filtering.

**Figure 3:**
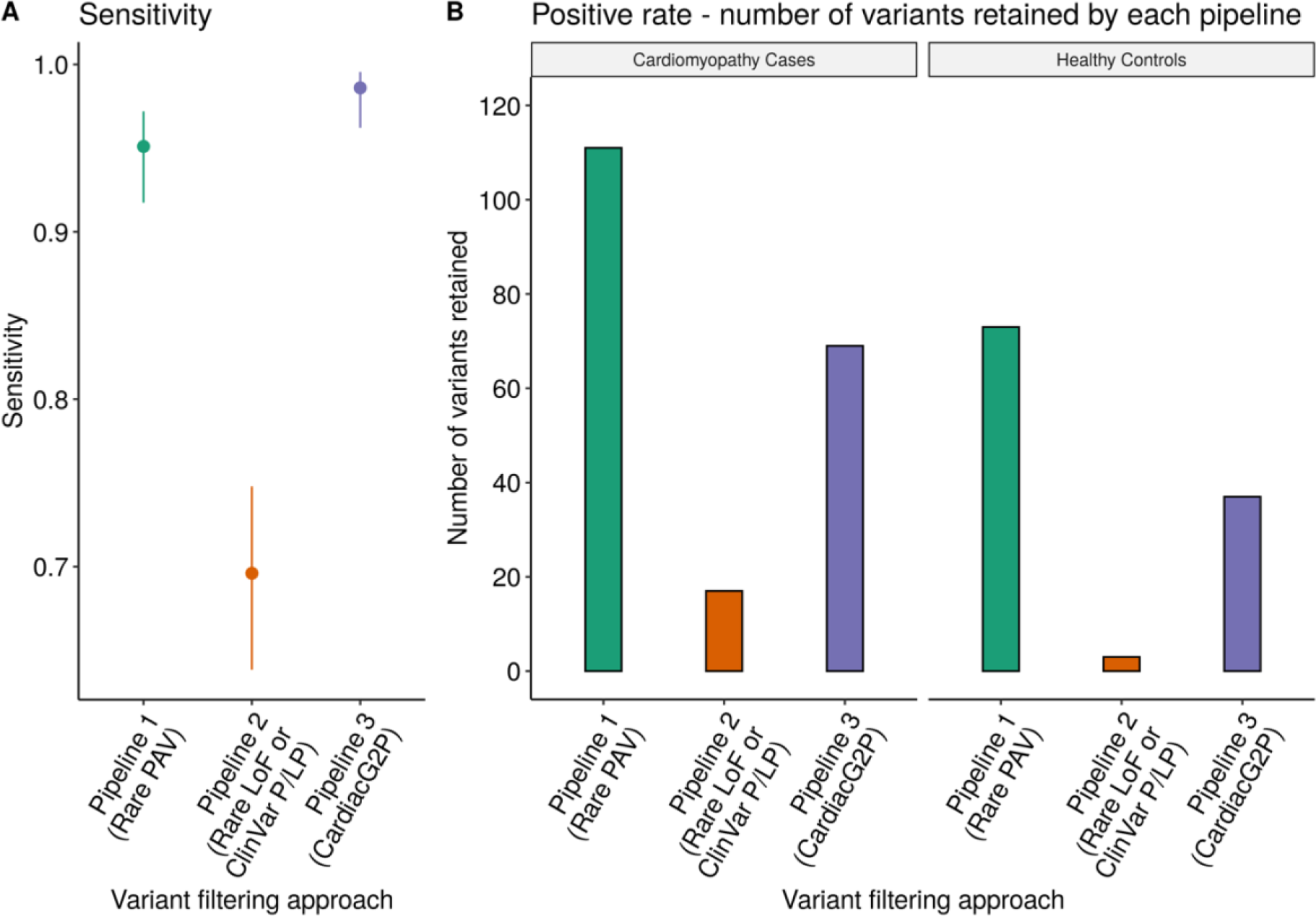
A variant prioritisation approach that incorporates structured data representing disease mechanisms and allelic requirement for specific gene-disease pairs (CardiacG2P) outperforms other scalable variant-prioritisation approaches. *A – comparison of the sensitivity of 3 variant filtering approaches to prioritise 286 variants classified as pathogenic/likely pathogenic (P/LP) for hypertrophic cardiomyopathy (HCM) and dilated cardiomyopathy (DCM). Error bars = 95% confidence intervals (CI). Pipeline 1 (green), prioritises all rare protein altering variants (PAV), sensitivity 0.95, 95% CI [0.92, 0.97]; Pipeline 2 (red), prioritises all rare loss of function (LoF) variants, and those classified as P/LP by ClinVar, sensitivity 0.70, 95% CI [0.64, 0.75]; Pipeline 3 (purple), prioritises variant classes according to specific characteristics of each gene-disease pair (CardiacG2P), sensitivity 0.99, 95% CI [0.96, 1.0]. CardiacG2P has a higher sensitivity when compared to Pipeline 1, P^fisher^ = 0.028 and Pipeline 2, P^fisher^ = 1.8x10^-24^*. *B – the positive rate (number of variants retained) by 3 variant filtering approaches for a cardiomyopathy cases (left panel), using a dataset of 5681 unique variants from 200 individuals with confirmed HCM/DCM, and healthy controls (right panel), using a dataset of 6060 unique variants from 200 healthy individuals. Pipeline 1 (green), filtering for rare PAV; Pipeline 2 (red), filtering for rare LoF variants or those classified as P/LP by ClinVar; Pipeline 3 (purple), filtering using CardiacG2P. CardiacG2P demonstrated more efficient variant prioritisation compared to Pipeline 1 in both the disease cohort (P^fisher^ = 0.002) and healthy controls (P^fisher^ = <0.0001)*.

#### 2. Assessing variant prioritisation – the number of variants retained for further analysis

We compared the number of variants retained by the 3 pipelines filters to assess the positive rate of each approach (see Figure 3B). A pipeline with a high positive rate requires more downstream human effort for final variant adjudication.

First, we compared sequencing data (5681 unique variants) from 200 individuals with a confirmed diagnosis of HCM or DCM. CardiacG2P prioritised 69 variants, pipeline 1 prioritised 111 variants, and pipeline 2 prioritised 17.

Since the cardiomyopathy cohort would be very substantially enriched for true positives, we also assessed the positive rate in a healthy cohort, indicative of variants that may require follow up during opportunistic screening for secondary findings. 6060 unique variants found in 200 healthy volunteers were analysed by each pipeline, with CardiacG2P prioritising 37 variants, pipeline 1 prioritising 73 variants, and pipeline 2 prioritising 3 variants.

Pipeline 2 prioritises fewest variants in both contexts (17/5681 and 3/6060 respectively). This is to be expected as it filters on only high impact LoF variants or variants classified as P/LP by ClinVar. However, this method also demonstrated the lowest sensitivity for P/LP variants (69.6%), because LoF is not a known mechanism for many of the ICC genes and any pathogenic missense or other non-truncating variants will be wrongly discarded by this method. In the disease cohort, compared to pipeline 1 which retains all PAVs, CardiacG2P demonstrated more efficient variant prioritisation retaining significantly fewer variants, (P_fisher_ = 0.002). In the healthy cohort, where we would expect a higher number of false positive variants to be prioritised, CardiacG2P retained half the number of variants compared to pipeline 1 (37 vs. 73 variants, P_fisher_ = <0.0001). CardiacG2P also maintained the highest sensitivity of all 3 pipelines at 98.6%.

## Discussion

Accurate variant classification in ICC genes requires robust strength of a gene-disease relationship and knowledge of inheritance pattern, disease mechanism and pathogenic variant classes(33). The literature is constantly expanding with newly reported variants and re-evaluation of variant classifications happening frequently, presenting challenges with updating these classificaitons. In ClinVar alone there are over 1 million variants submitted.

Over 49,000 have conflicting interpretations and others are submitted under multiple phenotypes making the relevant disease for the variant classification unclear. Variant classification is expanding beyond laboratories with long standing interest and expertise in cardiovascular genetics. The ACMG secondary findings list means that others will need to rapidly acquire proficiency in reporting variants in CV genes. To aid this process, we have curated the mode of inheritance, allelic requirement, and disease associated variant consequences, for 65 ClinGen-curated ICC gene-disease pairs (48 unique genes) and, following review by multidisciplinary expert panels, present this information as a publicly available structured dataset both here and via CardiacG2P (https://www.ebi.ac.uk/gene2phenotype/downloads), to aid variant analysis. This dataset is compatible with the existing G2P plugin for the widely used Ensembl Variant Effect Predictor. Variant Effect Predictor.

Overall, for 36/65 gene-disease relationships, disease is due to altered gene product sequence, not a decrease in gene product level. Therefore, for over 50% of the ICC genes evaluated here, current data cautions against a default prioritisation of predicted protein-truncating variants as pathogenic, with LoF as a presumed mechanism. The majority of the ICC genes are characterised by autosomal dominant inheritance with incomplete penetrance, however there are notable examples of autosomal recessive and X-linked inheritance and more fully penetrant variants.

As well as the structured data, we have included narrative summaries to capture key notes that arose during evidence collection and expert discussion that may also aid variant filtering and interpretation. Throughout these discussions, several themes that relate to all the ICC genes emerged. It is widely accepted that ICC genes often display incomplete penetrance, however given that most penetrance estimates have been made using cases(34), expert opinion and emerging evidence agree that overall penetrance may be lower than previously reported. This is particularly relevant and should be considered when assessing patients who have a pathogenic variant identified as a secondary finding outside of families with known disease(34,35).

There are many examples of autosomal dominant ICC gene-disease relationships where compound heterozygous and homozygous variants, or variants in more than 1 known disease gene, are also reported. Approximately 10% of genotype positive LQTS patients have >1 pathogenic variant in >=1 LQTS related gene(36,37). There was debate amongst the expert panel on how this should be recorded. In those instances where phenotypic features of people with biallelic variants are truly different to those with monoallelic variants (e.g. Jervell Lange-Nielsen Syndrome), this may represent true autosomal recessive or digenic inheritance and should be recorded as such. However, it was recognised that for many of the ICC genes, disease severity and penetrance are often the main distinguishing features between monoallelic and biallelic disease. In this circumstance, autosomal dominant inheritance is recorded with further information in the narrative summary acknowledging that if a second P/LP variant is identified, the disease often appears to be more penetrant and more severe (38–41)and can even lead to neonatal lethality.

It is important to interpret variants in the context of a gene-disease relationship rather than in the gene alone(42). There are several ICC genes implicated in more than one phenotype. For some, distinct mechanisms drive different diseases, e.g. *MYH7*-related HCM and *MYH7*-related DCM. Although both are caused primarily by missense variants in *MYH7* altering the gene product structure, distinct alleles have opposing effects on sarcomere force generation and drive different phenotypes(43,44). In contrast, although *DSP* is also associated with multiple phenotypes (including DCM, DCM with cutaneous features, ARVC, and Carvajal syndrome), these are overlapping and it does not appear that distinct mechanisms drive different presentations. Similarly, although the phenotype most frequently shown by patients with *CALM* pathogenic variants is LQTS, others display CPVT and sudden unexplained death and some *CALM* variants have been associated with both LQTS and CPVT, without evidence of distinct mechanisms underlying different phenotypic manifestations(42,45).

Here we have evaluated CardiacG2P as a first-tier variant filter. This variant consequence and allelic requirement aware approach increases the efficiency of variant prioritisation, without compromising on sensitivity, in comparison to two generic bioinformatic filtering pipelines (see Figure 3). CardiacG2P retains significantly fewer variants than a pipeline where all PAVs are prioritised. The difference between CardiacG2P and the generic pipelines is even more marked in a healthy cohort, highlighting benefits in reducing the analytical burden of assessing secondary findings. CardiacG2P correctly identified 282/286 previously classified P/LP variants. The four variants that were not retained comprised 1 *TTN* missense variant and 2 intronic and 1 synonymous variant in *LMNA*. The 3 *LMNA* variants were all predicted to impact splicing with functional data available to support 2 of them. The *TTN* missense variant was classified as LP because it was also predicted to have a splicing effect and had been detected in 4 in-house DCM patients before. CardiacG2P filters are based on the consequence assigned by VEP, and upstream annotation by VEP had not recorded these 4 variants as impacting splicing. Improvements in prediction of variant consequence, especially for variants impacting splicing, will allow these to be retained. While our framework recognises that some intronic or coding variants can impact splicing, it is not an expected consequence for the vast majority of such variants and therefore these will not be routinely retained. Rarely there will be instances where pathogenic variants are filtered by G2P if the upstream consequence annotation is incomplete or incorrect, so we must caution against simply discarding all non-prioritised variants and must continue to improve tools for variant consequence annotation.

As our knowledge of genes and specific variants contributing to ICCs expands, it is possible to update the CardiacG2P dataset dynamically and subsequently include new information in the VEP G2Pplugin.

## Conclusions

As variant reporting moves away from labs with expertise in certain disease areas, it is vital that accurate variant classifications are maintained. Here, we present evidenced-based inheritance and variant consequence curations for robustly associated ICC genes with the benefit of expert review and opinion. We present this data for the first time in a structured format using new standardised terminology. This dataset is a publicly available resource, CardiacG2P, and we have demonstrated here its utility in the filtering of genomic variants in ICC genes.

## List of abbreviations

|  |  |
| --- | --- |
| ACGS | Association for Clinical Genomic Science |
| ACMG SF V3.1 | American College of Medical Genetics and Genomics Secondary Findings list |
| AD | Autosomal dominant |
| AR | Autosomal recessive |
| ARVC | Arrhythmogenic right ventricular cardiomyopathy |
| ATS | Andersen-Tawil Syndrome |
| BrS | Brugada syndrome |
| ClinGen | The Clinical Genome Resource |
| CPVT | Catecholaminergic polymorphic ventricular tachycardia |
| CV | cardiovascular |
| DCM | Dilated cardiomyopathy |
| ES | Exome sequencing |
| G2P | Gene2phenotype |
| GCEPs | Gene Curation Expert Panels |
| GenCC | Gene Curation Coalition |
| GoF | Gain of function |
| GS | Genome sequencing |
| HCM | Hypertrophic cardiomyopathy |
| HPO | Human phenotype ontology |
| IC | Intrinsic cardiomyopathy |
| ICCs | Inherited cardiac conditions |
| Indels | Insertions or deletions |
| JLNS | Jervell and Lange-Nielsen |
| LoF | Loss of function |
| LQTS | Long QT syndrome |
| LVH | Left ventricular hypertrophy |
| ND | Naxos disease |
| NMD | Nonsense mediated decay |
| P/LP | Pathogenic/likely pathogenic |
| PAVs | Protein altering variants |
| PSCD | Primary systemic carnitine deficiency |
| PSI | Percent spliced in |
| PTCs | Premature termination codons |
| SNVs | Single nucleotide variants |
| SO | Sequence ontology |
| SQTS | Short QT syndrome |
| TS | Timothy syndrome |
| VCF | Variant call format |
| VEP | Variant Effect Predictor |

## Declarations

### Ethics approval and consent to participate

No individual patient data is reported.

Royal Brompton & Harefield Hospitals Cardiovascular Research Biobank participants provided written informed consent, HRA research ethics approval: South Central Hampshire B Research Ethics Committee 19/SC/0257. Healthy volunteers in the digital heart project provided written informed consent, HRA research ethics committee approval: London – West London and GTAC Research Ethics Committee 09/H0707/69.

### Consent for publication

Not applicable

### Availability of data and materials

All data generated during this study are included in this published article. A structured representation of the data is available in the Additional Materials, and also through G2P (https://www.ebi.ac.uk/gene2phenotype/downloads), which is also searchable through the GenCC portal (https://thegencc.org/).

### Competing interests

EMM is a Consultant for Amgen, AstraZeneca, Avidity Biosciences, Cytokinetics, PepGen, Pfizer, Stealth Biotherapeutics, Tenaya Therapeutics, and founder of Ikaika Therapeutics. CJ is a Consultant for Pfizer Inc (paid), StrideBio Inc (unpaid), and Tenaya Inc (unpaid). TL has research grant support from Pfizer. DPJ is a Consultant for Alexion, Alleviant, Cytokinetics, Novo Nordisk, Pfizer, and Tenaya Therapeutics. JI has research grant support from Bristol Myers Squibb. J.S.W. has received research support or consultancy fees from Myokardia, Bristol-Myers Squibb, Pfizer, and Foresite Labs. No other authors report competing interests.

### Funding

J.S.W was supported by the Sir Jules Thorn Trust [21JTA], Wellcome Trust [107469/Z/15/Z], Medical Research Council (UK), British Heart Foundation [RE/18/4/34215]; NHLI Foundation Royston Centre for Cardiomyopathy Research, and the NIHR Imperial College Biomedical Research Centre. KSJ was supported by the Wellcome Trust [222883/Z/21/Z]. AMR was supported by the British Heart Foundation Fellowship [FS/CRLF/21/23011]. PT was supported by the Wellcome Trust [200990/A/16/Z].This publication was supported in part by the National Human Genome Research Institute of the National Institutes of Health through the following grants: U24HG009650. AW and PvT are supported by CVON/Dutch Heart Foundation PREDICT2 (2018–30); RT is supported by the Canada Research Chairs program; TL receives support from BHF Research Accelerator; BC is a Senior Clinical Investigator of the Research Foundation – Flanders; EMM is supported by NIH HL128075, American Heart Association.

For the purpose of open access, the authors have applied a CC BY public copyright licence to any Author Accepted Manuscript version arising from this submission.

The views expressed in this work are those of the authors and not necessarily those of the funders.

### Authors’ contributions

JSW, AMR, KSJ conceived the work. RW, PJO, ME, AF, MG, KSJ curated gene-disease pairs and presented these to the expert panels for review. EO, APL, EMM, C.AAC, LB, OJ, BC, CJ, SP, TL, ME, DPJ, PvT, EJ, BA, REH, NK, CT, JI reviewed cardiomyopathy curations; JR, MC, WZ, AA, ACS, RT, VN, AW, MG reviewed channelopathy curations; JSW, PT, KSJ conceived the design for validating CardiacG2P experiment; PT performed the analysis for validating CardiacG2P; Manuscript written by KSJ, JSW, AMR; all authors read and approved the final manuscript.

## Data Availability

All data generated during this study are included in this published article. A structured representation of the data is available in the Additional Materials, and also through G2P (https://www.ebi.ac.uk/gene2phenotype/downloads), which is also searchable through the GenCC portal (https://thegencc.org/

https://thegencc.org

## Acknowledgements

The following authors have taken part in the ClinGen Cardiovascular Clinical Domain Working Group https://clinicalgenome.org/working-groups/clinical-domain/cardiovascular/ and/or are members of a ClinGen Gene Curation Expert Panel (GCEP) affiliated to this working group: Roddy Walsh, Matthew Edwards, Courtney Thaxton, Melanie Care, Wojciech Zareba, Arnon Adler, Amy C Sturm, Valeria Novelli, Marco Perez, Emma Owens, Iftikhar Kullo, Lucas Bronicki, Olga Jarinova, Bert Callewaert, Stacey Peters, Thomas Lumbers, Megan Mayers, Elizabeth Jordan, Babken Asatryan, Neesha Krishnan, Nadia Qureshi, Ray E Hershberger, Anwar A. Chahal, Andrew Landstrom, Cynthia James, Elizabeth M McNally, Daniel Judge, Peter van Tintelen, Arthur Wilde, Michael H. Gollob, Jodie Ingles, and James S Ware.

## List of Additional materials

Additional File 1:

File format - xls

Title *- Structured representation of data from curation of syndromic forms of (hypertrophic) cardiomyopathy gene-disease pairs*.

Description *– A table showing* the curation of syndromic forms of (hypertrophic) cardiomyopathy that can have isolated left ventricular hypertrophy as the presenting feature: structured representation of inheritance, allelic requirement, disease-associated variant consequence, and variant classes reported with evidence of pathogenicity for each gene-disease pair

Additional File 2:

File format – docx

Title - *Standard operating procedure for gene-disease curations*

Description - This document provides a template and standard operating procedure for the curation of inheritance, allelic requirement and disease mechanism for gene-disease pairs already curated by ClinGen using standardised terminology.

Additional File 3:

File format – docx

Title – *Inheritance and mechanism curation summaries for all gene-disease pairs*

Description – Data from individual gene-disease pair curations presented in individual tables with a narrative summary describing key messages from the expert review with relevant publication identifiers.

Additional File 4:

File format – xls

Title – *CardiacG2P dataset*

Description – The same information that is presented in Additional File 3 is included here in xls format. Sheet 1 (CardiacG2P_curated) includes a structured representation of inheritance and mechanism data for all curated gene-disease pairs. Inaddition this also includes information for 7 genes related to a syndrome where LVH is seen only with overt syndromic features. Sheet 2 (Narrative_summaries) has narrative summaries for each gene-disease pair as plain free text. Sheet 3 (Other_limited) is a list of gene-disease pairs where there is no established relationship (gene disease validity assertion from ClinGen); these are included for completeness.

## Notes

### Author Declarations

No individual patient data is reported. Royal Brompton & Harefield Hospitals Cardiovascular Research Biobank participants provided written informed consent, HRA research ethics approval: South Central Hampshire B Research Ethics Committee 19/SC/0257. Healthy volunteers in the digital heart project provided written informed consent, HRA research ethics committee approval: London West London and GTAC Research Ethics Committee 09/H0707/69.

