## Additional File 2 for "Beyond gene-disease validity: capturing structured data on inheritance, allelic-requirement, disease-relevant variant classes, and disease mechanism for inherited cardiac conditions"

**Using a framework of standardised terminologies to define inheritance, allelic requirement, disease-associated variant classes, and disease-associated variant consequence for gene-disease pairs.**

This document provides a template and standard operating procedure for the curation of inheritance, allelic requirement and disease mechanism for gene-disease pairs already curated by ClinGen using standardised terminology.

**TEMPLATE**

**ClinGen**:

Include summary of evidence to support gene-disease relationship. This can be found at

<https://clinicalgenome.org/> and searching for the specific gene.

Paste web link for ClinGen evidence summary page here e.g. for KCNQ1 http://search.clinicalgenome.org/kb/genes/HGNC:6294.

If the summary page is not on the ClinGen website, these can sometimes be found in the Supplementary data of the ClinGen curation paper.

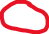

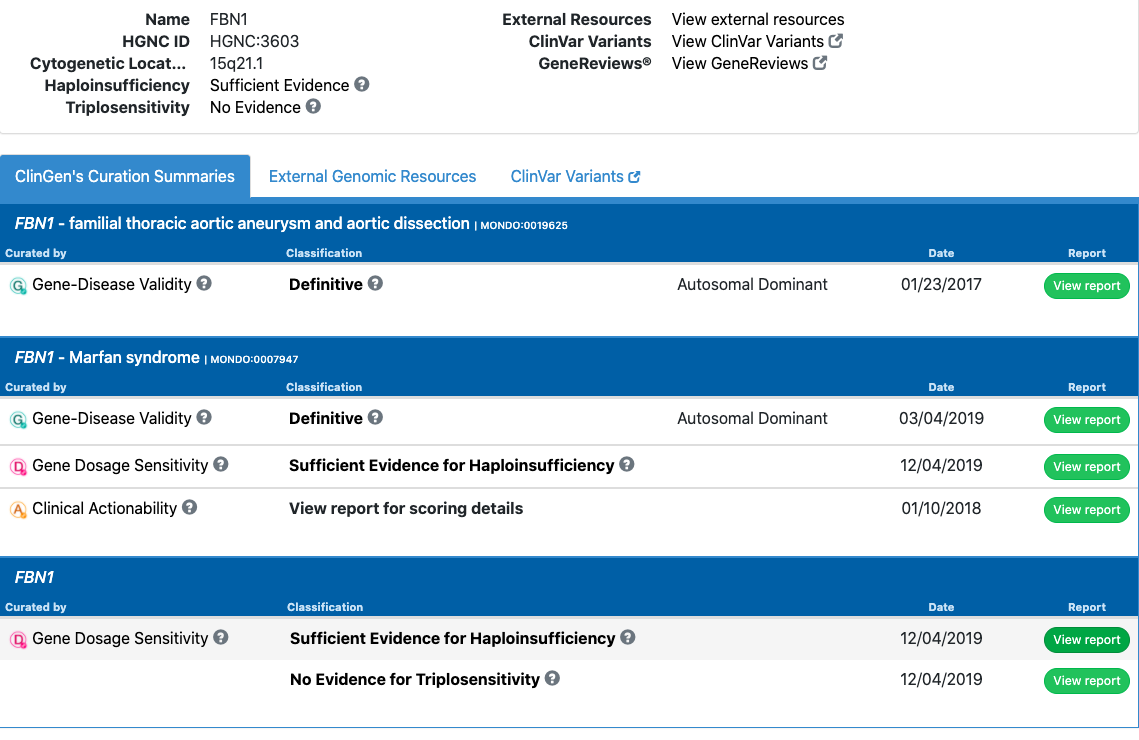

**Review of source material:**

Review and include the reference (PMID) for the relevant ClinGen gene-disease validity paper e.g. for hypertrophic cardiomyopathy PMID: 30681346. This is likely to include useful summary information and publications to refer to.

**Other Literature review:**

This is to gather new information, not to re-evaluate the gene-disease relationship. Evidence is collected primarily from published peer-reviewed literature, but can also be present in publicly accessible resources, such as variant databases. Up to date reviews from centres with particular expertise in a given gene or disease are particularly helpful.

Useful publication search engines include:

PubMed

Google Scholar

LitVar

GeneCards

Mastermind

Other useful information

GeneReviews and the “Molecular Genetics” section

Omim

ClinVar to search for relevant variant classes

PanelApp (If using a resource like PanelApp need to reference the assertion and check original references)

As these gene-disease pairs have all been classified as “Definitive” or “Strong” by ClinGen, they are usually well established and there may be abundant information. The goal is not to re-evaluate the gene-disease validity, and the literature review therefore does not have to be exhaustive. The literature search should be focused on establishing inheritance pattern, allelic requirement and where possible disease-associated variant class and functional consequences.

For example, for some gene-disease pairs it may be well established that the pattern of inheritance is autosomal dominant but there may be a small number of reports of recessive inheritance. A broad search of the literature can determine if other modes of inheritance have been reported, using search terms:

**Gene AND disease AND (“recessive” OR “autosomal recessive” OR “homozyg*” OR “compound heterozyg*” OR “biallelic”)**

**Gene AND disease AND (dominant OR “autosomal dominant” OR monoallelic OR heterozyg*)**

**Gene AND disease AND (“x-linked” OR “x linked” OR “X chromosome” OR “X linked dominant” OR “X linked recessive”)**

Any reports of a different mode of inheritance should be reviewed to see if they are relevant or not. For many cardiac genes, a second hit may lead to a more severe phenotype but that does not necessarily mean the inheritance follows a recessive or digenic pattern as both/either of the first and second hit would in fact cause disease in isolation.

For disease mechanism, literature review should focus on establishing the most likely functional consequences and the main variant classes associated. Curators should review the evidence for haploinsufficiency in the ClinGen Dosage Sensitivity curation (<http://search.clinicalgenome.org/kb/gene-dosage?page=1&size=25&order=asc&sort=symbol&search=>), pathogenic/likely pathogenic variant classes on ClinVar (Simple ClinVar can be a helpful tool to search ClinVar, see screen shot below and link <http://simple-clinvar.broadinstitute.org>) and other public variant databases where available. For well described genes, recent publications re-evaluating variants, expert reviews, meta analyses and reviews of burden testing are highly relevant.

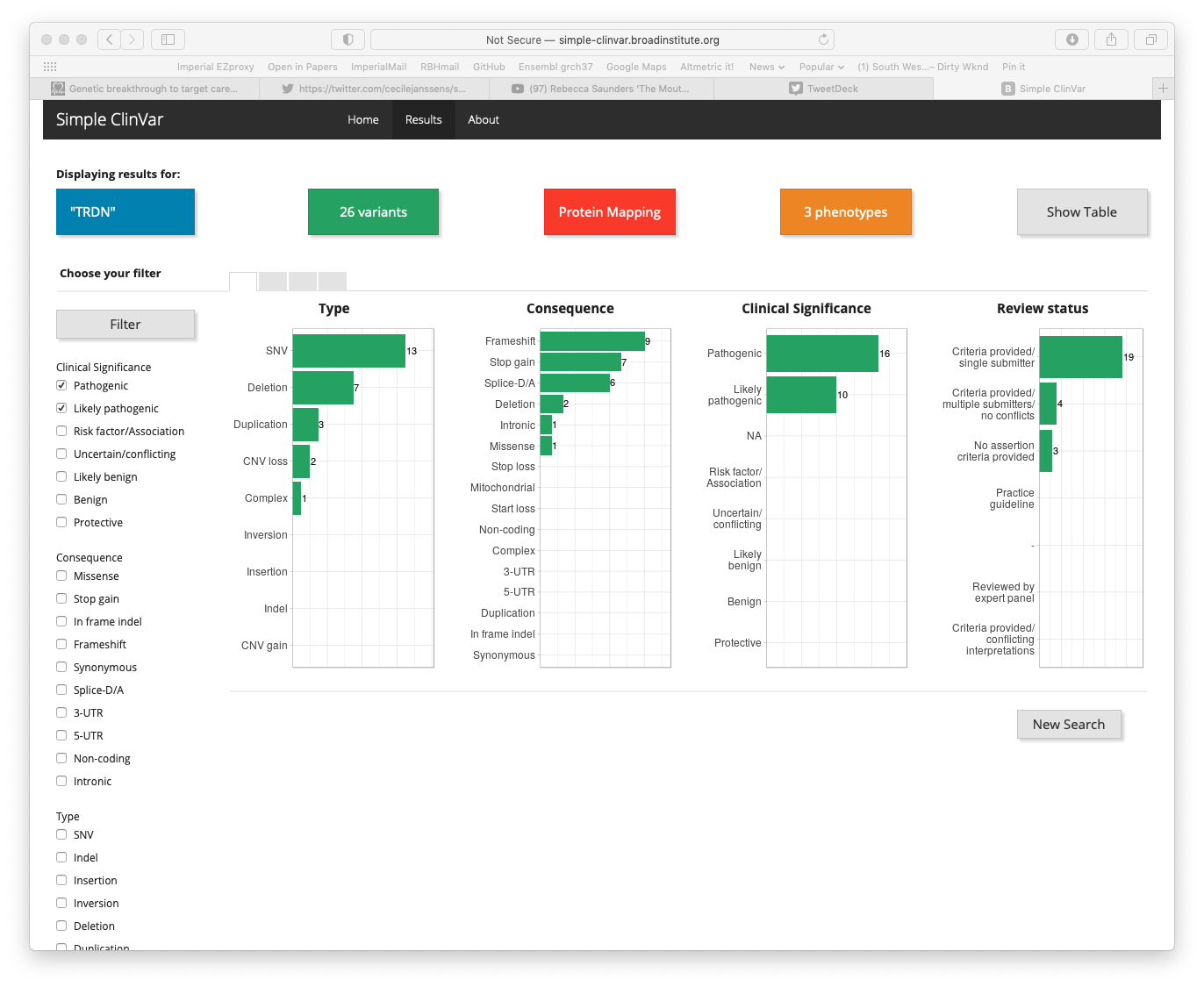

It is not necessary to review every variant. However if for example the predominant class of variant is missense but there are a small number of nonsense mutations reported, extra time should be spent determining whether there is sufficient evidence to include these as a pathogenic variant class before expanding the disease mechanism. Sufficient evidence could include segregation or functional evidence.

If high level reviews are not available for a gene-disease pair, for example a rare syndromic disease pairing, then a broad literature search may be necessary e.g. Gene AND disease AND (variant OR mutation). For a variant class to be included that would add to the predicted functional consequence, there should be sufficient qualitative evidence to support that such as segregation, functional or burden data.

Where there is uncertainty that cannot be resolved a note should be made in the narrative summary.

Include PMIDs where possible and or links to other resources.

**Inheritance and Allelic Requirement**

High level terms, inheritance modifiers and cross cutting modifiers:

| **allelic requirement term** | **inheritance term** | **HPO id** |
| --- | --- | --- |
| monoallelic_autosomal | Autosomal Dominant | HP:0000006 |
| biallelic_autosomal | Autosomal Recessive | HP:0000007 |
| monoallelic_X_heterozygous | X-linked Dominant | HP:0001423 |
| monoallelic_X_hemizygous | X-linked Recessive | HP:0001419 |
| monoallelic_Y_hemizygous | Y-linked | HP:0001450 |
| mitochondrial | Mitochondrial | HP:0001427 |
| monoallelic_PAR | PAR dominant | HP:0034340 |
| biallelic_PAR | PAR recessive | HP:0034341 |

**Cross cutting Inheritance Modifiers HP:0034335**

| **Inheritance Modifier**  **HP:0034335** | **Definition** |
| --- | --- |
| Typically mosaic | Description of conditions in which, for example, constitutive mutation is lethal and cases are exclusively or predominantly mosaic. A much lower variant allele fraction (VAF) cutoff would be needed in analysis pipelines. |
| Typically de novo  HP:0025352 | Description of conditions that are exclusively or predominantly observed to display de novo variants. In some cases, this may be due to the limited reproductive fitness of affected individuals. |
| Typified by incomplete penetrance  HP:0003829 | description of conditions in which only a limited proportion of individuals with a given genotype exhibit the disease regardless of age assuming a full lifespan of 80 years. For example, Van der Woude syndrome due to IRF6 causes cleft lip and/or palate with incomplete penetrance estimated at 80%, and *C9orf72* causes frontotemporal dementia and/or amyotrophic lateral sclerosis with approximately 50% penetrance. |
| Typified by complete penetrance  HP:0003829 | Description of conditions in which all individuals with a given genotype exhibit the disease within a full lifespan of 80 years. For example, penetrance of Neurofibromatosis type 1 due to NF1 is close to 100%. |
| Variable age of onset | Description of conditions in which age of onset is highly variable and in which manifestation of the disease phenotype is not dependent on the age of the subject. |
| Typified by age-related onset  HP:0003831 | Description of conditions in which age of onset is typically not congenital and in which manifestation of the disease phenotype is dependent on the age of the subject. |
| Congenital onset | Description of conditions which are manifest at or before birth, for example cleft lip or talipes. |
| Imprinted  HP:0034338 | Requires that the abnormal allele be paternal or maternal in origin. |
| Displays anticipation  HP:0003743 | a phenomenon in which the severity of a disorder increases, or the age of onset decreases, as the disorder is passed from one generation to the next, typically due to expansion of a repeat sequence. For example, Myotonic Dystrophy is caused by triplet repeat expansion in the *DMPK* gene. |
| Requires heterozygosity  HP:0034343 | covers rare instances of a condition that is most severe in the heterozygous state. Such disorders are rare and currently all are X-linked. Most X-linked recessive conditions manifest if hemizygous in males, or biallelic in females, though may have a mild phenotype in the heterozygous state in females. However, Craniofrontonasal dysplasia due to *EFNB1* and *PCDH19*-related epilepsy are both X-linked recessive and paradoxically more severe in females. Hemizygous males may be mildly affected but seldom manifest the full phenotype. Importantly the mutant allele can be inherited from a normal or very mildly affected father. The mechanism is currently accepted to be due to cellular interference whereby the two distinct cell populations (those with and without the variant) exhibit abnormal cellular interactions in the mosaic state - in women, who are functionally mosaic due to random X inactivation, or mosaic males. The same mechanism could theoretically be applicable to autosomal genes with a mosaic variant. |
| Sex-limited expression  HP:0001470 | Condition in which the phenotype only manifests in one sex, ie either manifests in males or females but not both. Example: Autosomal recessive sex reversal due to DHH on chr12 manifests only in XY males causing gonadal dysgenesis, while XX females are phenotypically normal. |
| Contiguous gene syndrome  HP:0001466 | Syndrome caused by the effects of abnormality (typically a deletion or duplication) of 2 or more adjacent genes. |

For Info:

**Mitochondrial -** the inheritance of a trait encoded in the mitochondrial genome. Persons with mitochondrial disease may be male or female but the mode of inheritance is strictly maternal. No male with the disease can transmit it to their offspring.

**PAR** - genes within the pseudoautosomal regions (PAR) are inherited like autosomal genes. PAR1 comprises 2.6mb of the short-arm of both X and Y chromosomes in humans. PAR2 is at the tip of the long arms, spanning 320kb.

Normal male mammals have two copies of these genes: one in the pseudoautosomal region of their Y chromosome, the other in the corresponding portion of their X chromosome. Normal females also possess two copies of pseudoautosomal genes, as each of their two X chromosomes contains a pseudoautosomal region. Crossing over between the X and Y chromosomes is normally restricted to the pseudoautosomal regions; thus, pseudoautosomal genes exhibit an autosomal, rather than sex-linked, pattern of inheritance. So, females can inherit an allele originally present on the Y chromosome of their father.

**Notes:**

- For monoallelic_X_heterozygous (**X-linked dominant)** conditions, we would understand that those diseases manifest when het or hem (or indeed hom/compound het - though this may be more severe or lethal).
- For monoallelic_X_hemizygous (**X-linked recessive)** conditions, we would understand that these would not manifest when heterozygous (though they can manifest with ameliorated phenotype, or manifest if skewed inactivation etc - primarily recessive with milder female expression)

Terms are specific to each disease-gene pair, so for example if there is good evidence for manifesting carriers of X_Hemizygous disorders presenting in infancy/early childhood that would be coded as X_Hem in DDG2P.  If carriers only have late onset cardiomyopathy (eg female carrier of DMD) they would be X_het in the Cardiac panel but not DD.

- **maternal/paternal imprinting:** *involving a gene that is imprinted with either paternal or maternal silencing* Could be used when the gene-disease pair requires that the abnormal allele be paternal or maternal in origin.

Example: Angelman syndrome is caused by disruption of maternally imprinted *UBE3A* allele

Use of **cross cutting modifiers** enables recording of data important to reproductive advice and family screening.

- **Typically mosaic -** conditions which are exclusively or predominantly mosaic, where a constitutive mutation is lethal.

*Mosaic - the presence of a pathogenic genetic variant in only a distinct population of cells in a given organism.*

Some diseases are characterised by mosaicism, since a constitutive variant is lethal and is not seen in the population. Example: Proteus syndrome caused by pathogenic mosaic heterozygous variants in *AKT1*. It is hypothesised that pathogenic germline *AKT1* variants would be lethal early in development. This modifier should not be used to describe the situation of mosaicism in a parent.

- **Typically de novo -** conditions that are exclusively or predominantly de novo due to the limited reproductive fitness of affected individuals.

*De Novo - a genetic variant that has arisen in the fertilised egg during embryogenesis and has not been inherited from either parent.* Example: Cardiofaciocutaneous syndrome caused by pathogenic variants in *BRAF*. Many of the other rasopathy genes also show predominantly de novo inheritance.

- **Typified by incomplete penetrance –** *Penetrance - A situation in which mutation carriers do not show clinically evident phenotypic abnormalities.* This modifier should be used for conditions typified by incomplete penetrance that may not manifest even during a full lifespan (80 yrs of age taken as standard). Patients manifesting disease in these conditions also tend to have a higher than expected rate of additional mutations in the same pathway ie second hits which increase penetrance.

Example: *DSC2* and arrhythmogenic cardiomyopathy. Many of the genes associated with inherited cardiac conditions exhibit reduced penetrance.

- **Typified by age-related onset –** conditions which have age related onset
- **Requires heterozygosity -***Disease is only observed (or observed with greater severity) in heterozygous female patients compared with hemizygous male individuals*.

Example: Craniofrontonasal dysplasia due to *EFNB1* - which requires heterozygosity and would not manifest (fully) if hemizygous.  Importantly the mutant allele can be inherited from a normal or very mildly affected father.

**Disease-associated variant classes:**

**List variant classes (using sequence ontology (SO) terms) in this gene proven to cause this disease:**

Consider whether the disease is associated with:

- missense & in-frame variants

- Protein truncating codons (PTCs) (aka premature truncating variants or loss of function (LoF) or radical)

for PTCs need to consider whether nonsense mediated decay (NMD) competent or not

In practice it is useful to know whether a gene-disease pair is associated with missense only, LoF truncating only, or both

See matrix below for variant class SO terms

List SO terms:

**List additional variant classes predicted to lead to the same functional consequence:**

Other variant classes that could be predicted to lead to the same functional consequence based on inferred mechanism (score 4 or 5, see matrix below) and therefore might cause the same phenotype.

The semi-quantitative scale is characterised from first principles by expert evaluation.

| almost never | 1 |
| --- | --- |
| unlikely | 2 |
| possible | 3 |
| probable | 4 |
| almost always | 5 |

|  | Predicted functional consequence | | | | |
| --- | --- | --- | --- | --- | --- |
|  | **Altered gene product level** | | | **Altered gene product**  **Sequence** | No effect  **(Functionally normal)** |
|  | **Increased gene product level** | level reduction | |  |  |
| **SO term** |  | **Decreased gene product level** | **Absent gene product** |  |  |
| splice_region_variant | 3 | 3 | 2 | 2 | 3 |
| splice_acceptor_variant | 1 | 4 | 3 | 3 | 2 |
| splice_acceptor_variant_NMD_triggering | 1 | 5 | 5 | 2 | 1 |
| splice_acceptor_variant_NMD_escaping | 1 | 2 | 1 | 4 | 1 |
| splice_donor_variant | 1 | 4 | 4 | 4 | 2 |
| splice_donor_variant_NMD_triggering | 1 | 5 | 5 | 2 | 1 |
| splice_donor_variant_NMD_escaping | 1 | 2 | 1 | 4 | 1 |
| start_lost | 1 | 5 | 5 | 2 | 1 |
| frameshift_variant | 1 | 5 | 5 | 2 | 1 |
| frameshift_variant_NMD_triggering | 1 | 5 | 5 | 2 | 1 |
| frameshift_variant_NMD_escaping | 1 | 2 | 1 | 4 | 1 |
| stop_gained | 1 | 5 | 5 | 2 | 1 |
| stop_gained_NMD_triggering | 1 | 5 | 5 | 2 | 1 |
| stop_gained_NMD_escaping | 1 | 2 | 1 | 4 | 1 |
| stop_lost | 1 | 1 | 1 | 4 | 1 |
| missense_variant | 2 | 2 | 2 | 5 | 1 |
| inframe_insertion | 2 | 2 | 1 | 5 | 1 |
| inframe_deletion | 2 | 2 | 1 | 5 | 1 |
| 5_prime_UTR_variant | 2 | 2 | 1 | 1 | 5 |
| 3_prime_UTR_variant | 2 | 2 | 1 | 1 | 5 |
| synonymous_variant | 2 | 2 | 2 | 2 | 5 |
| intron_variant | 2 | 2 | 2 | 2 | 5 |
| regulatory_region_variant | 2 | 2 | 1 | 1 | 5 |
| intergenic_variant | 1 | 1 | 1 | 1 | 5 |

**Disease-associated variant consequences:**

Once the variant classes associated with the disease are known, map these to the high level terms using the matrix above.

High level terms to describe variant consequences:

- Altered gene product level

- Unspecified change in gene product level

- Decreased gene product level

- Absent gene product

- Increased gene product level

- Altered gene product sequence

- Functionally normal

Notes:

- **Decreased/absent gene product level** – for example PTCs (protein truncating), gene-disrupting SVs, and gene-deletions (assuming NMD-competent PTC, and with caveats about splicing)
- **Increased gene product level** – for example non-disruptive gene duplications, some promoter or enhancer variants
- **Altered gene product sequence** – for example NMD-incompetent PTCs, other length-changing variants (in frame indels, stop loss), and missense.  Downstream mechanisms can be diverse: functionally null - misfolded, mislocalised, inactive, hypomorphic; disruptive presence of abnormal protein (gain of function, dominant negative etc)

**Narrative summary of molecular mechanisms:**

Summary of mechanism for gene-disease pair. For example, ‘Mechanism is likely loss of function of *NF1* due to reduction/absence of gene product or altered gene product sequence.’

Mention of specific mechanisms such as ‘dominant negative’ can be recorded here as well as any other useful information captured in the literature review section. Please record information on structural variants if they are relevant for this gene-disease pair.

**Please record information about other ACMG evidence types if it is available (this is not mandatory and we appreciate this won’t be available for many gene-disease pairs).**

Examples and notes:

**For PM1:**

- Is there a mutational hot spot for this gene-disease pair? If so, is there a guideline and genomic coordinates?
- Is there sufficient data to calculate etiological fraction by domain? If so please record it here

Notes:

Etiological fraction applied per domain/region can provide following levels of evidence for PM1: Strong >0.95, moderate >0.90, supporting >0.80.

For more information on Etiological Fraction see:

https://genomemedicine.biomedcentral.com/articles/10.1186/s13073-019-0616-z (PMID: 30696458)

See Walsh et al 2020 PMID: 32893267 for specific examples of this for LQTS and Brugada genes, in particular table 2: https://www.nature.com/articles/s41436-020-00946-5/tables/2

**For PVS1:**

- Is there any important transcript information, i.e. are there certain transcripts that are or are not relevant to this disease.

Notes:

For example in the *TTN* gene, truncating variants are only known to cause disease if impacting exons that are constitutively expressed in the heart (i.e. PVS1 doesn’t always apply)

See Whiffin et al 2018 PMID: 29369293 for examples of tailored application of PSV1

**For BS1/PM2**:

- What is the maximum credible population allele frequency for this disease?

Notes:

maximum credible population AF = prevalence x maximum allele contribution x 1/penetrance

See https://www.nature.com/articles/gim201726 (PMID: 28518168) for more information about maximum credible population AF and examples

See Walsh et al 2020 PMID: 32893267 for an example of filtering allele freq thresholds for LQTS and Brugada genes.

**For PP2**:

- If there are no mutational hotspots, what is the etiological fraction for the whole gene (this may not be available for very rare conditions where there may be insufficient case numbers to do the analysis).

Notes:

Etiological fraction applied per gene can provide following levels of evidence for PP2:

Strong >0.95, moderate >0.90, supporting >0.80.

For more information https://www.ncbi.nlm.nih.gov/pmc/articles/PMC5116235/ (PMID: 27532257)

**General notes:**

-If monoallelic and biallelic inheritance can cause the same disease, they should be recorded as separate entities if:

the variants causing the recessive version are truly recessive (i.e. would not be pathogenic in isolation)

biallelic variants lead to a different phenotype (not just a change in severity)

-If a dominant variant can also be seen on both alleles but the outcome is essentially the same disease, then this should be categorised as one entity using dominant and monoallelic.

For example:

AD and AR *DSC2* causing isolated ARVC are one disease gene pair
AR *DSC2* causing ARVC with cutaneous manifestations is a separate disease gene pair.
