## Additional File 3 for "Beyond gene-disease validity: capturing structured data on inheritance, allelic-requirement, disease-relevant variant classes, and disease mechanism for inherited cardiac conditions"

### INHERITED ARRHYTHMIA SYNDROMES

##### BRUGADA SYNDROME

| Gene | *SCN5A* |
| --- | --- |
| OMIM gene number | 600163 |
| Referral indication | Brugada syndrome |
| Disease grouping | Brugada syndrome |
| **Disease name** | ***SCN5A-*related Brugada syndrome** |
| MONDO ID | 0011001 |
| Gene disease validity (ClinGen) | DEFINITIVE |
| Inheritance | Autosomal dominant |
| Allelic requirement | Monoallelic autosomal |
| Inheritance modifiers | Typified by incomplete penetrance |
| Disease-associated variant consequence | Decreased gene product level; altered gene product sequence |
| Variant classes reported with evidence of pathogenicity | Splice region; splice acceptor; splice donor; frameshift variant NMD triggering; stop gained variant NMD triggering; missense; inframe insertion; inframe deletion |
| Restricted repertoire of pathogenic variants | NA |
| PMIDs | 9521325; 11748104; NBK1517; 25905440; 32850980; 25829473; 17075016; 17442746; 20564468; 20031634; 33164571; 29798782; 32533946; 20129283; 33131149; 30203441; 32893267; 29959160 |

*SCN5A-*related Brugada syndrome is caused by **decreased gene product level or altered gene product sequence** due to a variety of mechanisms (e.g. decreased expression of cardiac sodium channel, Nav1.5 in the sarcolemma, expression of non-functional channels, or altered gating properties leading to a decreased I_Na_ sodium current (e.g., delayed activation or earlier or faster inactivation)) (PMID: 29798782). The disease mechanism is loss of function.

To date, *SCN5A* is the only gene classified as having definitive evidence as a cause of monogenic Brugada syndrome (BrS) by ClinGen (PMID: 29959160). *SCN5A* pathogenic variants are identified in approx. 20- 30% of cases of European ancestry (PMID: 30139433; PMID 33164571). This contribution may differ in other populations.

*SCN5A-*related Brugada syndrome is characterized by autosomal dominant inheritance with incomplete penetrance (PMID 9521325; 11748104; 25905440; 20031634; 33164571; NBK1517).

Hundreds of variants, both truncating and non-truncating, have been described in association with Brugada syndrome. Kapplinger et al 2010 (PMID: 20129283) identified 293 distinct variants in *SCN5A*: 193 missense, 32 nonsense, 38 frameshift, 21 splice-site, and 9 in-frame deletions/insertions. More recently Walsh et al 2020 (PMID: 32893267) found that non truncating variants were highly enriched in European cases in the *SCN5A* transmembrane regions.

Gain of function variants are associated with Long QT syndrome. Although loss of function appears to be the accepted mechanism in Brugada syndrome the *SCN5A* genotype/phenotype association is still not completely understood. Uncommonly, a single variant can cause both loss of peak current and gain of late current and can lead to a mixed phenotype of LQTS and BrS (PMID: 29806494)*.*

##### CATECHOLAMINERGIC POLYMORPHIC VENTRICULAR TACHYCARDIA (CPVT)

| Gene | *RYR2* |
| --- | --- |
| OMIM gene number | 180902 |
| Referral indication | Catecholaminergic polymorphic ventricular tachycardia (CPVT) |
| Disease grouping | Classic CPVT phenotype |
| **Disease name** | ***RYR2*-related catecholaminergic polymorphic ventricular tachycardia (CPVT)** |
| MONDO ID | 0017990 |
| Gene disease validity (ClinGen) | DEFINITIVE |
| Inheritance | Autosomal dominant |
| Allelic requirement | Monoallelic autosomal |
| Inheritance modifiers | Typified by incomplete penetrance |
| Disease-associated variant consequence | Altered gene product sequence |
| Variant classes reported with evidence of pathogenicity | Missense; exon deletion |
| Restricted repertoire of pathogenic variants | Exon 3 deletion |
| PMIDs | 29453246; 11208676; 12093772; 17875969; 19216760; 23479668; 24394973; 26018045; 17081562; 30696458; 31112425 |

*RYR2* is associated with CPVT with autosomal dominant inheritance. The allelic requirement for pathogenicity is monoallelic autosomal, and the disease is typified by incomplete penetrance. Disease is due to an **altered gene product sequence**.

The first reports of *RYR2* variants in CPVT included numerous examples of *de novo* inheritance (Priori et al, 2001, PMID:11208676; Priori et al, 2002, PMID:12093772), although *RYR2* missense variants often occur through familial autosomal dominant inheritance.

The predicted functional consequence of *RYR2* pathogenic variants is Altered gene product sequence. The majority of causative *RYR2* variants in CPVT patients are heterozygous missense variants which are detected in up to 60% of cases with a definitive CPVT diagnosis (Kapplinger et al, 2018, PMID:29453246) and act through a gain-of-function mechanism. Pathogenic variants in CPVT cases tend to be clustered in a number of regions in the RYR2 gene/protein:

- Original exon hotspots: 3–15, 44–50, 83–90 and 93–105 (George et al, 2007, PMID:17081562).

- Updated exon hotspots: 3, 8, 14, 43, 47–49, 81, 83, 88–90, 93, 95, 97–101, 103, 105 (Kapplinger et al, 2018, PMID:29453246).

- Hotspots based on unsupervised clustering algorithm (not restricted to exon boundaries): amino acid residues 2138–2538, 3935–4196 and 4721–4959 (Walsh et al, 2019, PMID:30696458).

Rare cases of whole exon deletions of *RYR2* exon 3 have been described that can lead to a CPVT and/or left ventricular non-compaction phenotype (Bhuiyan et al, 2007, PMID:17875969; Marjamaa et al, 2009, PMID:19216760; Szentpali et al, 2013, PMID:23479668; Onho et al, 2014, PMID:24394973; Campbell et al, 2015, PMID:26018045; Mazzarotto et al, 2021, PMID:33500567).

| Gene | *CASQ2* |
| --- | --- |
| OMIM gene number | 114251 |
| Referral indication | Catecholaminergic polymorphic ventricular tachycardia (CPVT) |
| Disease grouping | Classic CPVT phenotype |
| **Disease name** | ***CASQ2*-related catecholaminergic polymorphic ventricular tachycardia (CPVT)** |
| MONDO ID | 0017990 |
| Gene disease validity (ClinGen) | DEFINITIVE |
| Inheritance | Autosomal recessive |
| Allelic requirement | Biallelic autosomal |
| Inheritance modifiers | NA |
| Disease-associated variant consequence | Absent gene product level; altered gene product sequence |
| Variant classes reported with evidence of pathogenicity | Splice region; splice donor variant NMD triggering; splice acceptor variant NMD triggering; frameshift variant NMD triggering; stop gained variant NMD triggering; missense |
| Restricted repertoire of pathogenic variants | NA |
| PMIDs | 12386154; 16908766; 21618644; 15176429; 32693635; 27157848 |

*CASQ2* is an established gene for autosomal recessive CPVT (the second most common genetic cause of the disease, responsible for up to 5% of cases); disease is due to **absent gene product level or altered gene product sequence**. For the ClinGen curation, the maximum points were achieved with only a small subset of initial genetic and experimental reports leading to a definitive classification.

Biallelic loss-of-function variants in *CASQ2* (both homozygous and compound heterozygous) have been reported in numerous CPVT probands, including frameshift, nonsense and splice donor/acceptor variants, as well as other splice region variants with verified effects on splicing, and missense variants with verified loss-of-function effects.

| Gene | *CASQ2* |
| --- | --- |
| OMIM gene number | 114251 |
| Referral indication | Catecholaminergic polymorphic ventricular tachycardia (CPVT) |
| Disease grouping | Classic CPVT phenotype |
| **Disease name** | ***CASQ2*-related catecholaminergic polymorphic ventricular tachycardia (CPVT)** |
| MONDO ID | 0017990 |
| Gene disease validity (ClinGen) | MODERATE |
| Inheritance | Autosomal dominant |
| Allelic requirement | Monoallelic autosomal |
| Inheritance modifiers | Typified by incomplete penetrance |
| Disease-associated variant consequence | Decreased gene product level; Altered gene product sequence |
| Variant classes reported with evidence of pathogenicity | Splice region; splice donor variant NMD triggering; splice acceptor variant NMD triggering; frameshift variant NMD triggering; stop gained variant NMD triggering; missense |
| Restricted repertoire of pathogenic variants | NA |
| PMIDs | 12386154; 16908766; 21618644; 15176429; 32693635; 27157848 |

*CASQ2* is an established gene for autosomal recessive CPVT (the second most common genetic cause of the disease, responsible for up to 5% of cases).

Recent reports have also associated monoallelic or heterozygous *CASQ2* variants with CPVT. Although the evidence for this association is not definitive, available data suggests that disease is likely due to **decreased gene product level or altered gene product sequence**. The main evidence for autosomal dominant *CASQ2* association comes from an international multi-centre study describing CPVT patients with *CASQ2* variants (Ng et al, 2020, PMID:32693635). This study included 12 probands with heterozygous variants in *CASQ2*, as well as an assessment of heterozygous relatives of probands with homozygous/compound heterozygous *CASQ2* variants (8/37 of these heterozygous relatives had a positive CPVT phenotype). While this study provides a substantive body of evidence to support autosomal dominant *CASQ2* association with CPVT, the data should be cautiously interpreted. The multi-centre nature of the study precluded standardised phenotyping of the probands and relatives and therefore not every phenotype-positive individual may have a definitive diagnosis of CPVT. Additionally, several of the variants described have a gnomAD population minor allele frequency that is incompatible with being a penetrant autosomal dominant variant for a disease with the prevalence of CPVT (>1x10-5). The variants also included presumed nonsense mediated decay-escaping C-terminal truncating variants and a splice region variant without a proven effect on splicing. In contrast to 97% penetrance for homozygous/compound heterozygous individuals, penetrance for heterozygotes was only 33% although it is expected that the population level penetrance of *CASQ2* heterozygous variants will be lower still. Further support for the pathogenicity of heterozygous *CASQ2* variants comes from a study that described the heterozygous p.Lys180Arg variant segregating with disease in a family (the published LOD score was 3.0 although there were only five meioses between genotype and phenotype positive individuals) (Gray et al, 2016, PMID:27157848).

| Gene | *CALM1* |
| --- | --- |
| OMIM gene number | 114180 |
| Referral indication | Catecholaminergic polymorphic ventricular tachycardia (CPVT) |
| Disease grouping | Atypical CPVT phenotype |
| **Disease name** | ***CALM1*-related catecholaminergic polymorphic ventricular tachycardia (CPVT)** |
| MONDO ID | 0017990 |
| Gene disease validity (ClinGen) | MODERATE |
| Inheritance | Autosomal dominant |
| Allelic requirement | Monoallelic autosomal |
| Inheritance modifiers | Typically *de novo* |
| Disease-associated variant consequence | Altered gene product sequence |
| Variant classes reported with evidence of pathogenicity | Missense |
| Restricted repertoire of pathogenic variants | NA |
| PMIDs | 31983240; 23040497; 25557436; 32929985; 24563457; 26309258; 31170290 |

The disease mechanism in *CALM1*-related CPVT is likely a dominant negative effect caused by **altered gene product sequence** which reduces the calcium dependent inactivation of the calcium channel CaV1.2, resulting in increased inward calcium channel current (ICaL) and repolarization delay.

The three *CALM* genes encode an identical 149aa protein, calmodulin. The *CALM1* gene is located on chromosome 14, *CALM2* on chromosome 2 and *CALM3* on chromosome 19. Calmodulin protein is involved in many calcium-dependent intracellular processes. All three *CALM* genes have been classified as Definitive for LQTS and Moderate for CPVT, certain variants are only associated with one or other phenotype, whereas others are associated with a mixed or variable phenotypes.

All variants with evidence of pathogenicity identified in the *CALM* genes are missense: at least 35 distinct missense variants have been identified and reported in the International Calmodulinopathy Registry including 11 (31%) in CALM1, 16 (46%) in CALM2, and 8 (23%) in CALM3. The majority of variants across the *CALM* genes affect amino acid residues in the EF-hand Ca2+ binding loop III and IV. Some variants have additional evidence in that the same substitution has been seen in paralogous *CALM* genes, or that the same substitution has been seen in a related phenotype (LQT). Most variants are unique, but 9 were present in more than one index case and among these, three (p.Asn98Ser, p.Asp130Gly, and p.Phe142Leu, identified in 10, 5, and 4 families, respectively.) appear to be recurrent. While p.Asp130Gly and p.Phe142Leu have always been reported as associated with the LQTS phenotype, the p.Asn98Ser has phenotypic variability, including LQTS, CPVT, idiopathic VF and sudden death. The majority of *CALM* variants are *de novo*.

Pathogenic variants in the *CALM* genes have been associated with: presentation in infancy or early childhood (up to 5 years); marked sinus bradycardia or atrioventricular block and QT prolongation; and, predominantly with *CALM1* variants, a mild-to-severe neurological impairment, including seizures, development delay, motor and/or cognitive disability.

The phenotype most frequently shown by patients with *CALM* variants is LQTS, but some patients display other phenotypes including CPVT, idiopathic VF and sudden unexplained death. Some variants have been associated with both LQTS and CPVT, however Crotti et al (PMID: 31170290) report that "despite the relatively small numbers of cases, a significant association (P = 0.001) was observed between location of mutation and phenotype (Supplementary material online, Figure S1). Indeed, a pathogenic variant in EF-hand IV Ca2+ binding loop was found in the majority (17/32, 53%) of CALM-LQTS index cases but in only one of the nine CALM-CPVTs (11%). Conversely, variants identified in CALM-CPVT index cases were mostly located either in EF-hand III (n = 5, 56%), or in the inter-EF hand I-II linker (n = 3, 33%)."

For the ClinGen CPVT curation, *CALM1* scored in the moderate range based on two reported variants in patients with CPVT or mixed CPVT/LQT phenotypes (p.Asn54Ile and p.Asn98Ser) and associated functional data. However, due to the unique characteristics of the calmodulin genes and their gene-disease associations, it was agreed by the expert panel to include them in this curation. The reasons were: 1) all three genes have already been established as disease-causing for an inherited arrhythmia syndrome (LQTS), 2) there are multiple examples of patients presenting with a phenotype indistinguishable from CPVT for each, either *de novo* or supported by functional evidence and 3) the genes encode for identical proteins, are all expressed in the heart and equivalent *de novo* variants in the three genes have been shown to lead to similar phenotypes, highlighting the functional equivalence of these genes.

| Gene | *CALM2* |
| --- | --- |
| OMIM gene number | 114182 |
| Referral indication | Catecholaminergic polymorphic ventricular tachycardia (CPVT) |
| Disease grouping | Atypical CPVT phenotype |
| **Disease name** | ***CALM2*-related catecholaminergic polymorphic ventricular tachycardia (CPVT)** |
| MONDO ID | 0017990 |
| Gene disease validity (ClinGen) | MODERATE |
| Inheritance | Autosomal dominant |
| Allelic requirement | Monoallelic autosomal |
| Inheritance modifiers | Typically *de novo* |
| Disease-associated variant consequence | Altered gene product sequence |
| Variant classes reported with evidence of pathogenicity | Missense |
| Restricted repertoire of pathogenic variants | NA |
| PMIDs | 31983240; 27100291; 24917665; 25557436; 26309258; 31170290 |

The disease mechanism in *CALM2*-related CPVT is likely a dominant negative effect caused by **altered gene product sequence** which reduces the calcium dependent inactivation of the calcium channel CaV1.2, resulting in increased inward calcium channel current (ICaL) and repolarization delay.

The three CALM genes encode an identical 149aa protein, calmodulin. The *CALM1* gene is located on chromosome 14, *CALM2* on chromosome 2 and *CALM3* on chromosome 19. Calmodulin protein is involved in many calcium-dependent intracellular processes. All three *CALM* genes have been classified as Definitive for LQTS and Moderate for CPVT, certain variants are only associated with one or other phenotype, whereas others are associated with a mixed or variable phenotypes.

All variants with evidence of pathogenicity identified in the CALM genes are missense: at least 35 distinct missense variants have been identified and reported in the International Calmodulinopathy Registry including 11 (31%) in CALM1, 16 (46%) in CALM2, and 8 (23%) in CALM3. The majority of variants across the CALM genes affect amino acid residues in the EF-hand Ca2+ binding loop III and IV. Some variants have additional evidence in that the same substitution has been seen in paralogous CALM genes, or that the same substitution has been seen in a related phenotype (LQTS). Most variants are unique, but 9 were present in more than one index case and among these, three (p.Asn98Ser, p.Asp130Gly, and p.Phe142Leu, identified in 10, 5, and 4 families, respectively.) appear to be recurrent. While variants p.Asp130Gly and p.Phe142Leu have always been reported as associated with the LQTS phenotype, the p.Asn98Ser has phenotypic variability, including LQTS, CPVT, idiopathic VF and sudden death. The majority of *CALM* variants are *de novo*.

Pathogenic variants in the *CALM* genes have been associated with: presentation in infancy or early childhood (up to 5 years); marked sinus bradycardia or atrioventricular block and QT prolongation; and, predominantly with *CALM1* variants, a mild-to-severe neurological impairment, including seizures, development delay, motor and/or cognitive disability.

The phenotype most frequently shown by patients with *CALM* variants is LQTS, but some patients display other phenotypes including CPVT, idiopathic VF and sudden unexplained death. Some variants have been associated with both LQTS and CPVT, however Crotti et al (PMID: 31170290) report that "despite the relatively small numbers of cases, a significant association (P = 0.001) was observed between location of mutation and phenotype (Supplementary material online, Figure S1). Indeed, a pathogenic variant in EF-hand IV Ca2+ binding loop was found in the majority (17/32, 53%) of CALM-LQTS index cases but in only one of the nine CALM-CPVTs (11%). Conversely, variants identified in CALM-CPVT index cases were mostly located either in EF-hand III (n = 5, 56%), or in the inter-EF hand I-II linker (n = 3, 33%)."

For the ClinGen CPVT curation, *CALM2* scored in the moderate range based on four reported variants in patients with CPVT or mixed CPVT/LQT phenotypes (p.Glu46Lys in two *de novo* cases, p.Asn98Ser and p.Asp132Glu) and associated functional data.

However, due to the unique characteristics of the calmodulin genes and their gene-disease associations, it was agreed by the expert panel to include them in this curation. The reasons were: 1) all three genes have already been established as disease-causing for an inherited arrhythmia syndrome (LQTS), 2) there are multiple examples of patients presenting with a phenotype indistinguishable from CPVT for each, either *de novo* or supported by functional evidence and 3) the genes encode for identical proteins, are all expressed in the heart and equivalent *de novo* variants in the three genes have been shown to lead to similar phenotypes, highlighting the functional equivalence of these genes.

| Gene | *CALM3* |
| --- | --- |
| OMIM gene number | 114183 |
| Referral indication | Catecholaminergic polymorphic ventricular tachycardia (CPVT) |
| Disease grouping | Atypical CPVT phenotype |
| **Disease name** | ***CALM3*-related catecholaminergic polymorphic ventricular tachycardia (CPVT)** |
| MONDO ID | 0017990 |
| Gene disease validity (ClinGen) | MODERATE |
| Inheritance | Autosomal dominant |
| Allelic requirement | Monoallelic autosomal |
| Inheritance modifiers | Typically *de novo* |
| Disease-associated variant consequence | Altered gene product sequence |
| Variant classes reported with evidence of pathogenicity | Missense |
| Restricted repertoire of pathogenic variants | NA |
| PMIDs | 31983240; 27516456; 31170290 |

The disease mechanism in *CALM1*-related CPVT is likely a dominant negative effect caused by **altered gene product sequence** which reduces the calcium dependent inactivation of the calcium channel *Ca*_v_*1*.2, resulting in increased inward calcium channel current (ICaL) and repolarization delay.

The three *CALM* genes encode an identical 149aa protein, calmodulin. The *CALM1* gene is located on chromosome 14, *CALM2* on chromosome 2 and *CALM3* on chromosome 19. Calmodulin protein is involved in many calcium-dependent intracellular processes. All three *CALM* genes have been classified as Definitive for LQTS and Moderate for CPVT, certain variants are only associated with one or other phenotype, whereas others are associated with a mixed or variable phenotypes.

All variants with evidence of pathogenicity identified in the *CALM* genes are missense: at least 35 distinct missense variants have been identified and reported in the International Calmodulinopathy Registry including 11 (31%) in CALM1, 16 (46%) in CALM2, and 8 (23%) in CALM3. The majority of variants across the CALM genes affect amino acid residues in the EF-hand Ca2+ binding loop III and IV. Some variants have additional evidence in that the same substitution has been seen in paralogous *CALM* genes, or that the same substitution has been seen in a related phenotype (LQTS). Most variants are unique, but 9 were present in more than one index case and among these, three (p.Asn98Ser, p.Asp130Gly, and p.Phe142Leu, identified in 10, 5, and 4 families, respectively.) appear to be recurrent. While variants p.Asp130Gly and p.Phe142Leu have always been reported as associated with the LQTS phenotype, the variant p.Asn98Ser has phenotypic variability, including LQTS, CPVT, idiopathic VF and sudden death. The majority of *CALM* variants are *de novo*.

Pathogenic variants in the *CALM* genes have been associated with: presentation in infancy or early childhood (up to 5 years); marked sinus bradycardia or atrioventricular block and QT prolongation; and, predominantly with *CALM1* variants, a mild-to-severe neurological impairment, including seizures, development delay, motor and/or cognitive disability.

The phenotype most frequently shown by patients with *CALM* variants is LQTS, but some patients display other phenotypes including CPVT, idiopathic VF and sudden unexplained death. Some variants have been associated with both LQTS and CPVT, however Crotti et al (PMID: 31170290) report that "despite the relatively small numbers of cases, a significant association (P = 0.001) was observed between location of mutation and phenotype (Supplementary material online, Figure S1). Indeed, a pathogenic variant in EF-hand IV Ca2+ binding loop was found in the majority (17/32, 53%) of CALM-LQTS index cases but in only one of the nine CALM-CPVTs (11%). Conversely, variants identified in CALM-CPVT index cases were mostly located either in EF-hand III (n = 5, 56%), or in the inter-EF hand I-II linker (n = 3, 33%)."

For the ClinGen CPVT curation, *CALM3* scored in the limited range based on two reported variants in patients with CPVT or mixed CPVT/LQT phenotypes (p.Asp132Glu and p.Ala103Val) and associated functional data.

However, due to the unique characteristics of the calmodulin genes and their gene-disease associations, it was agreed by the expert panel to include them in this curation. The reasons were: 1) all three genes have already been established as disease-causing for an inherited arrhythmia syndrome (LQTS), 2) there are multiple examples of patients presenting with a phenotype indistinguishable from CPVT for each, either *de novo* or supported by functional evidence and 3) the genes encode for identical proteins, are all expressed in the heart and equivalent *de novo* variants in the three genes have been shown to lead to similar phenotypes, highlighting the functional equivalence of these genes.

| Gene | *TECRL* |
| --- | --- |
| OMIM gene number | 614021 |
| Referral indication | Catecholaminergic polymorphic ventricular tachycardia (CPVT) |
| Disease grouping | Atypical CPVT phenotype |
| **Disease name** | ***TECRL*-related catecholaminergic polymorphic ventricular tachycardia (CPVT)** |
| MONDO ID | 0017990 |
| Gene disease validity (ClinGen) | DEFINITIVE |
| Inheritance | Autosomal recessive |
| Allelic requirement | Biallelic autosomal |
| Inheritance modifiers | NA |
| Disease-associated variant consequence | Absent gene product level; altered gene product sequence |
| Variant classes reported with evidence of pathogenicity | Splice donor variant NMD triggering; stop gained variant NMD triggering; missense; exon deletion |
| Restricted repertoire of pathogenic variants | Exon 2 deletion |
| PMIDs | 27861123; 30790670; 33367594; 32173957 |

**Absent gene product level and altered gene product sequence** of *TECRL* is associated with CPVT. The disease mechanism appears to be loss of function.

*TECRL* is rarely associated with CPVT, but several reports have described biallelic loss of function variants in cases. These include both homozygous and compound heterozygous inheritance, with a variety of variant types described – splice donor variant, stop gained, exon deletion (exon 2), missense variants (including homozygous p.Arg196Gln detected in 2 patients with exome sequencing), and a large duplication encompassing all of the *TECRL* gene with an uncertain consequence.

These cases presented with phenotypic features typical of CPVT, including exercise and emotion induced syncope and cardiac arrest and ventricular arrhythmias during exercise testing. A mild prolonged QT interval was observed in several cases, especially after stimulation by epinephrine or exercise, although overall the phenotypes are much more typical of CPVT than LQTS.

| Gene | *TRDN* |
| --- | --- |
| OMIM gene number | 603283 |
| Referral indication | Catecholaminergic polymorphic ventricular tachycardia (CPVT) |
| Disease grouping | Atypical CPVT phenotype |
| **Disease name** | ***TRDN*-related catecholaminergic polymorphic ventricular tachycardia (CPVT)** |
| MONDO ID | 0017990 |
| Gene disease validity (ClinGen) | DEFINITIVE |
| Inheritance | Autosomal recessive |
| Allelic requirement | Biallelic autosomal |
| Inheritance modifiers | NA |
| Disease-associated variant consequence | Absent gene product level; altered gene product sequence |
| Variant classes reported with evidence of pathogenicity | Frameshift variant NMD triggering; stop gained variant NMD triggering; missense; exon deletion |
| Restricted repertoire of pathogenic variants | Exon 2 deletion |
| PMIDs | 22422768; 26200674; 26768964; 30479949; 25922419 |

**Absent gene product level and altered gene product sequence** of *TRDN* is associated with CPVT. The disease mechanism appears to be loss of function.

*TRDN* is rarely associated with CPVT, but several reports have described biallelic loss of function variants in cases. These include both homozygous and compound heterozygous variants, with a variety of variant types described - stop gained, frameshift, exon deletion (exon 2), intronic variants (with proven effect on splicing) and missense variants (proven to lead to degraded protein).

Biallelic loss of function variants in *TRDN* have also been associated with LQTS with an atypical presentation (ClinGen: strong evidence). This, "triadin knockout syndrome" can lead to variable phenotypes.

##### LONG QT SYNDROME (LQTS)

| Gene | *KCNQ1* |
| --- | --- |
| OMIM gene number | 607542 |
| Referral indication | Long QT Syndrome (LQTS) |
| Disease grouping | Familial Long QT Syndrome |
| **Disease name** | ***KCNQ1*-related long QT Syndrome** |
| MONDO ID | 0008646 |
| Gene disease validity (ClinGen) | DEFINITIVE |
| Inheritance | Autosomal dominant; autosomal recessive |
| Allelic requirement | Monoallelic autosomal; biallelic autosomal |
| Inheritance modifiers | Typified by incomplete penetrance |
| Disease-associated variant consequence | Decreased gene product level; altered gene product sequence |
| Variant classes reported with evidence of pathogenicity | Splice acceptor; splice donor; frameshift; frameshift variant NMD triggering; stop gained; stop gained variant NMD triggering; missense; inframe insertion; inframe deletion; structural variants (whole exon deletions); structural variants (intragenic multi exon tandem duplication) |
| Restricted repertoire of pathogenic variants | NA |
| PMIDs | 31983240; 23900354; 23392653; 23591039; 27041150; 28944242; NBK1129; 15840476; 19716085; 9927399; 22995932; 29650123; 21185501; 27807201; 17470695; 25854863; 29532034; 25174857; 32893267; 22456477; 21787999; 27262388; 19841300; 29504689 |

Pathogenic variants in *KCNQ1* cause LQTS due to **decreased gene product level or altered gene product sequence.** This leads to loss of function due to a variety of mechanisms including defects in ion permeation, channel gating and trafficking.

*KCNQ1* variants account for approx. 30%-45% of LQTS cases (PMID: 22456477; PMID: 21787999; PMID: 27262388; PMID: 29504689).

*KCNQ1*-related LQTS is typically inherited as an autosomal dominant trait characterised by incomplete penetrance. Rarely biallelic *KCNQ1* variants can cause autosomal recessive LQTS or Jervell and Lange-Nielsen syndrome (JLNS) (severe cardiac phenotype and sensorineural hearing loss). JLNS results when there is complete absence of IK_s_ (slowly activating delayed rectifier potassium channel/current) (PMID: 23591039).

It has been noted that approximately 10% of genotype positive LQT patients have more than 1 pathogenic variant in >=1 LQTS-related gene (PMID: 15840476; PMID: 19716085). Biallelic disease appears to be more penetrant with a more severe phenotype.

Missense variants are responsible for the majority of KCNQ1-related LQTS cases although truncating variants including nonsense, frameshift, splice site and structural variants (multi exon deletions and a multi exon duplication (in-frame tandem repeat of exons 3-6, leading to duplication of the second to fifth transmembrane domains of the channel)) are also reported (PMID: 19716085; PMID: 18774102; PMID: 25174857).

ClinGen have concluded there is sufficient evidence for haploinsufficiency. Variants conferring a dominant negative effect have also been described. Moss et al (PMID: 17470695) reported that dominant-negative variants are associated with a more severe phenotype than variants conferring haploinsufficiency.

Phenotypes can differ according to regionality with some regions associated with more malignant phenotypes e.g. C-loop variants are associated with higher risk of sudden cardiac death (PMID: 22456477*).* In addition, there are gene regions where there is a high confidence for pathogenicity e.g. transmembrane regions and C-terminus domains (PMID: 32893267; PMID: 19841300).

| Gene | *KCNH2* |
| --- | --- |
| OMIM gene number | 152427 |
| Referral indication | Long QT Syndrome (LQTS) |
| Disease grouping | Familial Long QT Syndrome |
| **Disease name** | ***KCNH2*-related long QT Syndrome** |
| MONDO ID | 0013367 |
| Gene disease validity (ClinGen) | DEFINITIVE |
| Inheritance | Autosomal dominant |
| Allelic requirement | Monoallelic autosomal |
| Inheritance modifiers | Typified by incomplete penetrance |
| Disease-associated variant consequence | Decreased gene product level; altered gene product sequence |
| Variant classes reported with evidence of pathogenicity | Splice acceptor; splice donor; frameshift variant NMD triggering; stop gained variant NMD triggering; missense; inframe insertion; inframe deletion; structural variants (whole exon deletions & duplications) |
| Restricted repertoire of pathogenic variants | NA |
| PMIDs | 31983240; 26320108; 27761161; NBK1129; 15840476; 18774102; 19926013; 32893267; 21185499; 10491368; 10841244; 12621127; 19716085; 21185501; 9927399; 22995932; 29650123; 25417810; 24932360 |

*KCNH2-*related LQTS is due to **decreased gene product level or altered gene product sequence**. The disease mechanism is **loss of function** due to a variety of mechanisms including disruption of synthesis of channel subunits, reduction in intracellular transport or trafficking, defects in ion permeation or channel gating.

Both haploinsufficiency and a dominant negative effect are proposed mechanisms causing loss of function of *KCNH2*. About 60% of LQT2 mutations are missense variants, the remaining 40% are nonsense, frameshift, insertions, deletions, duplications (intragenic tandem duplication), or involve a splice site.

*KCNH2* variants account for approximately 30% of LQTS cases (PMID: 24932360).

Walsh et al showed that non truncating variants in *KCNH2* are enriched in ion channel transmembrane regions and specific N-terminus and C-terminus domains and have >95% probability of pathogenicity (PMID: 32893267).

Shimizu et al report correlations between specific variant types and location and clinical phenotype (PMID: 19926013). Patients with missense variants in the transmembrane pore region have significantly higher cardiac event rates than those with missense variants in either N-terminus, transmembrane non-pore, or C-terminus regions.

It has been noted that approximately 10% of genotype positive LQT patients have more than 1 pathogenic variant in >=1 gene. Biallelic pathogenic variants or digenic pathogenic variants appear to be generally associated with a more severe phenotype with longer QTc interval and a higher incidence of cardiac events.

| Gene | *SCN5A* |
| --- | --- |
| OMIM gene number | 600163 |
| Referral indication | Long QT Syndrome (LQTS) |
| Disease grouping | Familial Long QT Syndrome |
| **Disease name** | ***SCN5A*-related long QT Syndrome** |
| MONDO ID | 0011377 |
| Gene disease validity (ClinGen) | DEFINITIVE |
| Inheritance | Autosomal dominant |
| Allelic requirement | Monoallelic autosomal |
| Inheritance modifiers | Typified by incomplete penetrance |
| Disease-associated variant consequence | Altered gene product sequence |
| Variant classes reported with evidence of pathogenicity | Missense; inframe insertion; inframe deletion |
| Restricted repertoire of pathogenic variants | NA |
| PMIDs | 7889574; NBK1129; 29806494; 29798782; 31983240; 11463728; 26320108; 15840476; 19716085; 21185501; 9927399; 22995932; 29650123; 27566755; 32893267; 24932360 |

**Altered gene product sequence** of *SCN5A* causes long QT syndrome. The likely disease mechanism is gain of function.

Over 200 pathogenic missense variants and in-frame deletions or insertions have been reported. The sodium current mediated by Nav1.5 consists of peak and late components (INa-P and INa-L). It is thought that gain-of-function *SCN5A* pathogenic variants lead to enhanced INa-P and INa-L, which can trigger life-threating arrhythmias.

*SCN5A-*related LQTS accounts for 5-10% of LQTS cases (PMID:27566755).

Rare missense variants are estimated to occur in around 2% of healthy White and 5% of healthy non-White subjects so collectively missense variants are not rare in the healthy population.

*SCN5A-*related LQTS can present with specific features: patients may have marked resting bradycardia, QT interval prolongation more pronounced during slow heart rate (which might explain why arrhythmic events occur more frequently at rest), a first cardiac event that is lethal, and onset after puberty (PMID: 29798782).

It has been noted that approximately 10% of genotype positive LQT patients have more than 1 mutation in >=1 gene. Biallelic pathogenic variants or digenic pathogenic variants appear to be generally associated with a more severe phenotype with longer QTc interval and a higher incidence of cardiac events.

Note: loss of function variants in *SCN5A* are associated with Brugada syndrome and individual variants can have hybrid loss of function and gain of function effects causing a mixed phenotype.

| Gene | *CALM1* |
| --- | --- |
| OMIM gene number | 114180 |
| Referral indication | Long QT Syndrome (LQTS) |
| Disease grouping | Long QT syndrome with an atypical presentation |
| **Disease name** | ***CALM1*-related long QT syndrome** |
| MONDO ID | 0014548 |
| Gene disease validity (ClinGen) | DEFINITIVE |
| Inheritance | Autosomal dominant |
| Allelic requirement | Monoallelic autosomal |
| Inheritance modifiers | Typically *de novo* |
| Disease-associated variant consequence | Altered gene product sequence |
| Variant classes reported with evidence of pathogenicity | Missense |
| Restricted repertoire of pathogenic variants | NA |
| PMIDs | 23388215; 26969752; 31983240; 31170290 |

*CALM1-*related LQTS is caused by an **altered gene product sequence.** Reduction in the calcium dependent inactivation of the calcium channel Ca_V_1.2 results in increased inward calcium channel current (ICaL) and repolarization delay. The disease mechanism is likely a dominant negative effect.

The three *CALM* genes encode an identical 149aa protein, calmodulin. The *CALM1* gene is located on chromosome 14, *CALM2* on chromosome 2 and *CALM3* on chromosome 19. Calmodulin protein is involved in many calcium-dependent intracellular processes. All three *CALM* genes have been classified as Definitive for LQTS and Moderate for CPVT, certain variants are only associated with one or other phenotype, whereas others are associated with a mixed or variable phenotypes.

All variants with evidence of pathogenicity identified in the *CALM* genes are missense: at least 35 distinct missense variants have been identified and reported in the International Calmodulinopathy Registry including 11 (31%) in CALM1, 16 (46%) in CALM2, and 8 (23%) in CALM3. The majority of variants across the *CALM* genes affect amino acid residues in the EF-hand Ca2+ binding loop III and IV. Some variants have additional evidence in that the same substitution has been seen in paralogous *CALM* genes, or that the same substitution has been seen in a related phenotype (LQTS). Most variants are unique, but 9 were present in more than one index case and among these, three (p.Asn98Ser, p.Asp130Gly, and p.Phe142Leu, identified in 10, 5, and 4 families, respectively.) appear to be recurrent. While variants p.Asp130Gly and p.Phe142Leu have always been reported as associated with the LQTS phenotype, the variant p.Asn98Ser has phenotypic variability, including LQTS, CPVT, idiopathic VF and sudden death. The majority of *CALM* variants are *de novo*.

Pathogenic variants in the *CALM* genes have been associated with: presentation in infancy or early childhood (up to 5 years); marked sinus bradycardia or atrioventricular block and QT prolongation; and, predominantly with *CALM1* variants, a mild-to-severe neurological impairment, including seizures, development delay, motor and/or cognitive disability.

The phenotype most frequently shown by patients with *CALM* variants is LQTS, but some patients display other phenotypes including CPVT, idiopathic VF and sudden unexplained death. Some variants have been associated with both LQTS and CPVT, however Crotti et al (PMID: 31170290) report that "despite the relatively small numbers of cases, a significant association (P = 0.001) was observed between location of mutation and phenotype (Supplementary material online, Figure S1). Indeed, a pathogenic variant in EF-hand IV Ca2+ binding loop was found in the majority (17/32, 53%) of CALM-LQTS index cases but in only one of the nine CALM-CPVTs (11%). Conversely, variants identified in CALM-CPVT index cases were mostly located either in EF-hand III (n = 5, 56%), or in the inter-EF hand I-II linker (n = 3, 33%)."

| Gene | *CALM2* |
| --- | --- |
| OMIM gene number | 114182 |
| Referral indication | Long QT Syndrome (LQTS) |
| Disease grouping | Long QT syndrome with an atypical presentation |
| **Disease name** | ***CALM2*-related long QT syndrome** |
| MONDO ID | 0014550 |
| Gene disease validity (ClinGen) | DEFINITIVE |
| Inheritance | Autosomal dominant |
| Allelic requirement | Monoallelic autosomal |
| Inheritance modifiers | Typically *de novo* |
| Disease-associated variant consequence | Altered gene product sequence |
| Variant classes reported with evidence of pathogenicity | Missense |
| Restricted repertoire of pathogenic variants | NA |
| PMIDs | 23388215; 26969752; 31983240; 31170290; 27765793 |

*CALM2-*related LQTS is caused by an **altered gene product sequence** which reduces the calcium dependent inactivation of the calcium channel CaV1.2, resulting in increased inward calcium channel current (ICaL) and repolarization delay. The disease mechanism is likely a dominant negative effect.

The three *CALM* genes encode an identical 149aa protein, calmodulin. The *CALM1* gene is located on chromosome 14, *CALM2* on chromosome 2 and *CALM3* on chromosome 19. Calmodulin protein is involved in many calcium-dependent intracellular processes. All three *CALM* genes have been classified as Definitive for LQTS and Moderate for CPVT, certain variants are only associated with one or other phenotype, whereas others are associated with a mixed or variable phenotypes.

All variants with evidence of pathogenicity identified in the *CALM* genes are missense: at least 35 distinct missense variants have been identified and reported in the International Calmodulinopathy Registry including 11 (31%) in CALM1, 16 (46%) in CALM2, and 8 (23%) in CALM3. The majority of variants across the CALM genes affect amino acid residues in the EF-hand Ca2+ binding loop III and IV. Some variants have additional evidence in that the same substitution has been seen in paralogous *CALM* genes, or that the same substitution has been seen in a related phenotype (LQTS). Most variants are unique, but 9 were present in more than one index case and among these, three (p.Asn98Ser, p.Asp130Gly, and p.Phe142Leu, identified in 10, 5, and 4 families, respectively.) appear to be recurrent. While variants p.Asp130Gly and p.Phe142Leu have always been reported as associated with the LQTS phenotype, the variant p.Asn98Ser has phenotypic variability, including LQTS, CPVT, idiopathic VF and sudden death. The majority of *CALM* variants are *de novo*.

Pathogenic variants in the *CALM* genes have been associated with: presentation in infancy or early childhood (up to 5 years); marked sinus bradycardia or atrioventricular block and QT prolongation; and, predominantly with *CALM1* variants, a mild-to-severe neurological impairment, including seizures, development delay, motor and/or cognitive disability.

The phenotype most frequently shown by patients with *CALM* variants is LQTS, but some patients display other phenotypes including CPVT, idiopathic VF and sudden unexplained death. Some variants have been associated with both LQTS and CPVT, however Crotti et al (PMID: 31170290) report that "despite the relatively small numbers of cases, a significant association (P = 0.001) was observed between location of mutation and phenotype (Supplementary material online, Figure S1). Indeed, a pathogenic variant in EF-hand IV Ca2+ binding loop was found in the majority (17/32, 53%) of CALM-LQTS index cases but in only one of the nine CALM-CPVTs (11%). Conversely, variants identified in CALM-CPVT index cases were mostly located either in EF-hand III (n = 5, 56%), or in the inter-EF hand I-II linker (n = 3, 33%)."

| Gene | *CALM3* |
| --- | --- |
| OMIM gene number | 114183 |
| Referral indication | Long QT Syndrome (LQTS) |
| Disease grouping | Long QT syndrome with an atypical presentation |
| **Disease name** | ***CALM3*-related long QT syndrome** |
| MONDO ID | 0019171 |
| Gene disease validity (ClinGen) | DEFINITIVE |
| Inheritance | Autosomal dominant |
| Allelic requirement | Monoallelic autosomal |
| Inheritance modifiers | Typically *de novo* |
| Disease-associated variant consequence | Altered gene product sequence |
| Variant classes reported with evidence of pathogenicity | Missense |
| Restricted repertoire of pathogenic variants | NA |
| PMIDs | 23388215; 26969752; 31983240; 31170290 |

For LQTS, *CALM3* variants lead to an **altered gene product sequence**. The disease mechanism is likely a dominant negative effect which reduces the calcium dependent inactivation of the calcium channel CaV1.2, resulting in increased inward calcium channel current (ICaL) and repolarization delay.

The three *CALM* genes encode an identical 149aa protein, calmodulin. The *CALM1* gene is located on chromosome 14, *CALM2* on chromosome 2 and *CALM3* on chromosome 19. Calmodulin protein is involved in many calcium-dependent intracellular processes. All three *CALM* genes have been classified as Definitive for LQTS and Moderate for CPVT, certain variants are only associated with one or other phenotype, whereas others are associated with a mixed or variable phenotypes.

All variants with evidence of pathogenicity identified in the *CALM* genes are missense: at least 35 distinct missense variants have been identified and reported in the International Calmodulinopathy Registry including 11 (31%) in CALM1, 16 (46%) in CALM2, and 8 (23%) in CALM3. The majority of variants across the *CALM* genes affect amino acid residues in the EF-hand Ca2+ binding loop III and IV. Some variants have additional evidence in that the same substitution has been seen in paralogous *CALM* genes, or that the same substitution has been seen in a related phenotype (LQTS). Most variants are unique, but 9 were present in more than one index case and among these, three (p.Asn98Ser, p.Asp130Gly, and p.Phe142Leu, identified in 10, 5, and 4 families, respectively.) appear to be recurrent. While p.Asp130Gly and p.Phe142Leu have always been reported as associated with the LQTS phenotype, the p.Asn98Ser has phenotypic variability, including LQTS, CPVT, idiopathic VF and sudden death. The majority of *CALM* variants are *de novo*.

Pathogenic variants in the *CALM* genes have been associated with: presentation in infancy or early childhood (up to 5 years); marked sinus bradycardia or atrioventricular block and QT prolongation; and, predominantly with *CALM1* variants, a mild-to-severe neurological impairment, including seizures, development delay, motor and/or cognitive disability.

The phenotype most frequently shown by patients with *CALM* variants is LQTS, but some patients display other phenotypes including CPVT, idiopathic VF and sudden unexplained death. Some variants have been associated with both LQTS and CPVT, however Crotti et al (PMID: 31170290) report that "despite the relatively small numbers of cases, a significant association (P = 0.001) was observed between location of mutation and phenotype (Supplementary material online, Figure S1). Indeed, a pathogenic variant in EF-hand IV Ca2+ binding loop was found in the majority (17/32, 53%) of CALM-LQTS index cases but in only one of the nine CALM-CPVTs (11%). Conversely, variants identified in CALM-CPVT index cases were mostly located either in EF-hand III (n = 5, 56%), or in the inter-EF hand I-II linker (n = 3, 33%)."

| Gene | *TRDN* |
| --- | --- |
| OMIM gene number | 603283 |
| Referral indication | Long QT Syndrome (LQTS) |
| Disease grouping | Long QT syndrome with an atypical presentation |
| **Disease name** | ***TRDN*-related long QT syndrome** |
| MONDO ID | 0019171 |
| Gene disease validity (ClinGen) | DEFINITIVE |
| Inheritance | Autosomal recessive |
| Allelic requirement | Biallelic autosomal |
| Inheritance modifiers | NA |
| Disease-associated variant consequence | Absent gene product level |
| Variant classes reported with evidence of pathogenicity | Frameshift variant NMD triggering |
| Restricted repertoire of pathogenic variants | NA |
| PMIDs | 31983240; 25922419; 26768964; 22422768; 26200674; 30479949; 30649896 |

*TRDN-*related LQTS is due to an **absent gene product level**. The disease mechanism is loss of function (LoF).

***TRDN* is rarely associated with LQTS with an atypical presentation.** Following identification of a homozygous frameshift variant in *TRDN*, 4 other patients with homozygous or compound heterozygous frameshift variants were identified in a cohort of 33 unrelated genotype-negative LQTS patients.

The atypical LQTS phenotype described: extensive T-wave inversions in precordial leads V1 through V4, persistent or transient QT prolongation and severe disease expression of exercise-induced cardiac arrest in early childhood (≤3 years of age).

Biallelic loss of function variants in *TRDN* have also been associated with catecholaminergic polymorphic ventricular tachycardia (CPVT). As patients can have overlapping phenotypes, there is a term “triadin knockout syndrome”. The Triadin Knock Out Syndrome (TKOS) registry had 21 patients in 2019. All were homozygous or compound heterozygous for *TRDN* variants. The majority were frameshift or nonsense. There were a small number of missense or splice altering variants.

*TRDN* is expressed in both cardiac and skeletal muscle and undergoes extensive alternative splicing to produce several isoforms. Only biallelic LoF variants affecting cardiac isoforms should be considered as pathogenic for LQTS.

| Gene | *KCNQ1* |
| --- | --- |
| OMIM gene number | 607542 |
| Referral indication | Long QT Syndrome (LQTS) |
| Disease grouping | Syndrome with QT prolongation and cardiac arrhythmias |
| **Disease name** | ***KCNQ1-* related Jervell and Lange-Nielsen Syndrome** |
| MONDO ID | 0024540 |
| Gene disease validity (ClinGen) | DEFINITIVE |
| Inheritance | Autosomal recessive |
| Allelic requirement | Biallelic autosomal |
| Inheritance modifiers | NA |
| Disease-associated variant consequence | Absent gene product; altered gene product sequence |
| Variant classes reported with evidence of pathogenicity | Splice acceptor; splice donor; frameshift variant NMD triggering; stop gained variant NMD triggering; missense; structural variants (whole exon deletions; complex rearrangements) |
| Restricted repertoire of pathogenic variants | NA |
| PMIDs | 31983240; 9020846; 23392653; 23591039; 27041150; 27868350; 23392653; 11226272; 25187895 |

Biallelic pathogenic variants in *KCNQ1* cause JLNS due to **absent gene product level or altered gene product sequence.**

Both homozygous and compound heterozygous pathogenic variants in *KCNQ1* have been reported in JLNS. Nonsense, frameshift, splice site, whole exon deletions and missense variants have been reported as pathogenic.

Bhuiyan and Wilde (PMID: 23591039) compared two groups of patients with homozygous variants in *KCNQ1*. Those patients where there was residual I_Ks_ (slowly activating delayed rectifier potassium channel/current) even as little as 10%, had QT prolongation but no hearing loss (autosomal recessive LQTS), whereas those patients with complete absence of I_Ks_ had both QT prolongation and hearing loss (JLNS)*.*

They concluded that homozygous or compound heterozygous nonsense, frameshift or exon skipping variants resulting in 100% loss of I_Ks_ would result in JLNS. In addition, biallelic missense mutations that lead to a protein product that does not traffic to the cell membrane (or is subject to nonsense mediated decay) will lead to JLNS.

Note: heterozygous variants in *KCNQ1* lead to dominant LQTS.

| Gene | *KCNE1* |
| --- | --- |
| OMIM gene number | 176261 |
| Referral indication | Long QT Syndrome (LQTS) |
| Disease grouping | Syndromic QT prolongation and cardiac arrhythmias |
| **Disease name** | ***KCNE1*-related Jervell and Lange-Nielsen Syndrome 2** |
| MONDO ID | 0012871 |
| Gene disease validity (ClinGen) | MODERATE |
| Inheritance | Autosomal recessive |
| Allelic requirement | Biallelic autosomal |
| Inheritance modifiers | NA |
| Disease-associated variant consequence | Altered gene product sequence |
| Variant classes reported with evidence of pathogenicity | Missense; inframe insertion; inframe deletion, stop gained NMD escaping |
| Restricted repertoire of pathogenic variants | NA |
| PMIDs | 30461122; 9354783; NBK1405; 31941373; 9445165; 9328483; 10973849; 19716085; 16461811 |

*KCNE1-*related JLNS is due to **altered gene product sequence.** Inheritance is autosomal recessive. Homozygous or compound heterozygous *KCNE1* variants cause JLNS through loss of function (PMID: 9354783). The majority of JLNS patients have biallelic *KCNQ1* variants (JLNS1), only a minority have JLNS2 caused by biallelic *KCNE1* variants (PMID: 16461811).

The KCNE1 protein (previous names LQT5, mink, IsK) functions as a regulatory subunit of KCNQ1. It has 7 annotated exons but only one is protein coding (PMID: 30461122).

A small number of variants have been reported, mainly missense and inframe indels. In more recent studies, 4 nonsense variants have also been reported (PMID: 30461122; PMID: 31941373). However, in one of these studies (PMID: 30461122), patients were ascertained for deafness and QT prolongation is either variable or not reported in some family members. The authors did not carry out functional studies to determine the effect of these variants on mRNA but suggest that given there is only one coding exon, these nonsense variants would be unlikely to result in nonsense mediated decay.

The phenotype of *KCNE1-*related JLNS2 appears to be milder than JLNS1 (caused by biallelic *KCNQ1* variants) (PMID: 16461811; PMID: 31941373)

Heterozygous variants in *KCNE1* have been associated with LQT syndrome without deafness. However, Roberts et al observed a low ECG penetrance in these individuals with the majority not manifesting clinically (PMID: 31941373).

ClinGen classified the gene disease association, *KCNE1* and JLNS2, as Moderate stating that although there was a large amount of experimental evidence and no contradictory evidence, more case level evidence was required to reach a Definitive classification.

| Gene | *KCNJ2* |
| --- | --- |
| OMIM gene number | 600681 |
| Referral indication | Long QT Syndrome (LQTS) |
| Disease grouping | Syndromic QT prolongation and cardiac arrhythmias |
| **Disease name** | ***KCNJ2*-related Andersen Tawil Syndrome** |
| MONDO ID | 0008222 |
| Gene disease validity (ClinGen) | DEFINITIVE |
| Inheritance | Autosomal dominant |
| Allelic requirement | Monoallelic autosomal |
| Inheritance modifiers | NA |
| Disease-associated variant consequence | Altered gene product sequence |
| Variant classes reported with evidence of pathogenicity | Missense; inframe insertion; inframe deletion; stop gained NMD escaping |
| Restricted repertoire of pathogenic variants | NA |
| PMIDs | 31983240; 32947483; 32843460; NBK1264; 32299589; 21493816; 16217063; 32499698; 11371347; 12163457 |

*KCNJ2* pathogenic variants cause Andersen Tawil syndrome (ATS) through **altered gene product sequence.** Most of the pathogenic variants cause Kir2.1 (inwardly-rectifying potassium channel) loss-of-function, either via trafficking or gating defects.

Kir2.1, encoded by *KCNJ2*, contributes a major component of the cardiac action potential repolarization phase. Pathogenic variants of *KCNJ2* gene account for 60-70% of clinical ATS cases, termed type-1 ATS.

The cardiac manifestation includes QT-U abnormalities but not typical QT prolongation. Ventricular arrhythmias also differ from typical LQTS with frequent premature ventricular complexes and polymorphic non-sustained ventricular tachycardia but only rarely torsades de pointes. Extracardiac manifestations include dysmorphic features and periodic paralysis with hypo- and hyperkalemic episodes in some patients.

The majority of pathogenic variants in *KCNJ2* are missense changes. A small number of in-frame deletions, insertions and stop gained variants predicted to escape nonsense mediated decay (NMD) have also been reported in this single-exon gene.

The p.Arg218Trp pathogenic variant is considered a potential mutational hot spot [Davies et al 2005 PMID: 16217063].

Penetrance in families appears high, with estimates of ≥80%

| Gene | *CACNA1C* |
| --- | --- |
| OMIM gene number | 114205 |
| Referral indication | Long QT Syndrome (LQTS) |
| Disease grouping | Syndromic QT prolongation and cardiac arrhythmias |
| **Disease name** | ***CACNA1C*-related Timothy syndrome** |
| MONDO ID | 0010979 |
| Gene disease validity (ClinGen) | DEFINITIVE |
| Inheritance | Autosomal dominant |
| Allelic requirement | Monoallelic autosomal |
| Inheritance modifiers | Typically *de novo* |
| Disease-associated variant consequence | Altered gene product sequence |
| Variant classes reported with evidence of pathogenicity | Missense |
| Restricted repertoire of pathogenic variants | NA |
| PMIDs | 28211989; 25633834; NBK1403; 15863612; 15454078; 22106044; 26253506; 24728418; 31983240 |

*CACNA1C-*related Timothy syndrome is caused by variants leading to **altered gene product sequence**. A recurrent, ***de novo*, missense variant** in *CACNA1C* was described in 13 Timothy syndrome patients, p.Gly406Arg in **exon 8A** (PMID 15454078; PMID: 15863612). The disease mechanism appears to be gain-of-function through failed channel inactivation.

Classic Timothy syndrome (TS1) is a very rare multisystem disorder characterized by marked QT prolongation, syndactyly, immune deficiency, seizures, congenital heart defects, cognitive abnormalities, learning difficulties, and intermittent hypoglycaemia (PMID: 28211989).

*CACNA1C* has a complex genomic structure that undergoes extensive alternative splicing. Splawski et al identified 2 patients with ***de novo* missense variants** in **exon 8** of an alternate splice form (p.Gly406Arg, analogous to the exon 8a variant, and p.Gly402Ser). This splice form represents 80% of all cardiac mRNAs. The patients were described as having atypical Timothy syndrome (TS2), presenting with a more severe cardiac phenotype and without syndactyly (PMID: 15863612; PMID: 25633834).

Other missense variants in *CACNA1C* have been reported in association with isolated LQTS (PMID: 26253506; PMID: 25633834; PMID:24728418). However as of 2020, the ClinGen Cardiovascular Domain Working Group have classified the strength of evidence supporting an association between *CACNA1C* and LQTS as moderate (PMID 31983240).

##### SHORT QT SYNDROME (SQTS)

| Gene | *KCNH2* |
| --- | --- |
| OMIM gene number | 609620 |
| Referral indication | Short QT Syndrome (SQTS) |
| Disease grouping | Classic SQTS |
| **Disease name** | ***KCNH2*-related SQTS** |
| MONDO ID | 0012312 |
| Gene disease validity (ClinGen) | DEFINITIVE |
| Inheritance | Autosomal dominant |
| Allelic requirement | Monoallelic autosomal |
| Inheritance modifiers | NA |
| Disease-associated variant consequence | Altered gene product sequence |
| Variant classes reported with evidence of pathogenicity | Missense |
| Restricted repertoire of pathogenic variants | NA |
| PMIDs | 14676148, 19340359, 25335996, 21130771, 29876509, 29016797, 28491588, 30571592, 15828882, 18692916, 25974115, 21310316, 31072576, 7736582, 7889573, 29574456, 30582453, 9547387, 15673388, 29759541, 31049424, 30175559, 19088443, 30496390, 30947366 |

SQTS is a rare (< 1/10,000) AD inherited arrhythmia syndrome associated with atrial fibrillation, ventricular arrhythmia, and risk of sudden cardiac arrest. Diagnosis is based on a diagnostic scorecard (PMID: 21310316), similar to LQTS. Approximately 20% of diagnosed cases will have a genetic cause, and *KCNH2* is the most common identified gene.

It is noteworthy that of the 18 probands with SQTS in whom KCNH2 variants have been identified, 13 had one of 2 variants; 7 with p.Thr618Ile variant (ClinVar Variation ID# 67297) and 6 with p.Asn588Lys (ClinVar Variation ID# 14436; NM_172056.2). There is high penetrance for these recurrent variants.

Experimental evidence derived from non-patient cells, human-induced pluripotent stem cell-derived cells and a rabbit animal model (PMID: 30496390) all support this gene’s relationship with SQTS, with a gain-of-function mechanism. These experimental studies demonstrate that genetic variants identified in SQTS patients lead to potassium current perturbations concordant with SQTS phenotype and shortening of the QT interval.

Note loss-of-function variants in *KCNH2* are associated with LQTS.

| Gene | *KCNQ1* |
| --- | --- |
| OMIM gene number | 609621 |
| Referral indication | Short QT Syndrome (SQTS) |
| Disease grouping | Classic SQTS |
| **Disease name** | ***KCNQ1*-related SQTS** |
| MONDO ID | 0012313 |
| Gene disease validity (ClinGen) | STRONG |
| Inheritance | Autosomal dominant |
| Allelic requirement | Monoallelic autosomal |
| Inheritance modifiers | NA |
| Disease-associated variant consequence | Altered gene product sequence |
| Variant classes reported with evidence of pathogenicity | Missense |
| Restricted repertoire of pathogenic variants | Almost all evidence derived from p.V141M; p.V307L |
| PMIDs | 16109388, 28491547, 15159330, 24380499, 25974115, 26168993, 26346102, 26279191, 28491751, 8900283, 8528244, 29213224 |

Almost all evidence for *KCNQ1* as a cause of SQTS is derived from a single variant (p.Val141Met) identified in 9 unrelated probands. All 9 cases presented with severe bradycardia in-utero or at birth and in 6 atrial fibrillation was also reported. In one case complete atrioventricular block was documented. (PMIDs: 24818999, 26279191, 16109388, 24380499, 25974115, 28491547).

Five other variants have been reported for SQTS, but the phenotypic features of SQTS for 4 of these have been unclear or functional data supporting the mechanism of disease lacking in the published manuscripts. For the fifth additionally reported variant, 70-year-old patient presented with ‘idiopathic VF” and a short QT interval, a *KCNQ1* variant, p.Val307Leu, was reported as the likely genetic culprit, with in vitro studies reporting a gain-of-function effect of the mutant (Bellocq et al, Circ 2004).

Functional characterisation in cell models confirms a gain of function mechanism for Val141Met and Val307Leu.

Four cases of Val141Met are reported as *de novo* without noted confirmed paternity. Importantly, in none of the p.Val141Met cases was cardiac arrest or sudden cardiac death described.

Note loss-of-function variants in *KCNQ1* are associated with LQTS.

| Gene | *SLC4A3* |
| --- | --- |
| OMIM gene number | 106195 |
| Referral indication | Short QT Syndrome (SQTS) |
| Disease grouping | Classic SQTS |
| **Disease name** | ***SLC4A3*-related SQTS** |
| MONDO ID | 0000453 |
| Gene disease validity (ClinGen) | MODERATE |
| Inheritance | Autosomal dominant |
| Allelic requirement | Monoallelic autosomal |
| Inheritance modifiers | NA |
| Disease-associated variant consequence | Altered gene product sequence |
| Variant classes reported with evidence of pathogenicity | Missense |
| Restricted repertoire of pathogenic variants | p.Arg370His |
| PMIDs | 29167417 |

*SLC4A3* encodes a plasma membrane anion exchange protein. Genetic evidence supporting *SLC4A3* as a SQTS-causing gene is derived from a single publication in which exome sequencing was performed in 2 families, including one large pedigree (PMID 29167417). The same rare genetic variant (p.Arg370His, c.1109G>A) was identified in both families, suggesting they are possibly distantly related.

Experimental evidence from in vitro and zebrafish models suggests reduced membrane localization of the mutated protein leads to intracellular alkalinization and shortening of the cardiomyocyte action potential duration.

The genetic evidence, including the unbiased gene discovery approach of whole exome sequencing and segregation of the identified genetic variant with a large number of affected individuals within the presented pedigree, was considered relatively strong by the ClinGen GCEP. However, lack of other publications supporting this gene-disease relationship led to a score in the "Moderate" range (for gene-disease validity). Further information on ClinGen gene-disease validity can be found here: https://search.clinicalgenome.org/kb/genes/HGNC:11029

| Gene | *KCNJ2* |
| --- | --- |
| OMIM gene number | 609622 |
| Referral indication | Short QT Syndrome (SQTS) |
| Disease grouping | Classic SQTS |
| **Disease name** | ***KCNJ2*-related SQTS** |
| MONDO ID | 0012314 |
| Gene disease validity (ClinGen) | MODERATE |
| Inheritance | Autosomal dominant |
| Allelic requirement | Monoallelic autosomal |
| Inheritance modifiers | NA |
| Disease-associated variant consequence | Altered gene product sequence |
| Variant classes reported with evidence of pathogenicity | Missense |
| Restricted repertoire of pathogenic variants | NA |
| PMIDs | 15761194, 23440193, 2479485, 29615871, 22155372, 11410627, 19285083, 24794859, 19710529 |

Genetic variants in KCNJ2 have been identified in 6 patients from 5 families with unique variants, including at least 2 probands with a de-novo variant (paternity not confirmed). Experimental evidence demonstrated these variants lead to gain-of-function of the late repolarizing, KCNJ2-encoded Ik1 current in the heart, and abbreviation of the action potential duration. The ClinGen GCEP reviewing gene-disease validity considered these data sufficient for classifying the gene-disease relationship of KCNJ2 as “Moderate” but, in the absence of segregation or case-control data, the genetic evidence was not sufficient for a stronger classification. Further information on ClinGen gene-disease validity can be found here: https://search.clinicalgenome.org/kb/genes/HGNC:6263

| Gene | *SLC22A5* |
| --- | --- |
| OMIM gene number | 212140 |
| Referral indication | Short QT Syndrome (SQTS) |
| Disease grouping | Syndrome including shortened QT and cardiac arrhythmias |
| **Disease name** | ***SLC22A5*-related primary systemic carnitine deficiency** |
| MONDO ID | 0008919 |
| Gene disease validity (ClinGen) | DEFINITIVE |
| Inheritance | Autosomal recessive |
| Allelic requirement | Biallelic autosomal |
| Inheritance modifiers | NA |
| Disease-associated variant consequence | Altered gene product sequence |
| Variant classes reported with evidence of pathogenicity | Missense |
| Restricted repertoire of pathogenic variants | NA |
| PMIDs | 26190315, 31472821, 3009296 |

Variants in SLC22A5 cause autosomal recessive primary systemic carnitine deficiency (PSCD), a syndrome characterized by hypoketotic hypoglycemia, hyperammonemia, liver dysfunction, hypotonia and cardiomyopathy (PMID 26190315).

Homozygote or compound heterozygote variants have been identified in unexplained sudden cardiac death or resuscitated cardiac arrest cases without overt extra-cardiac manifestations (PMIDs: 31472821, 3009296). Furthermore, a short QT interval has been demonstrated in a carnitine deficient mouse model (31472821) as well as in patients with PSCD (31472821, 3009296). Importantly, however, the QT interval in these patients returns to normal with carnitine supplementation treatment.

Information on ClinGen gene-disease validity conclusion: the ClinGen GCEP conclude that PSCD is a metabolic and reversible SQTS-mimic. While there is no robust evidence of a relationship between SLC22A5 and true SQTS [ClinGen “disputed” classification], there is "definitive" evidence for a relationship between SLC22A5 and PSCD which may justify testing in individuals being investigated for possible SQTS. Further information on ClinGen gene-disease validity can be found here: https://search.clinicalgenome.org/kb/genes/HGNC:10969

### CARDIOMYOPATHY

##### ARRHYTHMOGENIC RIGHT VENTRICULAR CARDIOMYOPATHY

| Gene | *DSC2* |
| --- | --- |
| OMIM gene number | 125645 |
| Referral indication | Arrhythmogenic right ventricular cardiomyopathy (ARVC) |
| Disease grouping | Familial isolated arrhythmogenic cardiomyopathy |
| **Disease name** | ***DSC2*-related ARVC** |
| MONDO ID | 0012506 |
| Gene disease validity (ClinGen) | DEFINITIVE |
| Inheritance | Autosomal dominant; Autosomal recessive |
| Allelic requirement | Monoallelic autosomal; Biallelic autosomal |
| Inheritance modifiers | Typified by incomplete penetrance |
| Disease-associated variant consequence | Decreased gene product level; altered gene product sequence |
| Variant classes reported with evidence of pathogenicity | Splice region; splice acceptor; splice donor; frameshift; stop gained; missense; inframe insertion; inframe deletion |
| Restricted repertoire of pathogenic variants | NA |
| PMIDs | 31028357; 23911551; NBK1131; 21636032; 33831308; 26310507; 23863954; 24793512; 24070718; 34400560; 17033975; 17963498; 17186466; 20031616; 19863551; 31402444 |

*DSC2*-related ARVC is due to **decreased gene product level or altered gene product sequence** due to a variety of mechanisms (e.g. null alleles, trafficking defects, impaired proteolytic processing, absence of or impaired protein-protein interactions) (PMID: 31028357; 23911551; NBK1131).

Loss of function is the likely disease mechanism. ClinGen found there was some evidence to support haploinsufficiency as a mechanism. <https://search.clinicalgenome.org/kb/gene-dosage/HGNC:3036>

Autosomal dominant inheritance with incomplete penetrance is the most common mode of transmission (PMID: 21636032; 33831308). Homozygous and compound heterozygous variants have also been described in association with ARVC with or without cutaneous features (PMID: 26310507; 23863954; 24793512; 24070718; 34400560). In some cases, these appear to reflect autosomal recessive inheritance (PMID 24793512; 23863954, 33831308). Instances of digenic inheritance have been identified with *DSC2* variants along with other desmosomal gene pathogenic variants (PMID: 24070718).

A number of *DSC2* variants have been reported in the literature including nonsense, frameshift, splice, missense and inframe insertions and deletions (NBK1131; 17033975; 17963498; 17186466; 20031616; 19863551; 31402444).

*DSC2*-related ARVC appears to be characterised by an increased risk of biventricular involvement and heart failure when compared to PKP2-related ARVC (PMID: 34400560).

| Gene | *DSG2* |
| --- | --- |
| OMIM gene number | 125671 |
| Referral indication | Arrhythmogenic right ventricular cardiomyopathy (ARVC) |
| Disease grouping | Familial isolated arrhythmogenic cardiomyopathy |
| **Disease name** | ***DSG2*-related ARVC** |
| MONDO ID | 0012434 |
| Gene disease validity (ClinGen) | DEFINITIVE |
| Inheritance | Autosomal dominant; Autosomal recessive |
| Allelic requirement | Monoallelic autosomal; Biallelic autosomal |
| Inheritance modifiers | Typified by incomplete penetrance |
| Disease-associated variant consequence | Decreased gene product level; altered gene product sequence |
| Variant classes reported with evidence of pathogenicity | Splice acceptor; splice donor; frameshift; stop gained; missense; inframe insertion; inframe deletion |
| Restricted repertoire of pathogenic variants | NA |
| PMIDs | 21636032; 33831308; 33917638; 34400560**;** 24070718; 30454721; 25616645; 30790397; 34400560; 16505173; NBK1131; 27532257; 16823493; 27170944 |

*DSG2-*related ARVC is due to **decreased gene product level or altered gene product sequence** due to a variety of mechanisms. Much of the underlying pathogenesis of *DSG2* pathogenic variants is still unknown; it is believed that loss of *DSG2* compromises cell-to-cell adhesion between cardiomyocytes (PMID:26085008; NBK1131). There is also work revealing that desmosomal variants can reduce canonical Wnt signaling and activating Wnt with a GSK3B inhibitor can block disease pathogenesis (PMID 16823493; 27170944).

The usual mode of inheritance is **autosomal dominant characterized by incomplete penetrance** (PMID: 21636032; 33831308). Compound heterozygous and homozygous variants have been described. In some families, heterozygous carriers of these variants were not affected suggesting **autosomal recessive inheritance** (PMID: 33917638; 34400560**;** 24070718; 33831308; 30454721). Patients with >1 variant appear to have a more severe phenotype (PMID: 25616645; PMID: 30790397).

The majority of *DSG2* variants are rare missense variants with unknown significance/unknown mechanism of pathogenicity. In addition, nonsense, frameshift, insertions, deletions, and splice site variants have all been described (PMID: 16505173; NBK1131; 30790397; 27532257; 33917638).

*DSG2-*related ARVC appears to be characterised by an increased risk of biventricular involvement and heart failure when compared to *PKP2-*related ARVC (PMID: 34400560**;** 30790397).

| Gene | *DSP* |
| --- | --- |
| OMIM gene number | 125647 |
| Referral indication | Arrhythmogenic right ventricular cardiomyopathy (ARVC) |
| Disease grouping | Familial isolated arrhythmogenic cardiomyopathy |
| **Disease name** | ***DSP*-related ARVC** |
| MONDO ID | 0011831 |
| Gene disease validity (ClinGen) | DEFINITIVE |
| Inheritance | Autosomal dominant; Autosomal recessive |
| Allelic requirement | Monoallelic autosomal; Biallelic autosomal |
| Inheritance modifiers | Typified by incomplete penetrance |
| Disease-associated variant consequence | Decreased gene product level; altered gene product sequence |
| Variant classes reported with evidence of pathogenicity | Splice donor; splice acceptor; frameshift; stop gained; missense; inframe insertion; inframe deletion |
| Restricted repertoire of pathogenic variants | NA |
| PMIDs | 32372669; 23137101; 21636032; 33831308; 31319917; 20716751; 23810894; 24503780; 27532257; 31514951; 27761164; 21636032; 32808748; 33275305; 11063735; 27761164; 20940358; 22795705; 26604139; 30382575 |

*DSP*-related ARVC is due to **decreased gene product level or altered gene product sequence.**

The disease mechanism is loss of function via haploinsufficiency, dominant negative or both (PMID 32372669; 23137101; 16917092; NBK1131).

*DSP*-related ARVC is inherited in an **autosomal dominant** manner characterized by **incomplete penetrance** (PMID: 21636032; 33831308). However, ***DSP* is associated with multiple phenotypes which are heterogeneous and overlapping** (including DCM, DCM with cutaneous features, ARVC, and Carvajal syndrome) and **autosomal recessive inheritance has been reported**. There does not appear to be distinct mechanisms leading to different phenotypes <https://search.clinicalgenome.org/kb/gene-dosage/HGNC:3052>.

The initial variant description in *DSP* was in Carvajal syndrome characterized by woolly hair, keratoderma and ARVC. In 2000 three families from Ecuador were found to be homozygous for the variant 7901delG in *DSP* which produces a premature stop codon leading to a truncated desmoplakin protein missing the C domain of the tail region (PMID 11063735). Since then both **autosomal dominant** and **autosomal recessive** patterns of inheritance have been described in Carvajal syndrome (PMID: 27761164; 20940358; 22795705; 26604139; 23137101). This was followed by the description of a heterozygous variant in *DSP* in an Italian family with ARVC with co-segregation of the variant with disease. There have been reports of digenic inheritance with other desmosomal pathogenic variants.

In a retrospective multicentre study, curly hair and/or thick skin on the palms or soles (palmoplantar keratoderma) was commonly present in *DSP* patients (54/98, 55%) but not in *PKP2* patients (1/46, 2%) (PMID: 32372669). Maruthappu et al 2019 also describe 38 patients with arrhythmogenic cardiomyopathy who were carriers of a dominant loss-of-function (nonsense or frameshift) variants in *DSP*. Nearly all were found to have curly hair and palmoplantar keratoderma. However, there was one family described where the majority did not demonstrate a curly hair/cutaneous phenotype. The variant in this family was located in a fragment (c.3585–5379, (p.1195–1793)) only included in isoform 1 of *DSP* (it has previously been shown that isoform 2 is the major isoform regulating keratinocyte adhesion (PMID: 30382575).

Both **truncating** (stop gained, frame shift, splice site) and **non-truncating variants** in *DSP* have been reported in the literature associated with ARVC (PMID 31319917; 20716751; 23810894; 24503780; 27532257; 31514951; 27761164; 21636032; 32808748). Pathogenic truncating variants are more common.

Grondin et al 2020 re-evaluated reported missense variants and found an enrichment localizing to the spectrin repeat domain (SRD) in cases vs gnomAD. A similar hot spot location (amino acid residues 250-604) was reported by Kapplinger et al in 2011 (PMID: 32808748; 21636032).

Smith E et al 2020 report that *DSP* variants are associated with a distinct type of cardiomyopathy with a high prevalence of LV inflammation, fibrosis, and systolic dysfunction, and *DSP* cardiomyopathy should be considered in the differential diagnosis for myocarditis and sarcoidosis (PMID: 32372669).

| Gene | *PKP2* |
| --- | --- |
| OMIM gene number | 602861 |
| Referral indication | Arrhythmogenic right ventricular cardiomyopathy (ARVC) |
| Disease grouping | Familial isolated arrhythmogenic cardiomyopathy |
| **Disease name** | ***PKP2*-related ARVC** |
| MONDO ID | 0012180 |
| Gene disease validity (ClinGen) | DEFINITIVE |
| Inheritance | Autosomal dominant, Autosomal recessive |
| Allelic requirement | Monoallelic autosomal |
| Inheritance modifiers | Typified by incomplete penetrance |
| Disease-associated variant consequence | Decreased gene product level; altered gene product sequence |
| Variant classes reported with evidence of pathogenicity | Splice region; splice acceptor; splice donor; stop gained; frameshift; missense; structural variants (deletions; duplications) |
| Restricted repertoire of pathogenic variants | NA |
| PMIDs | 33831308; 21636032; 23736219; 34120153; 30830208; 25616645; 24070718; 17010805; NBK1131; 30619891; 24704780; 28740174; 22781308; 20301310; 17041889 |

*PKP2* pathogenic variants cause ARVC through **decreased gene product level or altered gene product sequence.** *PKP2* encodes plakophilin-2 which is a protein of the desmosome and provides structural and functional integrity to adjacent cells. The disease mechanism in ARVC is loss of function (LoF). Rasmussen et al showed that truncating variants in *PKP2* resulted in PKP2 transcript and protein levels reduced to ≈50% (PMID: 24704780). Cerrone et al showed that loss of *PKP2* in adult myocytes was sufficient to generate an arrhythmogenic cardiomyopathy of right ventricle predominance in mice (PMID: 28740174)

*PKP2* is the major causative gene for ARVC and accounts for 34%-74% of cases (PMID: 20301310).

Inheritance is predominantly **autosomal dominant** characterised by variable expression and **incomplete penetrance** (PMID: 34120153; 21636032; 17010805).

Both recessive and digenic inheritance (with one pathogenic variant in *PKP2* and a second in another desmosomal gene) have been reported (including a recessive cryptic splice variant PMID 17041889) and appear to confer a more severe phenotype (PMID: 30830208; 25616645; 24070718; NBK1131). The expert panel noted instances where *PKP2* LoF variants on both alleles had resulted in neonatal lethality.

There are over 250 *PKP2* variants listed in ClinVar for ARVC (nonsense, frameshift, splice, missense, deletions, duplications, and complex rearrangements (PMID: 30619891; 25616645; 21636032; 34120153).

Dries et al report that *PKP2* truncating variants explain a large proportion of ARVC cases but there is no clear relationship between their transcript position and their likelihood of disease association (PMID: 30619891). Although missense variants are associated with disease and validated with functional studies (PMID 22781308), their mechanism and overall impact in ARVC is not completely understood. The majority of missense variants in ClinVar are classified as variants of uncertain significance.

| Gene | *TMEM43* |
| --- | --- |
| OMIM gene number | 612048 |
| Referral indication | Arrhythmogenic right ventricular cardiomyopathy (ARVC) |
| Disease grouping | Familial isolated arrhythmogenic cardiomyopathy |
| **Disease name** | ***TMEM43*-related ARVC** |
| MONDO ID | 0011459 |
| Gene disease validity (ClinGen) | DEFINITIVE |
| Inheritance | Autosomal dominant |
| Allelic requirement | Monoallelic autosomal |
| Inheritance modifiers | NA |
| Disease-associated variant consequence | Altered gene product sequence |
| Variant classes reported with evidence of pathogenicity | Missense |
| Restricted repertoire of pathogenic variants | S358L |
| PMIDs | 20301310; 18313022; 21214875; 23812740; 24598986; 33831308; 21391237; 29980933; 25343256; 22725725; 32062046 |

*TMEM43-*related ARVC is due to **altered gene product sequence.**

Pathogenic variants in *TMEM43* are a rare cause of ARVC (PMID: 20301310).

The majority of genetic evidence comes from **one founder missense variant, S358L** (PMID 18313022; 21214875; 23812740; 20301310; 24598986; 33831308). This was originally identified in Newfoundland and has subsequently been found in patients from other countries including USA, Germany, and Denmark (PMID: 33831308; 18313022; 23812740). It is reported that “[the variant occurs on] a common haplotype with those from Newfoundland, USA, and Denmark, suggesting that the mutation originated from a common founder. Examination of 40 control chromosomes revealed an estimated age of 1300-1500 years for the mutation, which proves the European origin of the Newfoundland mutation.” (PMID  24598986)

The disease mechanism is largely unknown. There is no evidence currently for haploinsufficiency (<https://search.clinicalgenome.org/kb/gene-dosage/HGNC:28472>).

Although ARVC is known to display incomplete penetrance, this particular founder variant appears to be more penetrant.

*TMEM43*-related ARVC is associated with a high risk of sudden cardiac death and characteristic clinical and electrocardiographic features (PMID: 32062046). Ventricular ectopy on Holter monitoring is commonly seen and can occur early in the natural history (PMID: 22725725).

PMID 21391237 described two patients with *TMEM43* heterozygous missense variants in Emery Dreifuss Muscular Dystrophy Related Myopathy. Other missense variants have been reported but their pathogenicity is debated.

| Gene | *JUP* |
| --- | --- |
| OMIM gene number | 173325 |
| Referral indication | Arrhythmogenic right ventricular cardiomyopathy (ARVC) |
| Disease grouping | Rare familial disorder with ARVC |
| **Disease name** | ***JUP*-related Naxos disease** |
| MONDO ID | 0011017 |
| Gene disease validity (ClinGen) | STRONG |
| Inheritance | Autosomal recessive |
| Allelic requirement | Biallelic autosomal |
| Inheritance modifiers | NA |
| Disease-associated variant consequence | Altered gene product sequence |
| Variant classes reported with evidence of pathogenicity | Frameshift variant NMD escaping, missense, inframe deletion |
| Restricted repertoire of pathogenic variants | NA |
| PMIDs | 10902626; 32966140; 17924338; 25820315 20031617; 25705887; 21673311; 11691526; 28098346; 15851108; 31402444; 20130592; 21320868; 8954745; 8858175 |

*JUP*-related Naxos disease (ARVC, woolly hair and palmoplantar keratoderma) is due to **altered gene product sequence** causing loss of function of *JUP* (PMID: 25705887; 21673311; 11691526; 10902626). *JUP* encodes the protein plakoglobin. Kaplan et al found that in 4 Naxos patients, Connexin43 expression at intercellular junctions was significantly reduced and mutant plakoglobin was expressed but failed to localize normally at intercellular junctions (PMID:15851108)

Inheritance is **autosomal recessive**.

The initial nine patients described ranged in age from 7 to 41 years. Since then, more patients have been discovered carrying the disease with an estimate of 1:1000 in the population of the Greek islands. The disease has also been diagnosed in other countries (PMID: 32966140).

A **homozygous 2bp deletion** in plakoglobin (*JUP*), c.2157delTG, causing a frameshift and premature termination of the protein and expression of a truncated plakoglobin lacking 56 residues from the C terminus was described in 2000 (PMID:10902626). The truncated protein was identified on western blot.

In 2017, a **homozygous missense variant** was described in 7 unrelated French-Canadian individuals. All had typical hair and skin findings; 4/7 had ARVC presenting after 28 years (PMID: 28098346). The effect of this variant in the heterozygous state was not investigated.

Two siblings of consanguineous parents were found to have a **homozygous 3bp deletion** in *JUP* c.901_903delGAG (p.Glu301del). Both had woolly hair and skin findings, only the older sister had ARVC and neither had palmoplantar keratoderma (PMID: 28098346).

In OMIM there have been reports of other types of homozygous variants (nonsense, splice, missense) in *JUP* causing overlapping phenotypes and segregating with disease. Data on biallelic LoF variants are sparse.  In mice, generation of a null mutation of the plakoglobin gene by homologous recombination results in embryonic lethality (PMID: 8954745; 8858175). There are 2 reports in humans who had skin features but no obvious cardiomyopathy (PMID: 20130592; 21320868). In one, *JUP* expression in the skin was absent. Cardiac *JUP* expression was not directly measured to establish the consequence in the heart – it is not known whether variant allele was expressed, degraded, or rescued by alternate splicing.

To note dominant pathogenic variants in *JUP* have also been rarely described in association with ARVC. Asimaki et al reported a dominant variant in *JUP* in a German family with ARVC and no obvious cutaneous abnormalities (PMID: 17924338). Other studies have identified heterozygous missense variants however their pathogenicity is still debated (PMID 25820315; 20031617; 31402444).

##### DILATED CARDIOMYOPATHY

| Gene | *BAG3* |
| --- | --- |
| OMIM gene number | 603883 |
| Referral indication | Dilated Cardiomyopathy (DCM) |
| Disease grouping | Familial Dilated Cardiomyopathy |
| **Disease name** | ***BAG3*-related DCM** |
| MONDO ID | 0013479 |
| Gene disease validity (ClinGen) | DEFINITIVE |
| Inheritance | Autosomal dominant |
| Allelic requirement | Monoallelic autosomal |
| Inheritance modifiers | Typified by incomplete penetrance |
| Disease-associated variant consequence | Decreased gene product level; altered gene product sequence |
| Variant classes reported with evidence of pathogenicity | Missense; stop gained NMD triggering; frameshift; splice acceptor; splice donor; structural variants (whole exon deletions), copy number variation (whole gene deletion) |
| Restricted repertoire of pathogenic variants | NA |
| PMIDs | 23518596; 20884878; 26796036; 21353195; 25008357; 27391596; 30442290; 31983221; 21898660; 20884878; 28737513; 31808029; 32160020 |

Pathogenic variants in *BAG3* cause an estimated 2-4% of familial DCM due to **decreased gene product level** or **altered gene product sequence.** This likely leads to loss of function causing several pathological effects on cardiomyocytes including direct destabilization of the Z-disc, impaired protein homeostasis leading to proteotoxicity and increased susceptibility to apoptosis.

Truncating variants are responsible for most *BAG3*-related DCM cases, the majority of which are nonsense and frameshift, and include several single, multi-exon, and whole gene deletions. ClinGen have concluded there is good evidence for haploinsufficiency. There is not yet evidence for dominant-negative effects.

There is no apparent enrichment for non-truncating mutations in any specific domain, and it should be noted that different missense variants in similar domains can lead to different phenotypes (e.g., p.Pro209Leu leading to myofibrillar myopathy, and p.Arg218Trp leading to DCM).

*BAG3*-related DCM is inherited as an autosomal dominant trait, characterised by incomplete penetrance.

| Gene | *DES* |
| --- | --- |
| OMIM gene number | 604765 |
| Referral indication | Dilated Cardiomyopathy (DCM) |
| Disease grouping | Familial Dilated Cardiomyopathy |
| **Disease name** | ***DES*-related Dilated Cardiomyopathy** |
| MONDO ID | 0011482 |
| Gene disease validity (ClinGen) | DEFINITIVE |
| Inheritance | Autosomal dominant |
| Allelic requirement | Monoallelic autosomal |
| Inheritance modifiers | NA |
| Disease-associated variant consequence | Altered gene product sequence |
| Variant classes reported with evidence of pathogenicity | Missense; splice acceptor variant NMD escaping |
| Restricted repertoire of pathogenic variants | NA |
| PMIDs | 17626518; 17325244; 10430757; 11728149; 17325244; 23300193; 23349452; 26724190; 17626518 |

Pathogenic variants in *DES* cause DCM due to **altered gene product sequence.**

Only missense variants in *DES* have been confidently reported as pathogenic in DCM. There are no reports of truncating variants; *DES* is yet to undergo ClinGen Dosage Haploinsufficiency investigations. *DES* has a pLI of 0.01 in gnomAD (o/e = 0.33 (0.19 - 0.6)).

There is some indication that truncating variants in *DES* are associated with myofibrillar myopathy rather than DCM. There is a report of a splice site variant in a DCM case, however, it causes exon 3 skipping to produce an inframe transcript (PMID: 17626518)

*DES*-related DCM is inherited in an autosomal dominant manner, and disease is generally highly penetrant. *DES* missense variants account for a very small proportion (estimated 2%) of genetically-explained DCM (PMID: 17325244).

| Gene | *DSP* |
| --- | --- |
| OMIM gene number | 615821 |
| Referral indication | Dilated Cardiomyopathy (DCM) |
| Disease grouping | Familial Dilated Cardiomyopathy |
| **Disease name** | ***DSP*-related DCM** |
| MONDO ID | 0005021 |
| Gene disease validity (ClinGen) | DEFINITIVE |
| Inheritance | Autosomal dominant |
| Allelic requirement | Monoallelic autosomal |
| Inheritance modifiers | NA |
| Disease-associated variant consequence | Decreased gene product level; altered gene product sequence |
| Variant classes reported with evidence of pathogenicity | Stop gained NMD triggering; frameshift variant; splice acceptor variant NMD triggering; splice donor variant NMD triggering; missense |
| Restricted repertoire of pathogenic variants | NA |
| PMIDs | 31983221; 31317183; 27532257; 32005173; 24503780; 23022708; 20716751; 32013205 |

Pathogenic variants in *DSP* cause DCM due to **decreased gene product level or altered gene product sequence** due to a variety of mechanisms.

Both missense and truncating mutations have been reported in DCM cases, however, there is significantly more evidence in support of truncating variants being pathogenic.

*DSP*-related DCM is inherited in an autosomal dominant manner, and disease is generally penetrant in families with an increased burden of LV fibrosis and ventricular tachyarrhythmia. Pathogenic *DSP* variants are likely to account for approximately 2-3% of familial DCM cases (PubMed ID 24503780 and 23022708)

Of note *DSP* is also associated ARVC for which *DSP* is one of the most common causes.

| Gene | *FLNC* |
| --- | --- |
| OMIM gene number | 102565 |
| Referral indication | Dilated Cardiomyopathy (DCM) |
| Disease grouping | Familial Dilated Cardiomyopathy |
| **Disease name** | ***FLNC*-related DCM** |
| MONDO ID | 0005021 |
| Gene disease validity (ClinGen) | DEFINITIVE |
| Inheritance | Autosomal dominant |
| Allelic requirement | Monoallelic autosomal |
| Inheritance modifiers | Typified by incomplete penetrance |
| Disease-associated variant consequence | Decreased gene product level |
| Variant classes reported with evidence of pathogenicity | Splice acceptor; splice donor; frameshift variant NMD triggering; stop gained NMD triggering |
| Restricted repertoire of pathogenic variants | NA |
| PMIDs | 27206985; 321126565; 27908349; 30067491; 2843697; 31627847; 27601210; 32150467; 22020047; 32160020; 32154132 |

Pathogenic variants in *FLNC* account for approximately 2-4% of familial DCM due to **decreased gene product level.** The mechanism is likely loss of function, leading to Z-disc disarray and weakened cell-cell adhesion, promoting arrhythmogenesis and fibrosis.

Truncating variants (stop-gained, frameshift and splicing) are responsible for almost all reported cases of *FLNC*-related DCM to date. It is possible that some loss-of-function missense variants could cause DCM and experts commented on their experience of families with missense variants segregating with disease. However, there is limited evidence for this. Xiao et al 2020 report an infant girl presenting with DCM and a paternally inherited missense variant, p.Arg441Ile in *FLNC* identified on exome sequencing. No functional studies were carried out (PMID: 32154132).

The *FLNC*-related DCM phenotype is frequently arrhythmogenic, characterized by a high burden of ventricular arrhythmias and myocardial fibrosis.

*FLNC*-related DCM is inherited as an autosomal dominant trait, penetrance in close relatives of an affected proband is generally high, although not complete, with mean age of onset in the late 4^th^ or early 5^th^ decade, +/- 10-15 years.

| Gene | *LMNA* |
| --- | --- |
| OMIM gene number | 115200 |
| Referral indication | Dilated Cardiomyopathy (DCM) |
| Disease grouping | Familial Dilated Cardiomyopathy |
| **Disease name** | ***LMNA*-related DCM** |
| MONDO ID | 0007269 |
| Gene disease validity (ClinGen) | DEFINITIVE |
| Inheritance | Autosomal dominant |
| Allelic requirement | Monoallelic autosomal |
| Inheritance modifiers | Typified by age-related onset |
| Disease-associated variant consequence | Decreased gene product level; altered gene product sequence |
| Variant classes reported with evidence of pathogenicity | Missense; stop gained NMD triggering; frameshift variant; splice acceptor variant NMD triggering; splice donor variant NMD triggering; structural variants (single and multi-exon deletion) |
| Restricted repertoire of pathogenic variants | NA |
| PMIDs | 28912180; NBK1674; 28912180; 10580070; 10662742; 11897440; 29095976; 18926329; 29367541; 20127487; 12854972; 12920062; 31983221 |

Pathogenic variants in *LMNA* cause DCM due to **decreased gene product level** or **altered gene product sequence,** likely due to a variety of mechanisms. Missense variants are more prevalent, and are responsible for a larger proportion of DCM cases, but truncating variants (nonsense, frameshift, and splice variants) are also associated with disease, most probably through loss of function mechanisms.

*LMNA* has a ClinGen Dosage sensitivity score of 2, indicating there is some evidence for dosage pathogenicity. Non-missense variants are reported to convey a higher risk of life-threatening arrhythmia (31155932).

*LMNA-*related DCM is inherited in an autosomal dominant manner, and disease is generally a highly penetrant and aggressive arrhythmogenic phenotype with high rates of heart failure and sudden cardiac death. *LMNA* missense and truncating variants account for ~5-8% of genetic DCM.

Of note *LMNA* is also associated with several other conditions (the laminopathies) including muscular dystrophies and Hutchinson-Gilford progeria. Some of these conditions are autosomal recessive and some dominant. There is evidence of clustering of variants to specific regions of the gene. However, DCM causing variants have been recorded across the gene and are so far all reported with dominant inheritance.

| Gene | *MYH7* |
| --- | --- |
| OMIM gene number | 160760 |
| Referral indication | Dilated Cardiomyopathy (DCM) |
| Disease grouping | Familial Dilated Cardiomyopathy |
| **Disease name** | ***MYH7*-related DCM** |
| MONDO ID | 0013262 |
| Gene disease validity (ClinGen) | DEFINITIVE |
| Inheritance | Autosomal dominant |
| Allelic requirement | Monoallelic autosomal |
| Inheritance modifiers | Typified by incomplete penetrance |
| Disease-associated variant consequence | Altered gene product sequence |
| Variant classes reported with evidence of pathogenicity | Missense |
| Restricted repertoire of pathogenic variants | NA |
| PMIDs | 29300372; 31983221; 11106718; 29666183; 29093449 |

*MYH7* encodes the β-myosin heavy chain, part of the sarcomere which plays a major role in cardiac muscle contraction.

Pathogenic variants in *MYH7* cause DCM due to **altered gene product sequence** primarily due to decrease in sarcomere force generation.

Dominant-negative missense variants resulting in an altered protein with reduced function are responsible for most MYH7-associated DCM cases, and a small number of in-frame indels have also been reported associated with disease. The likely disease mechanism is a reduction in the passive stiffness of myofibrils (PMID: 29093449) and deficit in force generation and force-holding capacity (PMID: 29666183). There is no good evidence that loss of function is a disease mechanism (e.g. PMID: 31983221), and MYH7 is not known to be haploinsufficient.

*MYH7*-related DCM is inherited in an autosomal dominant manner, and disease has incomplete penetrance, age-related onset, and variable expressivity. Pathogenic *MYH7* variants are likely to account for approximately 5-6% of familial DCM cases. There is evidence that biallelic variants can have an additive effect resulting in more severe cardiomyopathic phenotypes.

| Gene | *RBM20* |
| --- | --- |
| OMIM gene number | 613171 |
| Referral indication | Dilated Cardiomyopathy (DCM) |
| Disease grouping | Familial Dilated Cardiomyopathy |
| **Disease name** | ***RBM20*-related DCM** |
| MONDO ID | 0013168 |
| Gene disease validity (ClinGen) | DEFINITIVE |
| Inheritance | Autosomal dominant |
| Allelic requirement | Monoallelic autosomal |
| Inheritance modifiers | Typified by incomplete penetrance |
| Disease-associated variant consequence | Decreased gene product level; altered gene product sequence |
| Variant classes reported with evidence of pathogenicity | Missense; stop gained NMD triggering |
| Restricted repertoire of pathogenic variants | There is a mutation hotspot in exon 9 (amino acids 634-638) |
| PMIDs | 19712804; 22466703; 29650543; 32789749; 25979592; 26084686; 21846512; 20590677; 22004663; 29895960; 30871348; 30871351; 32851336; 29367541; 27496873; 29650543 |

Pathogenic variants in *RBM20* cause DCM due to **decreased gene product level or altered gene product sequence** due to a variety of mechanisms including altered splicing of targets. A dominant negative effect causing disrupted RNA binding is also a possible/likely mechanism.

Missense variants are responsible for the majority of DCM cases although a small number of truncating variants (nonsense) have been reported. Of note, there are multiple ClinVar entries of truncating variants associated with DCM reported by diagnostic laboratories, comparing to the very low number of truncating variants detected in gnomAD.

**Loss of function via truncating variants is not absolutely established, but highly likely.**

RBM20-related DCM is inherited in an autosomal dominant manner, and disease is often an aggressive arrhythmogenic phenotype with high rates of heart failure and sudden cardiac death. Pathogenic *RBM20* variants are likely to account for approximately 1.5-3% of familial DCM cases. There is a pathogenic variant hotspot in exon 9 (RS motif, amino acids 634-638).

| Gene | *SCN5A* |
| --- | --- |
| OMIM gene number | 601154 |
| Referral indication | Dilated Cardiomyopathy (DCM) |
| Disease grouping | Familial Dilated Cardiomyopathy |
| **Disease name** | ***SCN5A*-related DCM** |
| MONDO ID | 0011003 |
| Gene disease validity (ClinGen) | DEFINITIVE |
| Inheritance | Autosomal dominant |
| Allelic requirement | Monoallelic autosomal |
| Inheritance modifiers | NA |
| Disease-associated variant consequence | Decreased gene product level; altered gene product sequence |
| Variant classes reported with evidence of pathogenicity | Missense; stop gained NMD triggering |
| Restricted repertoire of pathogenic variants | NA |
| PMIDs | 26916278; 15671429; 15466643; 19808398; 21596231; 22675453; 22999724; 30847666; 27532257 |

Pathogenic variants in *SCN5A* cause DCM mainly due to **altered gene product sequence leading to gain of function, and more rarely decreased/absent gene product level.** The mechanism by which changes in sodium conductivity lead to cardiomyopathy is not fully understood; however, it is thought to be due to disruption of the voltage-sensing mechanism of this channel and subsequent disruption to action potential and cardiac contraction over time. It is unclear whether loss of function is a true disease mechanism (for DCM).

Pathogenic variants in *SCN5A* are likely to cause <2% of familial DCM cases (PMID: 26916278, 21596231) and are associated with an arrhythmogenic phenotype with high rates of sudden cardiac death. There are no defined hotspot regions for *SCN5A* missense variants in DCM, however, they do appear to commonly lie within voltage sensing regions (S3 and S4 transmembrane segments) of the protein.

Of note *SCN5A* loss of function is generally more associated with Brugada syndrome (BrS); *SCN5A* is the only gene definitively associated with BrS. Gain of function variants are also associated with Long QT syndrome.

| Gene | *TNNC1* |
| --- | --- |
| OMIM gene number | 191040 |
| Referral indication | Dilated Cardiomyopathy (DCM) |
| Disease grouping | Familial Dilated Cardiomyopathy |
| **Disease name** | ***TNNC1*-related DCM** |
| MONDO ID | 0012745 |
| Gene disease validity (ClinGen) | DEFINITIVE |
| Inheritance | Autosomal dominant |
| Allelic requirement | Monoallelic autosomal |
| Inheritance modifiers | NA |
| Disease-associated variant consequence | Altered gene product sequence |
| Variant classes reported with evidence of pathogenicity | Missense |
| Restricted repertoire of pathogenic variants | NA |
| PMIDs | 1554228; 19808376; 17021793; 18803402; 32038292; 27604170; 18212018; 20458010; 27532257; 17977476 |

Pathogenic variants in *TNNC1* are a rare (<1%) cause of familial DCM due to **altered gene product sequence***.* This leads to a likely ‘poison peptide’ dominant negative effect, causing alteration of troponin interactions, and altered (decreased) calcium binding of myofilaments and resulting in decreased force production.

Missense variants are responsible for all reported cases of *TNNC1*-related DCM, with no evidence that truncating variants are causative of disease.

There is no apparent enrichment for non-truncating mutations in any specific domain, although exon 1-3 show regional constraint (gnomAD database).

*TNNC1*-related DCM is inherited as an autosomal dominant trait, with high penetrance observed in families. There is currently insufficient evidence for autosomal recessive inheritance, although there are some reports of early onset DCM and other cardiomyopathy associated with compound heterozygosity for *TNNC1* variants (PMID: 27604170).

| Gene | *TNNT2* |
| --- | --- |
| OMIM gene number | 601494 |
| Referral indication | Dilated Cardiomyopathy (DCM) |
| Disease grouping | Familial Dilated Cardiomyopathy |
| **Disease name** | ***TNNT2*-related DCM** |
| MONDO ID | 0011095 |
| Gene disease validity (ClinGen) | DEFINITIVE |
| Inheritance | Autosomal dominant |
| Allelic requirement | Monoallelic autosomal |
| Inheritance modifiers | NA |
| Disease-associated variant consequence | Altered gene product sequence |
| Variant classes reported with evidence of pathogenicity | Missense |
| Restricted repertoire of pathogenic variants | NA |
| PMIDs | 27532257; 11106718; 15542288; 20031601; 20978592; 29367541 |

Pathogenic variants in *TNNT2* cause DCM due to **altered gene product sequence.**

Only missense variants have confidently been reported as pathogenic in *TNNT2* DCM cases. There are no reports of truncating variants and *TNNT2* has a ClinGen Dosage haploinsufficiency score of 0, and a PLi of 0 in gnomAD (19.6 expected and 20 observed).

*TNNT2-*related DCM is inherited in an autosomal dominant manner, and disease is often an aggressive arrhythmogenic phenotype with high rates of heart failure and sudden cardiac death. *TNNT2* missense mutations account for 3% of genetic DCM (PMID:27532257).

*TNNT2* variants are associated with an early-onset and more severe form of DCM.

Of note pathogenic variants in *TNNT2* are also associated hypertrophic and restrictive cardiomyopathies.

| Gene | *TTN* |
| --- | --- |
| OMIM gene number | 188840 |
| Referral indication | Dilated Cardiomyopathy (DCM) |
| Disease grouping | Familial Dilated Cardiomyopathy |
| **Disease name** | ***TTN*-related DCM** |
| MONDO ID | 0011400 |
| Gene disease validity (ClinGen) | DEFINITIVE |
| Inheritance | Autosomal dominant |
| Allelic requirement | Monoallelic autosomal |
| Inheritance modifiers | Typified by incomplete penetrance; Typified by age-related onset |
| Disease-associated variant consequence | Decreased gene product level; altered gene product sequence |
| Variant classes reported with evidence of pathogenicity | Splice acceptor; splice donor; frameshift variant NMD triggering; stop gained NMD triggering; missense [v rare]; structural variants (whole exon deletions); [classes must occur in A-band or >90% PSI exons] |
| Restricted repertoire of pathogenic variants | Variants must occur in exons (PSI > 0.9)  Missense variants with segregation evidence:  p.Trp976Arg  p.Ala178Asp  p.Cys3575Ser |
| PMIDs | 9817758; 9826585; 19789381; 18765796; 25589632; 11788824; 22335739; 32013205; 23418287; 26084686; 28045975; 25759365; 26315439; 29316444; 27869827; 29238064; 32160020; 27869827; 31849696; 27625337; 11788824 |

Pathogenic variants in *TTN* cause an estimated 15-20% of familial DCM due to **decreased gene product level and altered gene product sequence**. The likely disease mechanism is loss of function however, it is unclear whether this is due to haploinsufficiency or a dominant negative effect, and it is likely that both mechanisms contribute. It is likely that the reduced function has a direct effect on the sarcomere, leading to impaired contractility.

Truncating variants, specifically in exons constitutively expressed in cardiac tissue (PSI >0.9) see (PMID: **25589632; 27869827;** 32160020), are responsible for the vast majority of *TTN*-related DCM cases. Missense variants are difficult to interpret, and generally not classified as disease-causing, although there are reports of at least three missense variants with evidence of pathogenicity.

*TTN* missense variants with segregation evidence:

p.Trp976Arg (PMID: 11788824)

p.Ala178Asp (PMID: 27625337)

p.Cys3575Ser (<https://www.biorxiv.org/content/10.1101/2020.09.05.282913v1.full.pdf>) – note not yet peer reviewed

*TTN*-related DCM is inherited as an autosomal dominant trait and displays incomplete and age-related onset. *TTN* truncating variants are present in ~1% of the general population, although these variants are more likely to reside in isoforms with lower functional expression in cardiac tissue.

| Gene | *PLN* |
| --- | --- |
| OMIM gene number | 172405 |
| Referral indication | Dilated Cardiomyopathy (DCM) |
| Disease grouping | Familial Dilated Cardiomyopathy |
| **Disease name** | ***PLN*-related intrinsic cardiomyopathy** |
| MONDO ID | 0012362 |
| Gene disease validity (ClinGen) | DEFINITIVE |
| Inheritance | Autosomal dominant |
| Allelic requirement | Monoallelic autosomal |
| Inheritance modifiers | Typified by age-related onset |
| Disease-associated variant consequence | Decreased gene product level; altered gene product sequence |
| Variant classes reported with evidence of pathogenicity | Missense; inframe deletion; frameshift; stop gained; structural variants (whole exon deletions) |
| Restricted repertoire of pathogenic variants | NA |
| PMIDs | 12639993; 16432188; 33020536; 21167350 |

Pathogenic variants in *PLN* cause cardiomyopathy by **decreased gene product level or altered gene product sequence**.

Inheritance is typically autosomal dominant with incomplete penetrance but biallelic variants have been described and appear to confer an earlier onset and more severe phenotype (PMID: 12639993)

***PLN* is encoded by one coding exon** (52 amino acids). Schmitt et al described a missense variant (p.Arg9Cys) in a patient with DCM. The variant segregated with disease in the family; transgenic mice developed biventricular dilatation (PMID 12610310). A stop gained variant and inframe deletion have also been described (PMID: 12639993; PMID 16432188). There are 6 pathogenic/likely pathogenic variants reported on ClinVar: 1 missense, 2 stop gained, 2 frameshift and a large deletion all associated with dilated cardiomyopathy.

In the Netherlands there is a founder mutation p.Arg14del. Up to 10–15% of both dilated cardiomyopathy and arrhythmogenic cardiomyopathy patients are reported to be caused by *PLN*-R14del. (PMID: 33020536)

ClinGen found no difference in the molecular mechanism(s) underlying *PLN*-related DCM and HCM and observed that inter and intrafamilial variability in phenotype had been reported. Haghighi et al describe the same variant (p.Leu39X) in a family causing severe DCM in the homozygous state and both DCM and HCM phenotypes in the heterozygous state (PMID: 12639993). This variant has also been reported in other HCM families (PMID: 21167350). As a result, ClinGen curated *PLN* for an association with intrinsic cardiomyopathy and did not separately evaluate the evidence for the role in hypertrophic vs dilated phenotypes.

*PLN* is definitively associated with cardiomyopathy and the majority of variants reported appear to be associated with DCM. Experts commented that further investigation into *PLN* and HCM and the variant classes associated needs to be undertaken.

##### HYPERTROPHIC CARDIOMYOPATHY – Familial hypertrophic cardiomyopathy

| Gene | *ACTC1* |
| --- | --- |
| OMIM gene number | 102540 |
| Referral indication | Hypertrophic Cardiomyopathy (HCM) |
| Disease grouping | Familial hypertrophic cardiomyopathy |
| **Disease name** | ***ACTC1*-related HCM** |
| MONDO ID | 0012799 |
| Gene disease validity (ClinGen) | DEFINITIVE |
| Inheritance | Autosomal dominant |
| Allelic requirement | Monoallelic autosomal |
| Inheritance modifiers | NA |
| Disease-associated variant consequence | Altered gene product sequence |
| Variant classes reported with evidence of pathogenicity | Missense; inframe deletion |
| Restricted repertoire of pathogenic variants | NA |
| PMIDs | 10330430; NBK1768; 20031618; 28007147; 27532257; 10966831; 17611253; 26061005; 23604709; 28972856 |

*ACTC1* pathogenic variants cause HCM through **altered gene product sequence.** The disease mechanism is not definitively known but may involve impact on sarcomere force generation. There is currently little evidence to support haploinsufficiency as a mechanism.

Inheritance is autosomal dominant. There is limited information regarding penetrance. The initial missense variant identified by Mogensen et al was reported to be highly penetrant in family members but with a variable age of onset and severity (PMID:10330430).

*ACTC1* variants account for <3-5% of HCM cases (NBK1768; PMID: 20031618)

**Heterozygous missense variants are the major type of pathogenic variants found**. One inframe deletion (a deletion of a single amino acid, Phe92del) has been reported in association with HCM (PMID: 20031618). This variant is classified as likely pathogenic on ClinVar. Liu et al suggest it may act to change the local structure and arrangement of amino acids in the actomyosin binding site (PMID: 28972856)

Large deletions or duplications have not been described.

| Gene | *MYBPC3* |
| --- | --- |
| OMIM gene number | 600958 |
| Referral indication | Hypertrophic Cardiomyopathy (HCM) |
| Disease grouping | Familial hypertrophic cardiomyopathy |
| **Disease name** | ***MYBPC3*-related HCM** |
| MONDO ID | 0007268 |
| Gene disease validity (ClinGen) | DEFINITIVE |
| Inheritance | Autosomal dominant |
| Allelic requirement | Monoallelic autosomal |
| Inheritance modifiers | Typified by incomplete penetrance |
| Disease-associated variant consequence | Decreased gene product level; altered gene product sequence |
| Variant classes reported with evidence of pathogenicity | Splice region; splice acceptor; splice donor; frameshift; frameshift variant NMD triggering; stop gained; stop gained NMD triggering; missense; inframe insertion; inframe deletion; intronic; structural variants (whole exon deletions) |
| Restricted repertoire of pathogenic variants | NA |
| PMIDs | 28007147; 28912181; 7493025; 26573135; 25611685; 32731933; 29300372; NBK1768; 25611685; 30696458; 12707239; 20624503; 28912181; 27532257; 31877118; 22057632; 32163302; 32396390; 18467358; 17937428; 19151713; 32841044 |

*MYBPC3* pathogenic variants cause HCM through **decreased gene product level or altered gene product sequence** either leading to a reduction in MyBP-C content in the sarcomere **or** altered function. The disease mechanism is **loss of function**; There is evidence of haploinsufficiency (PMID 31877118; 22057632; 32841044).

Variants in *MYBPC3* and *MYH7* collectively account for up to 50% of all clinically recognised cases of HCM and constitute at least 75% of probands where a variant is identified (PMID: 28007147; NBK1768).

Inheritance is usually autosomal dominant, typified by incomplete penetrance and variable expressivity.

Homozygous and compound heterozygous variants have been reported and can lead to severe, early onset phenotypes (PMID: 26573135; 25611685; PMID: 18467358; PMID: 17937428).

**The majority of variants are heterozygous frameshift, nonsense, or splice site variants** that result in premature termination codons (PMID: 25611685; 31877118). Missense and inframe indels are also frequently reported and a subset have been shown to cause loss of function through failure of myofilament incorporation and rapid degradation, further supporting haploinsufficiency as a mechanism (PMID 32841044). Variants in *MYBPC3* affecting canonical splice site dinucleotides are a well-characterised cause of HCM. Furthermore, recent work has identified more deeply intronic variants associated with disease (<https://doi.org/10.3390/cardiogenetics11020009>**;** PMID: 32396390). There are 39 pathogenic/likely pathogenic *MYBPC3* intronic variants submitted on ClinVar.

The common intronic deletion, *MYBPC3*^Δ25^, detected in 4% to 8% of South Asian populations, is associated with cardiomyopathy as a risk allele (PMID: 19151713)

Although there is significant genetic and allelic heterogeneity in HCM, there are also several *MYBPC3* founder variants (PMID: 28912181; 27532257).

| Gene | *MYH7* |
| --- | --- |
| OMIM gene number | 160760 |
| Referral indication | Hypertrophic Cardiomyopathy (HCM) |
| Disease grouping | Familial hypertrophic cardiomyopathy |
| **Disease name** | ***MYH7*-related HCM** |
| MONDO ID | 0008647 |
| Gene disease validity (ClinGen) | DEFINITIVE |
| Inheritance | Autosomal dominant |
| Allelic requirement | Monoallelic autosomal |
| Inheritance modifiers | Typified by incomplete penetrance |
| Disease-associated variant consequence | Altered gene product sequence |
| Variant classes reported with evidence of pathogenicity | Missense; inframe deletion; stop gained NMD escaping |
| Restricted repertoire of pathogenic variants | NA |
| PMIDs | 29300372; 27532257; 30696458; 30924982; 25209314; 1975517; 1944483; 1552912; 15856146; 34460321; 30681346; 25611685; 20359594; 32731933; 12788380; 33500567; 27247418; 27532257 |

*MYH7* pathogenic variants cause HCM through **altered gene product sequence** leading to an increase in sarcomere force generation. Variants produce an abnormal activated protein that incorporates into the sarcomere as a ‘poison peptide’. Variants can either directly affect motor function or can impact on myosin "interacting head motif" and therefore impair inactivation kinetics. There is currently no evidence to support haploinsufficiency as a disease mechanism.

Inheritance is autosomal dominant, typified by incomplete penetrance and variable expressivity.

**Most pathogenic variants are missense**. There are some inframe deletions reported.

A frameshift variant has been identified in 3.3% of Egyptian HCM patients. It is predicted to result in a premature termination codon downstream of the last exon-exon junction of the gene that is expected to escape nonsense-mediated decay (NMD). (PMID: 34460321)

*MYH7* loss of function (LoF) variants are very rare and their contribution to inherited cardiomyopathy is incompletely understood. Notably in a recent study of LVNC, *MYH7* truncating variants, generally considered non-pathogenic for cardiomyopathies, were 20-fold enriched in LVNC cases over controls (PMID: 33500567)

While there is currently no evidence for a disease-causing role in the heterozygous state **in HCM**, compound heterozygosity of LoF variants along with missense variants can lead to extremely severe presentations, mimicking recessive inheritance.

There is a clustering of HCM variants in the head region conferring a high probability of pathogenicity (amino acid residues 181-937). (PMID: 27247418; 27532257; 29300372; 30696458)

Kelly et al 2018 (PMID 29300372) provide gene-specific adaptations of ACMG criteria for *MYH7* and HCM.

| Gene | *MYL2* |
| --- | --- |
| OMIM gene number | 160781 |
| Referral indication | Hypertrophic Cardiomyopathy (HCM) |
| Disease grouping | Familial hypertrophic cardiomyopathy |
| **Disease name** | ***MYL2*-related HCM** |
| MONDO ID | 0012112 |
| Gene disease validity (ClinGen) | DEFINITIVE |
| Inheritance | Autosomal dominant |
| Allelic requirement | Monoallelic autosomal |
| Inheritance modifiers | NA |
| Disease-associated variant consequence | Altered gene product sequence |
| Variant classes reported with evidence of pathogenicity | Missense |
| Restricted repertoire of pathogenic variants | NA |
| PMIDs | NBK1768; 32731933; 8673105; 9535554; 12404107; 30696458; 32453731; 23365102; 28007147; 28912181; 25611685; 16837010; 25324513; 16076902; 24111713 |

*MYL2* pathogenic variants cause autosomal HCM through **altered gene product sequence.** The disease mechanism is not definitively known but may involve destabilization of the interacting heads motif (PMID: 28606303).

There is currently insufficient evidence to support haploinsufficiency (https://search.clinicalgenome.org/kb/gene-dosage/HGNC:7583)

*MYL2* variants account for <3% of HCM cases (NBK1768; PMID: 32731933).

Inheritance is typically autosomal dominant. There is limited information regarding penetrance.

Homozygous and compound heterozygous variants have been described in association with a lethal myosinopathy (PMID 23365102).

A study reported a homozygous frameshift variant causing infantile onset HCM. Heterozygous parents were unaffected. The authors suggest a molecular mechanism by which loss-of-function variants in *MYL2* are recessive while missense variants are dominant in HCM. Several loss-of-function variants are reported in gnomAD suggesting *MYL2* is not intolerant to LoF variants. (PMID: 32453731)

**The majority of variants reported are missense** (PMID 8673105; PMID 9535554; PMID 12404107.)

On ClinVar, nearly all pathogenic variants are missense. There is one frameshift variant associated only with cardiomyopathy, not HCM specifically and a deletion encompassing exon 7 expected to result in a truncated protein - <https://www.ncbi.nlm.nih.gov/clinvar/variation/417460/>

Walsh et al found the *MYL2* gene to be significantly enriched for non-truncating variants, odds ratio 9.1 (6.2-13.3) (PMID: 30696458).

| Gene | *MYL3* |
| --- | --- |
| OMIM gene number | 160790 |
| Referral indication | Hypertrophic Cardiomyopathy (HCM) |
| Disease grouping | Familial hypertrophic cardiomyopathy |
| **Disease name** | ***MYL3*-related HCM** |
| MONDO ID | 0012111 |
| Gene disease validity (ClinGen) | DEFINITIVE |
| Inheritance | Autosomal dominant |
| Allelic requirement | Monoallelic autosomal |
| Inheritance modifiers | NA |
| Disease-associated variant consequence | Altered gene product sequence |
| Variant classes reported with evidence of pathogenicity | Missense |
| Restricted repertoire of pathogenic variants | NA |
| PMIDs | 33288880; NBK1768; PMID: 20031618; 28369730; 26443374; 12021217; 30696458; 8673105; 20031618; 25611685; 22957257; 29914921 |

*MYL3* pathogenic variants cause HCM through **altered gene product sequence.** The disease mechanism is not definitively known but may involve impairing protein-protein interaction with components of the sarcomere and destabilizing the interacting heads motif (PMID: 30275503; PMID: 28606303). There is currently no evidence to support haploinsufficiency as a mechanism (https://search.clinicalgenome.org/kb/gene-dosage/HGNC:7584).

*MYL3* variants account for <3% of HCM cases (NBK1768; PMID: 20031618; PMID: 28369730).

Inheritance is primarily autosomal dominant. There is limited information regarding penetrance given small numbers of variants identified. In a large family where a missense variant was identified (p. Arg94His), penetrance was estimated at 88% (PMID: 26443374).

Osborn et al 2021 (PMID: 33288880) report a homozygous missense variant in a large family with HCM and sudden cardiac death. Heterozygous carriers were unaffected. Another biallelic variant was also described by Olson et al in 2002 (E143K) (PMID: 12021217), however this variant has also been associated with HCM variably in the heterozygous state.

**Heterozygous missense variants are the major type of pathogenic variants found** (PMID 8673105; 20031618; 25611685). There are reports on ClinVar of frameshift and splice site variants, but these are classified as uncertain significance or conflicting. There is currently insufficient evidence to support loss of function as a mechanism.

Walsh et al reported a cluster of non-truncating *MYL3* variants in HCM at amino acid residues 143-180 (PMID: 30696458).

| Gene | *TNNI3* |
| --- | --- |
| OMIM gene number | 191044 |
| Referral indication | Hypertrophic Cardiomyopathy (HCM) |
| Disease grouping | Familial hypertrophic cardiomyopathy |
| **Disease name** | ***TNNI3*-related HCM** |
| MONDO ID | 0013369 |
| Gene disease validity (ClinGen) | DEFINITIVE |
| Inheritance | Autosomal dominant |
| Allelic requirement | Monoallelic autosomal |
| Inheritance modifiers | Typified by incomplete penetrance |
| Disease-associated variant consequence | Altered gene product sequence |
| Variant classes reported with evidence of pathogenicity | Missense; inframe deletion |
| Restricted repertoire of pathogenic variants | NA |
| PMIDs | 9241277; 28912181; 21839045; 23270746; 15607392; NBK1768; 32731933; 26440512; 25611685; 30696458; 21415410 |

*TNNI3* pathogenic variants cause HCM through **altered gene product sequence.** The disease mechanism is not definitively known but a review by Tardif et al reported functional studies on missense variants that found an increase in the Ca^2+^ sensitivity of myofilament activation (PMID: 21415410). ClinGen also conclude that “missense mutations have [been] shown to affect Ca2+ binding to myofilaments containing the mutant TNNI3 (PMIDs: 16531415 and 22675533) or result in an increased myofilament response to Ca2+ (PMID: 11735257).” https://search.clinicalgenome.org/kb/genes/HGNC:11947

*TNNI3* variants account for approx. 3-5% of HCM cases (NBK1768; PMID: 15607392)

Inheritance is **autosomal dominant** characterised by **incomplete penetrance** (PMID 26440512; 15607392; 9241277). Lorenzini et al found that subjects with *TNNI3* variants had a lower penetrance than variants in *MYBPC3, MYH7,* and *TNNT2* (PMID: 32731933). Experts commented that variants seen in *TNNI3* in severe/early onset disease were more likely to be *de novo* in origin.

Maron et al 2012 describe 4 HCM probands with both pathogenic variants in *TNNI3* and *MYBPC3* (PMID 21839045). In addition, 2 siblings have been reported with homozygous *TNNI3* missense variants (Arg162Trp) and severe myocardial hypertrophy. Parents of the affected children are consanguineous and along with other family members harbouring the same variant in the heterozygous state, were unaffected (PMID 23270746). Multiple disease-causing sarcomeric variants appear to be associated with more severe disease. There is not enough evidence to suggest biallelic variants in *TNNI3* have a distinct mechanism compared to monoallelic variants. Of note both dominant and recessive modes of inheritance have been reported in *TNNI3*-related DCM (PMID: 26440512).

**Heterozygous missense variants are the major type** of pathogenic variants found. There are also reports of **inframe deletions**. In a systematic review, Mogensen et al reported 91% of all variants were missense variants. Six variants (Arg141Gln, Arg145Trp, Arg157Val, Arg162Gln, Ser166Phe, and Lys183Del) appeared with a particularly high frequency and were identified in 116 of the 256 probands (45%) (PMID 26440512). There are reports on ClinVar of frameshift, splice site variants and nonsense variants but these are either classified as uncertain significance/conflicting, not associated with HCM specifically or have no functional evidence to demonstrate loss of function as a mechanism. ClinGen have concluded that there is currently limited evidence to support haploinsufficiency as a mechanism.

Mogensen et al found that 85% of variants were identified in exons 7 and 8 (amino acid residues 125 – 210). (PMID 26440512)

Similarly, Walsh et al detected a cluster of non-truncating *TNNI3* variants in HCM at amino acid residues 141–209 (PMID: 30696458).

| Gene | *TNNT2* |
| --- | --- |
| OMIM gene number | 191045 |
| Referral indication | Hypertrophic Cardiomyopathy (HCM) |
| Disease grouping | Familial hypertrophic cardiomyopathy |
| **Disease name** | ***TNNT2*-related HCM** |
| MONDO ID | 0007266 |
| Gene disease validity (ClinGen) | DEFINITIVE |
| Inheritance | Autosomal dominant |
| Allelic requirement | Monoallelic autosomal |
| Inheritance modifiers | Typified by incomplete penetrance |
| Disease-associated variant consequence | Altered gene product sequence |
| Variant classes reported with evidence of pathogenicity | Missense; stop gained NMD escaping; inframe deletion; splice donor variant NMD escaping |
| Restricted repertoire of pathogenic variants | NA |
| PMIDs | 8205619; 28973951; 7898523; 28007147; 28912181; 30578328; 10965086; 11034944; 32731933; NBK1768; 12707239; 22144547; 25611685; 30696458 |

*TNNT2* pathogenic variants cause HCM through **altered gene product sequence.** Troponin T is a regulatory protein found in striated muscles that forms a complex with troponin I (TnI) and troponin C (TnC) that, together with tropomyosin (TM), is required for Ca^2+^ dependent regulation of muscle contraction.

The mechanism appears to be dominant negative rather than haploinsufficiency. The majority of pathogenic variants are missense. A functional study of a splice variant associated with HCM concluded that the resulting truncated protein does not function as a “null protein, but rather as a dominant-negative leading to reduction in the level of calcium activated force production” (PMID: 8958207). Gangadharan et al suggest the primary reason by which *TNNT2* variants between residues 92 and 144 cause cardiomyopathy is by changing the affinity of TnT for Tm within the N terminal part of Troponin T (PMID: 28973951).

*TNNT2* variants account for <5% of HCM cases (NBK1768; 12860912)

Inheritance is **autosomal dominant** characterised by **incomplete penetrance** (PMID: 32731933).

Homozygous *TNNT2* variants causing HCM are rare and have been reported to be associated with more severe disease (PMID 30578328; 10965086; 11034944). Piroddi et al investigated a patient with severe, early onset HCM with a homozygous K280N variant. There was no family information available, however the authors demonstrated that it resulted in 100% mutant cTnT with no evidence of haploinsufficiency and suggested this supports the idea of a gene dose–dependent effect of HCM variants on the severity of the phenotype (PMID 30578328).

Experts commented that variants seen in *TNNT2* in severe/early onset disease were more likely to be *de novo* in origin.

**Nearly all pathogenic variants are missense** (PMID: 12707239; 22144547; 25611685). However, there are a few reports of inframe deletions, nonsense variants in the final exon which likely escape nonsense mediated decay and splice donor variants (PMID: 8958207; 12707239; 22144547; 25611685).

<https://www.ncbi.nlm.nih.gov/clinvar/variation/43673/>; <https://www.ncbi.nlm.nih.gov/clinvar/variation/177636/>

Walsh et al found non truncating variants in the *TNNT2* gene to be significantly enriched in HCM cases, odds ratio 11.4 (8.5 – 15.2). They found a clustering of non-truncating variants conveying a high probability of pathogenicity in the tropomyosin binding domain (amino acid residues 79 – 179) (PMID: 30696458).

| Gene | *TPM1* |
| --- | --- |
| OMIM gene number | 191010 |
| Referral indication | Hypertrophic Cardiomyopathy (HCM) |
| Disease grouping | Familial hypertrophic cardiomyopathy |
| **Disease name** | ***TPM1*-related HCM** |
| MONDO ID | 0007267 |
| Gene disease validity (ClinGen) | DEFINITIVE |
| Inheritance | Autosomal dominant |
| Allelic requirement | Monoallelic autosomal |
| Inheritance modifiers | Typified by incomplete penetrance |
| Disease-associated variant consequence | Altered gene product sequence |
| Variant classes reported with evidence of pathogenicity | Missense |
| Restricted repertoire of pathogenic variants | NA |
| PMIDs | 8205619; 24005378; 12860912; NBK1768; 7898523 |

*TPM1* pathogenic variants cause HCM through **altered gene product sequence.** *TPM1* encodes alpha-tropomyosin which acts to place the troponin complex on cardiac actin. The mechanism is through altered function rather than haploinsufficiency – Bottinelli et al examined the Asp175Asn variant from 2 HCM patients and found equal expression of wild type and mutant alpha-tropomyosin proteins (PMID 9440709). Gupte et al concluded that “*TPM1* mutations cause differences in protein stability, actin binding, and Tn conformation. All of these differences could converge to change the Ca^2+^ dependence of myosin activity” (PMID: 25548289).

*TPM1* variants account for <3% of HCM cases (NBK1768; PMID: 7898523; 12860912).

Inheritance is **autosomal dominant** characterised by **incomplete penetrance** (PMID: 32731933).

There are reports of homozygous or compound heterozygous variants in *TPM1.* (PMID: 33642254; PMID: 32744700).

A homozygous missense variant, (p.Gly3Arg), in exon 1 of *TPM1* was identified in triplets (two had HCM and one patent ductus arteriosus). The parents were heterozygous for the variant and unaffected clinically and on echocardiogram (PMID: 32744700).

**Pathogenic variants are nearly always heterozygous missense variants.** There is one *de novo* inframe deletion (6bp, 2 amino acids) associated with HCM classified as likely pathogenic on ClinVar, no additional details are given.

Redwood and Robinson reviewed *TPM1* variants in 2013 and reported “at least 15 described in the current literature (Table1). Most of these mutations are unique and have been reported in only a single family or individual…Each mutation is missense causing a single amino acid substitution, and no truncation or nonsense mutations have been reported.” (PMID 24005378)

Walsh et al found that in HCM non-truncating variants across the whole *TPM1* gene showed a high probability of pathogenicity. (PMID: 30696458)

##### HYPERTROPHIC CARDIOMYOPATHY – Intrinsic cardiomyopathy

| Gene | *PLN* |
| --- | --- |
| OMIM gene number | 172405 |
| Referral indication | Hypertrophic Cardiomyopathy (HCM) |
| Disease grouping | Familial Hypertrophic Cardiomyopathy |
| **Disease name** | ***PLN*-related intrinsic cardiomyopathy** |
| MONDO ID | 0012362 |
| Gene disease validity (ClinGen) | DEFINITIVE |
| Inheritance | Autosomal dominant |
| Allelic requirement | Monoallelic autosomal |
| Inheritance modifiers | Typified by incomplete penetrance |
| Disease-associated variant consequence | Decreased gene product level; altered gene product sequence |
| Variant classes reported with evidence of pathogenicity | Missense; inframe deletion; frameshift; stop gained; structural variants (whole exon deletions) |
| Restricted repertoire of pathogenic variants | NA |
| PMIDs | 12639993; 16432188; 33020536; 21167350 |

Pathogenic variants in *PLN* cause cardiomyopathy by **decreased gene product level or altered gene product sequence**.

Inheritance is typically autosomal dominant with incomplete penetrance but biallelic variants have been described and appear to confer an earlier onset and more severe phenotype (PMID: 12639993)

***PLN* is encoded by one coding exon** (52 amino acids). Schmitt et al described a missense variant (p.Arg9Cys) in a patient with DCM. The variant segregated with disease in the family; transgenic mice developed biventricular dilatation (PMID 12610310). A stop gained variant and inframe deletion have also been described (PMID: 12639993; PMID 16432188). There are 6 pathogenic/likely pathogenic variants reported on ClinVar: 1 missense, 2 stop gained, 2 frameshift and a large deletion all associated with dilated cardiomyopathy.

In the Netherlands there is a founder mutation p.Arg14del. Up to 10–15% of both dilated cardiomyopathy and arrhythmogenic cardiomyopathy patients are reported to be caused by *PLN*-R14del. (PMID: 33020536)

ClinGen found no difference in the molecular mechanism(s) underlying *PLN*-related DCM and HCM and observed that inter and intrafamilial variability in phenotype had been reported. Haghighi et al describe the same variant (p. Leu39X) in a family causing severe DCM in the homozygous state and both DCM and HCM phenotypes in the heterozygous state (PMID: 12639993). This variant has also been reported in other HCM families (PMID: 21167350). As a result, ClinGen curated *PLN* for an association with intrinsic cardiomyopathy and did not separately evaluate the evidence for the role in HCM and DCM phenotypes.

*PLN* is definitively associated with cardiomyopathy and the majority of variants reported appear to be associated with DCM. Experts commented that further investigation into *PLN* and HCM and the variant classes associated needs to be undertaken.

##### HYPERTROPHIC CARDIOMYOPATHY – rare syndromic disorders with HCM that can present with isolated left ventricular hypertrophy (LVH)

| Gene | *ALPK3* |
| --- | --- |
| OMIM gene number | 617608 |
| Referral indication | Hypertrophic Cardiomyopathy (HCM) |
| Disease grouping | Rare syndromic disorder with hypertrophic cardiomyopathy - isolated LVH |
| **Disease name** | ***ALPK3*-related hypertrophic cardiomyopathy** |
| MONDO ID | 0011001 |
| Gene disease validity (ClinGen) | STRONG |
| Inheritance | Autosomal recessive; Autosomal dominant |
| Allelic requirement | Biallelic autosomal; monoallelic autosomal |
| Inheritance modifiers | NA |
| Disease-associated variant consequence | Decreased gene product level; altered gene product sequence |
| Variant classes reported with evidence of pathogenicity | Stop gained; frameshift; splice acceptor; splice donor; missense |
| Restricted repertoire of pathogenic variants | NA |
| PMIDs | 27106955; 31074094; 32480058; 26846950; 21441111; 34263911; 34263907; 33191771 |

*ALPK3* pathogenic variants cause hypertrophic cardiomyopathy through **decreased gene product level** or **altered gene product sequence**.

**Biallelic** *ALPK3* variants were first associated with a rare recessive form of cardiomyopathy by Almomani et al in 2016 (PMID:26846950), with several other reports since (PMID:27106955; PMID: 31074094; PMID: 32480058). Patients display variable phenotypes but often present at birth or early childhood (at least 4 patients have presented in utero) with dilated cardiomyopathy (DCM) that progressed to a HCM phenotype over time. **Extra-cardiac features including musculoskeletal and craniofacial abnormalities** are also commonly observed in these cases (PMID: 32480058; PMID: 34263911).

Herkert et al 2020 reviewed the variants and phenotype in 19 paediatric patients with biallelic *ALPK3* variants (including 9 previously published cases) and identified 11 loss-of-function (LoF) variants (including nonsense, frameshift and intronic variants with predicted severe effect on splicing), seven compound LoF and deleterious missense variants, and one homozygous deleterious missense variant, c.5155G>C, p.(Ala1719Pro) (PMID: 32480058). The clinical manifestations associated with the missense variants were similar to those associated with other damaging *ALPK3* variants.

To note heterozygous LoF *ALPK3* variants have also been reported and are enriched in adults with cardiomyopathy (PMID: 34263907; PMID: 32480058; PMID: 33191771). Haploinsufficiency is the proposed mechanism. Herkert et al found notable differences between the clinical features associated with monoallelic and biallelic *ALPK3* cardiomyopathy, including absence or undetected extracardiac phenotypes. “Whether these differences reflect graded dose-responses to ALPK3 deficits or distinct mechanisms by which monoallelic or biallelic variants cause disease remains unknown.” (PMID: 32480058).

There does not appear to be a distinct mechanism between monoallelic and biallelic variants.

The disease mechanism is **loss of function** of *ALPK3* likely due to creation of premature stop codons, leading to nonsense-mediated decay or truncated proteins with partial or complete removal of the kinase domain (PMID:26846950; 27106955; 32480058; 21441111). Herkert et al predicted that missense variants could result in a conformational change that affects protein folding or flexibility, protein-protein or protein-DNA interaction, or the activity of the alpha-kinase domain (PMID: 32480058).

| Gene | *CACNA1C* |
| --- | --- |
| OMIM gene number | 114205 |
| Referral indication | Hypertrophic Cardiomyopathy (HCM) |
| Disease grouping | Rare syndromic disorder with hypertrophic cardiomyopathy - isolated LVH |
| **Disease name** | ***CACNA1C*-related Timothy syndrome** |
| MONDO ID | 0010979 |
| Gene disease validity (ClinGen) | DEFINITIVE |
| Inheritance | Autosomal dominant |
| Allelic requirement | Monoallelic autosomal |
| Inheritance modifiers | Typically *de novo* |
| Disease-associated variant consequence | Altered gene product sequence |
| Variant classes reported with evidence of pathogenicity | Missense |
| Restricted repertoire of pathogenic variants | A recurrent, *de novo* missense variant causing Classic Timothy Syndrome (p.Gly406Arg) has been described |
| PMIDs | 28211989; 25633834; NBK1403; 15863612; 15454078; 22106044; 26253506; 24728418; 31983240; NBK1403; 30681346; 33797204; 16360093 |

*CACNA1C*-related Timothy syndrome is caused by variants leading to **altered gene product sequence**.

Classic Timothy syndrome (TS1) is a very rare multisystem disorder characterized by marked QT prolongation, syndactyly, immune deficiency, seizures, congenital heart defects, hypertrophic cardiomyopathy, cognitive abnormalities, learning difficulties, and intermittent hypoglycaemia (PMID: 28211989; NBK1403). Infants can present with severe biventricular hypertrophy (PMID 30681346). In some individuals LVH may be the presenting feature, without recognised syndromic features, fulfilling a clinical diagnosis of HCM.

A recurrent, ***de novo*, missense variant** in *CACNA1C* was described in 13 Timothy syndrome patients, p.Gly406Arg in **exon 8A** (PMID 15454078; PMID: 15863612). The mechanism appears to be gain-of-function through failed channel inactivation. Boczek et al suggest Ca2+ mishandling may lead to ventricular hypertrophy (PMID: 26253506).

*CACNA1C* has a complex genomic structure that undergoes extensive alternative splicing. Splawski et al identified 2 patients with ***de novo* missense variants** in **exon 8** of an alternate splice form (p.Gly406Arg, analogous to the exon 8a variant, and p.Gly402Ser). This splice form represents 80% of all cardiac mRNAs. The patients were described as having atypical Timothy syndrome (TS2), presenting with a more severe cardiac phenotype (biventricular hypertrophy, moderate biventricular dysfunction, more severe QT prolongation and multiple arrythmias) and without syndactyly (PMID: 15863612; PMID: 25633834).

Boczek et al describe a novel variant **p.Arg518Cys-CACNA1C** as the probable pathogenic substrate for COTS [cardiac only Timothy syndrome]. The phenotype included LQTS, hypertrophic cardiomyopathy, congenital heart defects and sudden cardiac death. Follow-up cohort analysis revealed two additional pedigrees, with very similar phenotypes, having variants at the exact same amino acid position **(****p.Arg518Cys and** **p.Arg518His).** (PMID: 26253506)

Other missense variants in *CACNA1C* have been reported in association with isolated LQT (PMID: 26253506; PMID: 25633834; PMID:24728418). However as of 2020, the ClinGen Cardiovascular Domain Working Group have classified the strength of evidence supporting an association between *CACNA1C* and LQTS as moderate (PMID 31983240).

| Gene | *DES* |
| --- | --- |
| OMIM gene number | 125660 |
| Referral indication | Hypertrophic Cardiomyopathy (HCM) |
| Disease grouping | Rare syndromic disorder with hypertrophic cardiomyopathy - isolated LVH |
| **Disease name** | *DES*-related Myofibrillar myopathy |
| MONDO ID | 0011076 |
| Gene disease validity (ClinGen) | DEFINITIVE |
| Inheritance | Autosomal dominant; autosomal recessive |
| Allelic requirement | Monoallelic autosomal; biallelic autosomal |
| Inheritance modifiers | Typified by incomplete penetrance; Typified by age-related onset |
| Disease-associated variant consequence | Altered gene product sequence; absent gene product |
| Variant classes reported with evidence of pathogenicity | Splice acceptor variant NMD escaping; splice donor variant NMD escaping; frameshift variant NMD triggering; frameshift variant NMD escaping; stop gained NMD triggering; stop gained NMD escaping; missense variant; inframe deletion |
| Restricted repertoire of pathogenic variants | NA |
| PMIDs | 30681346; 19181099; 20718792; 29926427; 19433360; 23815709; 16217025; 11073539; 31718026; 9736733 |

*DES*-related myofibrillar myopathy is typically inherited as an autosomal dominant condition. Penetrance is incomplete with age-related onset, but simplex cases have often been found to be due to *de novo* heterozygous variants rather than recessive variants. Over 100 different variants have been reported, including missense variants, in-frame indels, and splice-site variants leading to exon skipping.

There is evidence for a possible genotype-phenotype correlation for heterozygous *DES* variants, with variants located in specific domains more likely to cause either a neurological or cardiac phenotype.

There is a wide range of cardiac phenotypes associated with *DES*, including hypertrophic cardiomyopathy, restrictive cardiomyopathy, dilated cardiomyopathy, arrhythmogenic cardiomyopathy, and left-ventricular non-compaction. Cardiac involvement can be the presenting feature, even without recognised syndromic features, fulfilling a clinical diagnosis of HCM.

The mechanism of autosomal dominant disease is likely a dominant-negative effect of protein-altering variants leading to abnormal intermediate filament aggregation.

The autosomal recessive phenotype has been associated with an earlier age of onset, and has been linked with combinations of protein-altering variants and/or null variants. It has been suggested that biallelic loss of function leads to the autosomal recessive phenotype, indicating a distinct mechanism of disease to the dominant phenotype. The heterozygous parents of patients with the recessive phenotype are reported as being clinically unaffected, suggesting that heterozygous loss of function is tolerated.

| Gene | *FHL1* |
| --- | --- |
| OMIM gene number | 300163 |
| Referral indication | Hypertrophic Cardiomyopathy (HCM) |
| Disease grouping | Rare syndromic disorder with hypertrophic cardiomyopathy - isolated LVH |
| **Disease name** | *FHL1*-related Emery-Dreifuss muscular dystrophy |
| MONDO ID | 0010680 |
| Gene disease validity (ClinGen) | DEFINITIVE |
| Inheritance | X-linked recessive |
| Allelic requirement | Monoallelic X hemizygous |
| Inheritance modifiers | Typified by age-related onset |
| Disease-associated variant consequence | Decreased gene product level; altered gene product sequence |
| Variant classes reported with evidence of pathogenicity | Splice region variant; frameshift variant NMD escaping; stop gained NMD escaping; stop lost; missense |
| Restricted repertoire of pathogenic variants | NA |
| PMIDs | 30681346; 19716112; 20186852; 32993534; 22523091 |

*FHL1*-related Emery-Dreifuss muscular dystrophy is inherited as an X-linked condition. Carrier females are usually unaffected, but some exhibit a cardiac and/or skeletal muscle phenotype.

The precise mechanism is unclear, but may be mediated by reduced function or dominant-negative effects of altered FHL1, associated with frameshift/truncating variants predicted to escape NMD, missense variants, or loss of the native stop codon.

*FHL1* pathogenic variants are responsible for a minority of Emery-Dreifuss muscular dystrophy cases (~1.2% according to GeneReviews). Hypertrophic cardiomyopathy is more typical of *FHL1*-related Emery-Dreifuss muscular dystrophy, while *LMNA* and *EMD* Emery-Dreifuss muscular dystrophy are more commonly associated with dilated cardiomyopathy.

There are reports of *FHL1* variants causing isolated hypertrophic cardiomyopathy, without skeletal myopathy.

| Gene | *FLNC* |
| --- | --- |
| OMIM gene number | 102565 |
| Referral indication | Hypertrophic Cardiomyopathy (HCM) |
| Disease grouping | Rare syndromic disorder with hypertrophic cardiomyopathy - isolated LVH |
| **Disease name** | FLNC-related Myofibrillar myopathy |
| MONDO ID | 0019150 |
| Gene disease validity (ClinGen) | DEFINITIVE |
| Inheritance | Autosomal dominant |
| Allelic requirement | Monoallelic autosomal |
| Inheritance modifiers | Typified by age-related onset |
| Disease-associated variant consequence | Altered gene product sequence |
| Variant classes reported with evidence of pathogenicity | Stop gained NMD escaping; missense variant; inframe deletion |
| Restricted repertoire of pathogenic variants | NA |
| PMIDs | 30681346; 15929027; 25351925; 19050726; 32022900; 26666891; 27908349; 31245841; 32112656; 20697107; 21135393 |

*FLNC*-related myofibrillar myopathy is inherited as an autosomal dominant condition with age-related onset. Myofibrillar myopathy and isolated hypertrophic cardiomyopathy have been associated with protein-altering variants in *FLNC* (missense, in-frame deletion, and nonsense predicted to escape NMD).

The specific mechanism of disease for pathogenic missense variants is not clear, but there is a cluster of HCM-related variants in the ROD2 domain, suggesting that these variants may affect interaction with the sarcomeric Z-disk. Variants associated with skeletal myopathy have been shown to induce intracellular aggregation of mutant FLNC.

Loss-of-function variants in *FLNC* have been associated with non-syndromic dilated and arrhythmogenic cardiomyopathy.

Sequencing of *FLNC* exons 46-48 is complicated by the presence of a pseudogene with >98% homology.

| Gene | *GLA* |
| --- | --- |
| OMIM gene number | 300644 |
| Referral indication | Hypertrophic Cardiomyopathy (HCM) |
| Disease grouping | Rare syndromic disorder with hypertrophic cardiomyopathy - isolated LVH |
| **Disease name** | ***GLA*-related Fabry disease** |
| MONDO ID | 0010526 |
| Gene disease validity (ClinGen) | DEFINITIVE |
| Inheritance | X-linked recessive |
| Allelic requirement | Monoallelic X hemizygous |
| Inheritance modifiers | NA |
| Disease-associated variant consequence | Decreased gene product level; altered gene product sequence |
| Variant classes reported with evidence of pathogenicity | Missense; inframe insertion; inframe deletion; splice donor; splice acceptor; frameshift; stop gained; intronic variants leading to cryptic splice site; structural variants |
| Restricted repertoire of pathogenic variants | NA |
| PMIDs | 18940466; 26937390; 6023233; 2539398; 32640076; NBK1292; 27560961; 30988410; 11322659; 34576250; 34776082; 32640076 |

Pathogenic *GLA* variants cause Fabry disease by **decreased gene product level or altered gene product sequence**. The disease mechanism is loss of function (PMID: 18940466; PMID 26937390; PMID 6023233; PMID 2539398). *GLA* pathogenic variants result in mRNA instability and/or severely truncated a-galactosidase A (a-Gal A) enzyme or an enzyme with markedly decreased activity (NBK1292)

Fabry disease is an X-linked lysosomal storage disease caused by pathogenic variants in the *GLA* gene leading to a greatly reduced or absent activity of a-Gal A, responsible for metabolizing glycosphingolipids. This condition is associated with a progressive accumulation of globotriaosylceramide (Gb3) and its deacylated form, globotriaosylsphingosine (lysoGb3), potentially affecting any organ or tissue (PMID: 32640076)

Fabry disease is inherited in an X linked manner. Heterozygous females typically have milder symptoms at a later age of onset than males. Rarely, they may be relatively asymptomatic throughout a normal life span or may have symptoms as severe as those observed in males with the classic phenotype (NBK1292).

Variant classes include **missense, nonsense, splice site, frameshift, in-frame deletions, and structural variants**. A recurrent intronic variant (c.640-801G\>A) is recognised as pathogenic and leads to aberrant mRNA splicing.

Many variants are unique however there are recognised recurrent variants also. (NBK1292; PMID 27560961; PMID 30988410; PMID 11322659; PMID: 34576250)

Pathogenic variants leading to complete loss of function of the gene product are usually associated with classic forms of the disease, whereas variants resulting in amino acid substitutions and residual enzyme activity can present atypically with either symptoms not specific to Fabry's (e.g. cardiomyopathy) or a milder phenotype and later onset. Attempts to correlate genotype with clinical presentation have been largely unsuccessful. (PMID 27560961; PMID: 18940466). Experts noted that conduction disease can be the presenting or only feature of disease.

The expert consensus on the management of Fabry Disease in 2020 suggested that “Assessment of plasma lyso-Gb_3_ should be considered for assessment of disease severity in FD patients or in the diagnostic algorithm for patients with *GLA* genetic variants of unknown significance.” (PMID: 32640076)

| Gene | *LAMP2* |
| --- | --- |
| OMIM gene number | 309060 |
| Referral indication | Hypertrophic Cardiomyopathy (HCM) |
| Disease grouping | Rare syndromic disorder with hypertrophic cardiomyopathy - isolated LVH |
| **Disease name** | *LAMP2*-related Danon disease |
| MONDO ID | 0010281 |
| Gene disease validity (ClinGen) | DEFINITIVE |
| Inheritance | X-linked dominant |
| Allelic requirement | Monoallelic X heterozygous |
| Inheritance modifiers | Typified by age-related onset |
| Disease-associated variant consequence | Decreased gene product level; altered gene product sequence |
| Variant classes reported with evidence of pathogenicity | Copy number variants; splice region variant; splice acceptor variant NMD triggering; splice donor variant NMD triggering; frameshift variant NMD triggering; stop gained NMD triggering; start lost; missense variant |
| Restricted repertoire of pathogenic variants | NA |
| PMIDs | 30681346; 10972294; 30857840; 20173215; 19588270; 19057086; 16217705; 15907287; 15673802 |

Pathogenic *LAMP2* variants cause Danon disease. Danon disease follows X-linked inheritance, with high penetrance of cardiomyopathy (dilated or hypertrophic) in hemizygous males. Onset is age-related, with an older age of onset in females than in hemizygous males, and males are more likely to have syndromic involvement (including skeletal myopathy and cognitive impairment).

In the majority of cases, the **mechanism is loss of function due to decreased gene product**, as a result of frameshift variants, nonsense variants, splice-site variants, and copy-number variants. Individual pathogenic missense variants have been reported. Some missense variants also cause a reduction in protein level, e.g. due to aberrant splicing, but there are pathogenic missense variants which have been shown to affect protein structure rather than reducing protein expression.

A possible transcript-specific mechanism has been suggested, with variants affecting exon 9b reported as causing a primarily skeletal muscle phenotype, with limited cardiac involvement.

| Gene | *PRKAG2* |
| --- | --- |
| OMIM gene number | 602743 |
| Referral indication | Hypertrophic Cardiomyopathy (HCM) |
| Disease grouping | Rare syndromic disorder with hypertrophic cardiomyopathy - isolated LVH |
| **Disease name** | ***PRKAG2*-related cardiomyopathy** |
| MONDO ID | 0010946 |
| Gene disease validity (ClinGen) | DEFINITIVE |
| Inheritance | Autosomal dominant |
| Allelic requirement | Monoallelic autosomal |
| Inheritance modifiers | NA |
| Disease-associated variant consequence | Altered gene product sequence |
| Variant classes reported with evidence of pathogenicity | Missense; inframe insertion |
| Restricted repertoire of pathogenic variants | NA |
| PMIDs | 26729852; 32259713; 32646570; 32646569; 12015471; 28009297; 11371514 |

*PRKAG2* pathogenic variants cause disease through **altered gene product sequence**.

PRKAG2 syndrome is a rare, early-onset **autosomal dominant inherited** disease, characterized by ventricular pre-excitation, supraventricular arrhythmias, and cardiac hypertrophy (PMID: 26729852; 32259713; 32646570).

There is a debate about penetrance but there appears to be variable expressivity of the clinical phenotype which may be variant specific (PMID: 32646569; 12015471).

*PRKAG2* variants have been recognized mainly in the context of patients with non-sarcomeric familial hypertrophic cardiomyopathy associated with Wolff-Parkinson-White (WPW) syndrome (PMID: 26729852). The *PRKAG2* gene encodes for the 5’ Adenosine Monophosphate-Activated Protein Kinase (AMPK), specifically for its γ2 regulatory subunit (PRKAG2). *PRKAG2* pathogenic variants are suspected to modify the tri-dimensional structure of AMPK, altering its affinity for AMP and **modifying the enzyme activity** (PMID: 32259713; 28009297).

Nearly all pathogenic variants are **missense** (PMID: 32259713; 32646569; 12015471; 28009297).

Blair et al. documented a TTA codon insertion in exon 5 in 2001 in a family affected by severe early onset cardiomyopathy and multiple sudden deaths in early adult life (PMID: 11371514). The variant is in a highly conserved region and co-segregated with disease. This has not been reported again on ClinVar.

Lopez-Sainz A et al reported one frameshift and one intronic variant (the rest missense) in their multi-centre retrospective study of 90 PRKAG2 variant carriers in 2020 (PMID: 32646569). However, the significance of truncating variants remains uncertain with insufficient evidence to support loss of function as a mechanism of disease. ClinGen found no evidence to support haploinsufficiency as a mechanism (https://search.clinicalgenome.org/kb/gene-dosage/HGNC:9386).

Two commonly reported variants are C.905G>A (p.Arg302Gln) and c.1463A>T (p.Asn488Ile). Lopez-Sainz A et al found they were present in 44% of the patients included in the cohort (PMID: 32646569).

| Gene | *PTPN11* |
| --- | --- |
| OMIM gene number | 176876 |
| Referral indication | Hypertrophic Cardiomyopathy (HCM) |
| Disease grouping | Rare syndromic disorder with hypertrophic cardiomyopathy - isolated LVH |
| **Disease name** | *PTPN11*-related Noonan syndrome |
| MONDO ID | 0008104 |
| Gene disease validity (ClinGen) | DEFINITIVE |
| Inheritance | Autosomal dominant |
| Allelic requirement | Monoallelic autosomal |
| Inheritance modifiers | NA |
| Disease-associated variant consequence | Altered gene product sequence |
| Variant classes reported with evidence of pathogenicity | Missense; inframe deletion |
| Restricted repertoire of pathogenic variants | NA |
| PMIDs | 30681346; 23312968; 21269411; 11992261; 16358218; 14974085; 15521065; 15240615; 18348260; 19760651; 24739123; 25974318; 21533187 |

*PTPN11*-related Noonan syndrome is inherited as an autosomal dominant condition with variable expressivity. Pulmonary stenosis is the most common cardiac manifestation. The penetrance of hypertrophic cardiomyopathy in *PTPN11*-related Noonan syndrome has been reported as 20-30%.

There is no definite evidence of *PTPN11* variants causing isolated cardiac disease without other features of Noonan syndrome, but the syndromic phenotype can be subtle and may be underdiagnosed. This suggests that cardiac involvement can be the initial presentation in some cases, and *PTPN11* should be considered in the differential diagnosis of HCM.

*PTPN11* variants are the most common cause of Noonan syndrome, accounting for approximately 50% of cases. The mechanism is likely gain of function due to missense variants (particularly variants disrupting interaction between the N-SH2 and PTP domains), leading to activation of the RAS-MAPK pathway. There are at least two reports of Noonan syndrome due to single-residue in-frame deletions in *PTPN11*.

There are reports of structural duplications including *PTPN11* causing Noonan syndrome, but all patients reported to date have had a relatively non-specific phenotype and large duplications encompassing other genes. It is therefore not clear whether gene duplication specifically causes Noonan syndrome.

Allelic disorders include Noonan syndrome with multiple lentigines (NSML, previously known as LEOPARD syndrome), caused by dominant-negative *PTPN11* variants, and metachondromatosis, caused by loss-of-function *PTPN11* variants.

| Gene | *PTPN11* |
| --- | --- |
| OMIM gene number | 176876 |
| Referral indication | Hypertrophic Cardiomyopathy (HCM) |
| Disease grouping | Rare syndromic disorder with hypertrophic cardiomyopathy - isolated LVH |
| **Disease name** | *PTPN11*-related Noonan syndrome with multiple lentigines |
| MONDO ID | 0100082 |
| Gene disease validity (ClinGen) | DEFINITIVE |
| Inheritance | Autosomal dominant |
| Allelic requirement | Monoallelic autosomal |
| Inheritance modifiers | NA |
| Disease-associated variant consequence | Altered gene product sequence |
| Variant classes reported with evidence of pathogenicity | Missense |
| Restricted repertoire of pathogenic variants | NA |
| PMIDs | 12058348; 12161596; 17697839; 11992261; 16358218; 15121796; 16377799; 33354767 |

Noonan syndrome with multiple lentigines (NSML, previously known as LEOPARD syndrome) is inherited as an autosomal dominant condition. Missense variants in *PTPN11* are responsible for the majority of cases. Rare cases have been attributed to variants in *RAF1, BRAF*, and *MAP2K1.*

There is no definite evidence of *PTPN11* variants causing isolated cardiac disease without other features of NSML, but the syndromic phenotype can be subtle and may be underdiagnosed. This suggests that cardiac involvement can be the initial presentation in some cases, and *PTPN11* should be considered in the differential diagnosis of HCM.

*PTPN11* variants associated with NSML are clustered in the phosphotyrosine phosphatase (PTP) domain and have been shown to inhibit phosphatase activity. Since null variants have not been associated with the NSML phenotype, it has been suggested that NSML is caused by a dominant-negative effect rather than simple loss of function, possibly due to altered interaction between PTPN11 and binding partners.

Allelic disorders include Noonan syndrome, caused by gain-of-function variants in *PTPN11*, and metachondromatosis, caused by loss-of-function *PTPN11* variants.

| Gene | *RAF1* |
| --- | --- |
| OMIM gene number | 164760 |
| Referral indication | Hypertrophic Cardiomyopathy (HCM) |
| Disease grouping | Rare syndromic disorder with hypertrophic cardiomyopathy - isolated LVH |
| **Disease name** | *RAF1*-related Noonan syndrome |
| MONDO ID | 0012690 |
| Gene disease validity (ClinGen) | DEFINITIVE |
| Inheritance | Autosomal dominant |
| Allelic requirement | Monoallelic autosomal |
| Inheritance modifiers | NA |
| Disease-associated variant consequence | Altered gene product sequence |
| Variant classes reported with evidence of pathogenicity | Missense; inframe deletion |
| Restricted repertoire of pathogenic variants | NA |
| PMIDs | 30681346; 17603482; 17603483; 30762279; 29271604; 24782337; 22786616; 25974318 |

*RAF1* variants are responsible for ~5% of Noonan syndrome cases. *RAF1*-related Noonan syndrome is inherited as an autosomal dominant condition.

The mechanism is likely *RAF1* gain of function, due to missense variants leading to activation of the Ras-MAPK pathway.

The penetrance of hypertrophic cardiomyopathy is higher in patients with *RAF1* variants, compared to other forms of Noonan syndrome. There is no definite evidence of *RAF1* variants causing isolated cardiac disease without other features of Noonan syndrome, but *RAF1* variants have been identified in multiple patients who were being investigated for HCM, indicating that the syndromic phenotype can be subtle and may be underdiagnosed. Since cardiac involvement can be the initial presentation in some cases, *RAF1* should be considered in the differential diagnosis of HCM.

There is evidence for a genotype-phenotype correlation, with variants in the CR2 domain of *RAF1* more likely to be associated with HCM. There is a further mutational hotspot in the kinase domain of CR3, but pathogenic variants have also been identified outside these hotspots.

There are reports of patients with chromosomal duplications including *RAF1*, but their phenotype is relatively non-specific and there is no definitive evidence that whole-gene duplications cause Noonan syndrome.

| Gene | *RIT1* |
| --- | --- |
| OMIM gene number | 609591 |
| Referral indication | Hypertrophic Cardiomyopathy (HCM) |
| Disease grouping | Rare syndromic disorder with hypertrophic cardiomyopathy - isolated LVH |
| **Disease name** | *RIT1*-related Noonan syndrome |
| MONDO ID | 0014143 |
| Gene disease validity (ClinGen) | DEFINITIVE |
| Inheritance | Autosomal dominant |
| Allelic requirement | Monoallelic autosomal |
| Inheritance modifiers | NA |
| Disease-associated variant consequence | Altered gene product sequence |
| Variant classes reported with evidence of pathogenicity | Missense |
| Restricted repertoire of pathogenic variants | NA |
| PMIDs | 30681346; 23791108; 26446362; 27101134; 25959749 |

*RIT1* variants are responsible for ~5% of Noonan syndrome cases. *RIT1*-related Noonan syndrome is inherited as an autosomal dominant condition.

The penetrance of hypertrophic cardiomyopathy is higher in patients with *RIT1* variants, compared to other forms of Noonan syndrome (estimated at 70-75%). There is no definite evidence of *RIT1* variants causing isolated cardiac disease without other features of Noonan syndrome, but the syndromic phenotype can be subtle and may be underdiagnosed. This suggests that cardiac involvement can be the initial presentation in some cases, and *RIT1* should be considered in the differential diagnosis of HCM.

The mechanism is likely *RIT1* gain of function, due to missense variants leading to activation of the Ras-MAPK pathway. The precise mechanism by which missense variants lead to gain of function remains unclear. There is clustering of variants, particularly in the switch II region.

| Gene | *TTR* |
| --- | --- |
| OMIM gene number | 176300 |
| Referral indication | Hypertrophic Cardiomyopathy (HCM) |
| Disease grouping | Rare syndromic disorder with hypertrophic cardiomyopathy - isolated LVH |
| **Disease name** | ***TTR*-related hereditary ATTR amyloidosis** |
| MONDO ID | 0019441 |
| Gene disease validity (ClinGen) | DEFINITIVE |
| Inheritance | Autosomal dominant |
| Allelic requirement | Monoallelic autosomal |
| Inheritance modifiers | Typified by incomplete penetrance |
| Disease-associated variant consequence | Altered gene product sequence |
| Variant classes reported with evidence of pathogenicity | Missense; inframe insertion; inframe deletion |
| Restricted repertoire of pathogenic variants | NA |
| PMIDs | 25604431; NBK1194; 28213611; 15185500; 30328212; 15930086; 9191784; 29941560; 32969287 |

The mechanism in *TTR*-related hereditary ATTR amyloidosis is due to **altered gene product sequence**.

The mechanism appears to be **gain-of-function**; pathogenic variants cause either tetramer dissociation or monomer denaturation, which both contribute to the formation of amyloid fibrils in tissue (PMID 25604431; NBK1194).

Of note, wildtype *TTR* can also cause an age-related late onset form of amyloidosis where patients almost exclusively present with cardiac involvement.

*TTR*-related hereditary ATTR amyloidosis is characterised by **autosomal dominant inheritance** with **incomplete penetrance**. There are reports of homozygous and compound heterozygous variants. Biallelic variants appear to cause a more severe phenotype (PMID: 28213611; 15185500; 30328212), however one study reported no difference in phenotype between homozygous and heterozygous carriers (PMID: 15930086).

*TTR*-related hereditary ATTR amyloidosis includes the phenotypes ATTR amyloid neuropathy, ATTR cardiac amyloidosis, ATTR leptomeningeal/CNS amyloidosis. Phenotype varies considerably; individuals can present with multi-systemic phenotypes including, polyneuropathy, carpal tunnel syndrome, cardiomyopathy, gastrointestinal features, autonomic insufficiency, and renal insufficiency. "The prevalence of HCM and/or RCM in Hereditary ATTR amyloidosis is high but exact percentages have not been accurately defined due the high prevalence of HCM and RCM (~25%) development in senile cardiac amyloidosis, which occurs due to accumulation of wildtype TTR accumulation in the heart with age." (ClinGen summary)

>100 different variants have been reported in the Hereditary Amyloidosis Registry (http://www.amyloidosismutations.com).

Variants are all **missense** apart from one inframe deletion of a valine residue in exon 4 (Plasma transthyretin levels in the mutant gene carriers measured by nephelometry were very low) and one 6 nucleotide duplication in exon 3. The duplication was reported to be associated with a particularly aggressive phenotype and although it did not alter the protein secondary or tertiary structure, it decreased the stability of the TTR monomer and tetramer (PMID: 9191784; PMID: 29941560).

Several variants including TTR-V30M, TTR-T60A, and TTR-V122L are commonly associated with cardiac amyloidosis (PMID 28739313). There are other genotype phenotype correlations (PMID 25604431; NBK1194).

The ATTRVal122Ile variant (also reported as pVal142Ile) is a risk allele in patient subpopulations of African American descent. The variant in this population has a high prevalence (estimated 3-4%) and patients are at particular risk for developing cardiac-related ATTR. (PMID: 28213611).

A systematic review in 2020 evaluating specific therapies for transthyretin cardiac amyloidosis supported the use of tafamidis (a tetramer stabiliser) and noted that other novel therapeutic targets including transthyretin gene silencers are currently under investigation (PMID: 32969287).
